# Beyond Signal Detection: Sequential Target Trial Emulations to Confirm Previously Detected Adverse Drug Event Signals for Atorvastatin in Older Medicare Beneficiaries

**DOI:** 10.64898/2026.08.12.26360302

**Authors:** Christopher G. Rowan, Camille Maringe, Nancy A. Dreyer, Macarius M. Donneyong, Gregory L. Peck, Steven M. Brunelli

## Abstract

**Background and objective:** High dimensional, hypothesis free signal detection in claims data can identify adverse drug events (ADEs), yet these exploratory signals remain vulnerable to false positives and residual confounding. Rigorous internal confirmation using causal inference methods is therefore a critical intermediate step in a staged pharmacovigilance paradigm before external validation. We aimed to confirm or refute previously detected ADE signals associated with atorvastatin initiation among older adults after myocardial or cerebral infarction.

**Methods:** Internal confirmatory sequential target trial emulations were conducted using Medicare fee-for-service claims (2017–2019). Eligible participants were statin naïve beneficiaries aged ≥ 65 years hospitalized for myocardial or cerebral infarction. Up to 14 nested daily trials were constructed beginning on the discharge date. Pragmatic treatment strategies compared atorvastatin initiation with initiation of a different new outpatient medication. Per protocol effects were estimated using inverse probability weighted Fine-Gray models treating death as a competing risk. Previously detected ADE signals and clinically coherent alternatives within the same outcome families were evaluated according to prespecified dual criteria controlling for the false discovery rate and residual bias. Absolute risks and numbers needed to harm (NNH) were reported. A prespecified sensitivity analysis restricted inference to the first two trials with optimal covariate balance; a post hoc analysis examined effect modification by concomitant high risk antithrombotic therapy.

**Results:** Of 70,130 eligible patients, 39,948 initiated atorvastatin (81% high intensity) and 19,182 initiated a different new medication. After weighting, baseline characteristics were closely balanced. Associations meeting dual confirmatory criteria formed a gradient of support. The strongest findings were early (days 1 to 30) acute hemorrhagic cerebrovascular disease (sHR 2.20, 95% CI 1.35-3.58; NNH 351) and acute hepatic failure (sHR 1.72, 1.16-2.55; NNH 468), both robust to high risk antithrombotic therapy and to the restricted trial set with optimal covariate balance. Intermediate support was observed for biliary tract disease (women; sHR 1.45, 1.14-1.85; NNH 102) and musculoskeletal injuries (men; sHR 1.66, 1.24-2.21; NNH 71). Tentative evidence was observed for cardiac valve disorders (attenuated in the absence of high risk antithrombotic therapy) and sensory symptoms (non-White patients). Prediabetes and posthemorrhagic anemia were not confirmed.

**Conclusions:** Several previously detected ADE associations with atorvastatin initiation met dual confirmatory criteria and formed a gradient of confirmatory support. Strongest evidence supported early hemorrhagic cerebrovascular disease, hepatic dysfunction, and musculoskeletal injuries. Absolute excess risks were modest yet clinically relevant to a high risk post-infarction population (NNH 71-468) and must be interpreted alongside the established benefits of high intensity statin therapy (number needed to treat to prevent one event (NNT) 28-93). These findings support a two stage active pharmacovigilance paradigm (signal detection followed by rigorous confirmation) and justify heightened clinical vigilance for the confirmed events, while underscoring the need for external validation in independent populations and data sources.

**KEY POINTS:** *Question:* Can previously detected signals of adverse drug events associated with atorvastatin initiation in older adults be confirmed under more rigorous dual confirmatory criteria?

*Findings:* In this internal confirmatory Medicare claims study using sequential target trial emulation, associations meeting dual confirmatory criteria formed a gradient of support. The strongest findings were early acute hemorrhagic cerebrovascular disease (sHR 2.20) and acute hepatic failure (sHR 1.72), both robust to high risk antithrombotic therapy and to the optimally balanced early trials. Intermediate support was observed for biliary tract disease and musculoskeletal injuries. Weaker evidence was observed for cardiac valve disorders (attenuated in the absence of high risk antithrombotic therapy) and sensory symptoms. Prediabetes and posthemorrhagic anemia were not confirmed.

*Meaning:* Although atorvastatin remains foundational for secondary cardiovascular prevention (NNT 28–93 to prevent one event), these findings support heightened clinical vigilance for early hemorrhagic, hepatic, and musculoskeletal events in older adults after myocardial or cerebral infarction, while underscoring the need for external validation.

**PLAIN LANGUAGE SUMMARY:** Older adults who start atorvastatin after a heart attack or stroke may face a higher risk of certain serious side effects, according to this large study of Medicare insurance records.

Researchers previously identified possible safety signals for atorvastatin using advanced methods that mimic clinical trials with real world data. This follow up study carefully reexamined those signals with stricter eligibility rules, more precise outcome definitions, and statistical techniques designed to reduce false findings.

The analysis confirmed a range of associations that varied in strength. The strongest evidence pointed to higher rates of bleeding in the brain (especially in the first 30 days) and acute liver failure. Intermediate evidence supported bile duct problems and muscle strains or joint injuries. Weaker evidence was found for certain heart valve problems and for dizziness or sensory symptoms. Some earlier possible signals, such as prediabetes, were not confirmed.

Although atorvastatin remains an important treatment to prevent future heart attacks and strokes, these findings suggest that older adults starting the drug may benefit from close monitoring for early bleeding in the brain, liver problems, and muscle or joint injuries, and that suspicion for these should be heightened when suggestive symptoms present following atorvastatin initiation. Patients and clinicians should discuss these potential risks, remain alert to early warning signs, and recognize that further studies in other populations are still needed to confirm the results.

## INTRODUCTION

Rare, delayed, and context dependent adverse drug events (ADEs) are usually detected only after regulatory approval.^1^ Preapproval trials, constrained by modest sample sizes, brief follow-up, and narrow eligibility criteria, possess limited statistical power to detect such events.^2–9^ This gap is particularly consequential for older adults, who account for the largest share of medication use, experience the highest burden of competing risks, and exhibit the most pronounced age-related alterations in pharmacokinetics and pharmacodynamics, yet remain systematically underrepresented in the trials that underpin marketing authorization.^7,10–20^

Although spontaneous reporting systems have identified important safety problems, they lack a defined denominator, suffer from variable report quality, and remain vulnerable to substantial reporting bias.^21^ Population based claims and electronic health record databases furnish both a clearly enumerated source population and rich longitudinal clinical detail, thereby offering opportunity for rigorous pharmacovigilance, which unfortunately has not yet been fully realized. Even within these data sources, safety signals detected by high dimensional screening remain susceptible to residual confounding, outcome misclassification, multiplicity, and discovery stage analytic choices. Robust confirmation therefore demands a prespecified evaluation in which hypotheses are fixed in advance, eligibility criteria are expanded, the false discovery rate is controlled more stringently, and quantitative bias analysis is applied systematically.

Building on a prior signal detection study that identified associations between atorvastatin initiation and a number of clinical sequelae,^22,23^ we conducted internal confirmatory sequential target trial emulations that: 1) were hypothesis-driven, 2) utilized the same study population as the signal detection study (i.e., were internal validation studies), 3) were systematic in that each followed the same rules based sequence, and 4) utilized a pragmatic active comparator design mirroring routine post discharge care. This paradigm subjected preliminary signals to increasingly restrictive eligibility criteria, refined outcome definitions, stricter control of the false discovery rate, and formal quantitative bias analysis. The underlying hypothesis for each signal was effectively: Among older, previously statin-naïve Medicare beneficiaries discharged following myocardial infarction or ischemic stroke, *what is the per protocol effect of initiating atorvastatin, versus initiating different outpatient medications within two weeks post-discharge, on the subdistribution hazard ratio of the event, treating death as a competing risk?*

## METHODS

### Study Population and Sequential Trial Framework

The internal confirmatory analyses used the same study population and sequential target trial framework as the preceding signal detection study.^23^ Eligible participants were statin naïve Medicare fee-for-service beneficiaries aged ≥ 65 years who were hospitalized for myocardial infarction or cerebral infarction (primary diagnosis, length of stay ≥ 3 days) and discharged home between January 1, 2017 and December 31, 2019 (study period). This hospitalization defined the index (time zero) event.^22^

For each outcome evaluated, up to 14 nested daily trials (Trials 0–13) were conducted, beginning on the discharge date, which served as the start date for Trial 0. At each trial origin, eligibility required that beneficiaries were alive, free of prior statin exposure, continuously enrolled in Medicare Parts A, B, and D, free of the outcome under study, and without contraindications to atorvastatin (**Figure 1**). These criteria synchronized eligibility, treatment assignment, and the start of follow-up with the target trial protocol (supplemental material **Table S1**).

**Figure 1.**
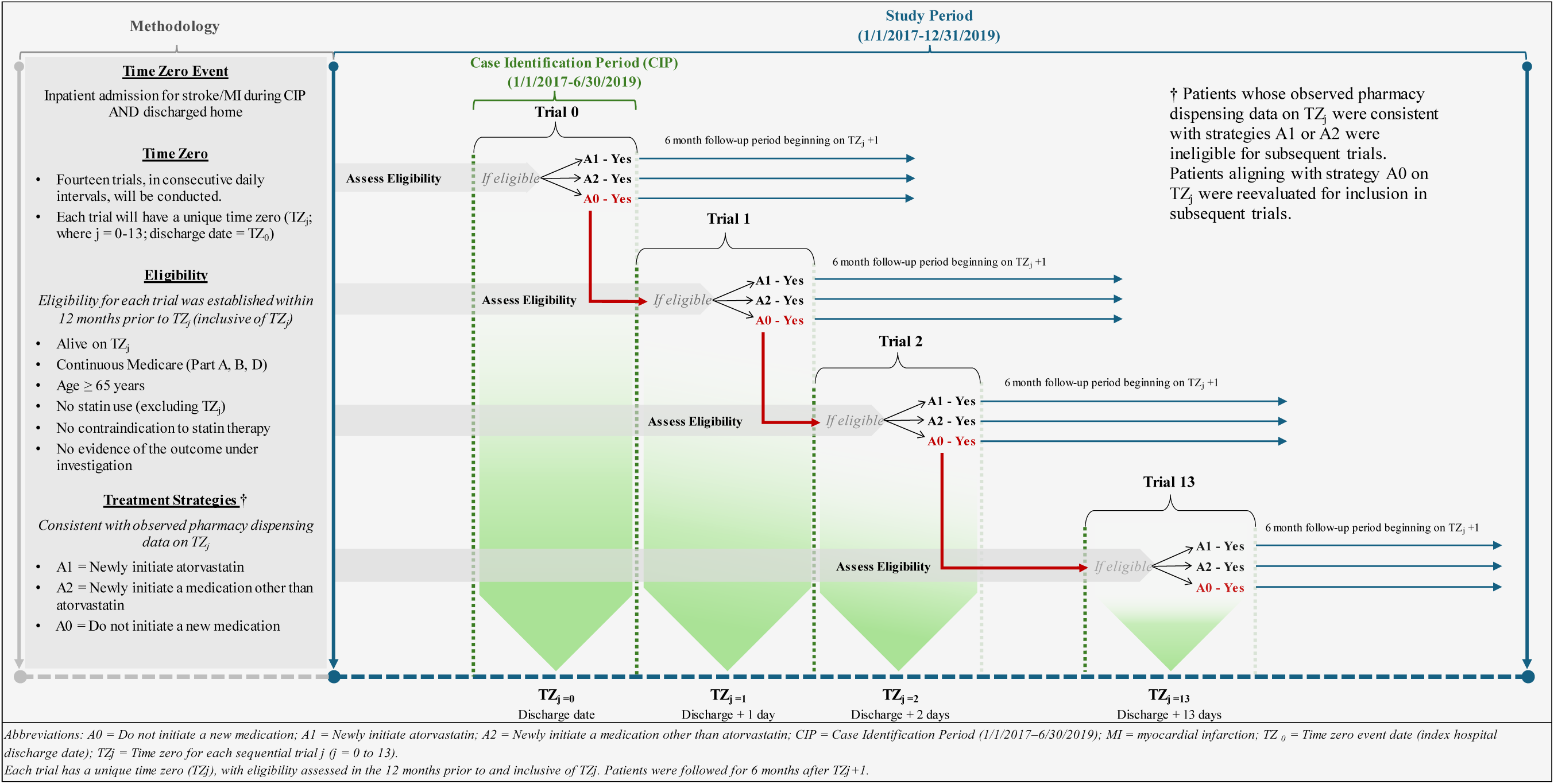
Study Schema: Target Trial Emulation Sequential Trials Approach A1

### Treatment Strategies

On each trial start date, treatment strategies were “assigned” according to observed pharmacy dispensings. Three pragmatic strategies were defined to reflect post discharge real world clinical decisions in a post-infarction population: (treatment strategy A1) initiation of atorvastatin (with any concurrent or other newly prescribed therapies); (treatment strategy A2) initiation of a different new outpatient medication (with any concurrent or other newly prescribed therapies); and (treatment strategy A0) no new medication initiation.

It should be noted that newly initiated medications other than atorvastatin (used to classify the A2 treatment strategy) consisted primarily of cardiovascular medications, as would be expected in a post-infarction population. A random selection of A2 medications used to classify discontinuation is provided in supplemental material **Table S2**; the distribution of medications initiated during the six-month follow-up period under both A1 and A2 strategies appears in supplemental material **Table S9** and demonstrates broadly comparable utilization patterns across arms.

Beneficiaries whose pharmacy records were consistent with A1 or A2 were included in the first trial under that treatment strategy and became ineligible for subsequent trials. Those assigned to strategy A0 remained eligible for subsequent trials until they met the criteria for A1 or A2. Sequential trial nesting synchronized eligibility determination, treatment assignment, and the start of follow-up with the observable decision to initiate therapy, thereby eliminating immortal time bias and survivor bias that would arise in a single trial design indexed on the date of medication initiation.

### Follow-up and Censoring

Within each nested trial (for each outcome), follow-up began the day after the trial start date and continued for up to 182 days or until the outcome or a censoring event. Within this period, we pre-specified intervals to capture possible transient risks (i.e., 1–30, 31–91, and 92–182 days). The trial start date served as the reference for baseline covariate ascertainment, confirmation of outcome absence, eligibility verification, and treatment assignment. Beneficiaries who experienced the outcome on the trial start date were excluded from that trial and all subsequent trials.

Censoring was classified as informative (death), administrative (end of study or loss of Medicare enrollment), or artificial (deviation from the assigned treatment strategy under the per protocol framework). Treatment strategy deviation included initiation of a non-atorvastatin statin, a switch between strategies, or discontinuation of the assigned treatment strategy (classified by a dispensing gap > 45 days). For strategy A2, one newly dispensed medication was randomly selected to define discontinuation. To maintain symmetry, switching from A1 to A2 was randomly generated according to the observed frequency of A2 to A1 switches, thereby reflecting the heterogeneous composition of medications within treatment strategy A2.

### Outcomes

The internal confirmatory analyses evaluated the ADE signals that exceeded the detection threshold in the discovery phase,^23^ together with refined outcomes drawn from the same clinical family (see **Table 1** and additional details in supplemental material **Table S3**). Within each outcome family, a hybrid definitional strategy was applied. At least one outcome retained the original Clinical Classifications Software Refined (CCSR) category and code list (with expanded exclusions), while complementary outcomes were classified using published outcome definitions or Sentinel Initiative outcome algorithms and code lists. This approach preserved continuity with the discovery signals while permitting evaluation of refined and alternative outcome definitions within the same outcome family. Exclusion criteria were applied primarily at the family level; for selected outcomes within the abnormal laboratory findings family (including acute hepatic failure), exclusions were enforced at the sub-family level.

**Table 1.** Outcome Definitions.

| Definitions of Outcomes: Data Sources, Coding Algorithms, and Exclusion Criteria |  |  |  |
| --- | --- | --- | --- |
| Outcomes Assessed | Outcome Source | Code List | Exclusion criteria |
| **Bleeding events** |  |  |  |
| Acute posthemorrhagic anemia* | CCSR | CCSR BLD004 | Exclude if baseline evidence of bleeding event<sup>†</sup> |
| Gastrointestinal bleeding | Sentinel | See supplemental material for details |  |
| Major extracranial bleeding | Sentinel | See supplemental material for details |  |
| Acute hemorrhagic cerebrovascular disease* | CCSR | CCSR CIR021 |  |
| Intracranial hemorrhage (traumatic & non-traumatic) | Sentinel | See supplemental material for details |  |
| Composite: bleed | - | Composite of all outcomes in this category |  |
| **Cardiac valve events** |  |  |  |
| Nonrheumatic and unspecified valve disorders* | CCSR | CCSR CIR003 | Exclude if baseline evidence of cardiac valve disorder or valve intervention<sup>†</sup> |
| Nonrheumatic and unspecified valve disorders plus imaging | CCSR | CCSR CIR003 AND cardiac imaging |  |
| Cardiac valve intervention procedure | Sentinel | See supplemental material for details |  |
| Composite: valve disorder or intervention | - | Composite of all outcomes in this category |  |
| **Sprains, strains, and tendon injury** |  |  |  |
| Sprains and strains, initial encounter* | CCSR | CCSR INJ024 | Exclude if baseline evidence of sprain, strain or tendon injury<sup>†</sup> |
| Sprain | Published study | See supplemental material for details |  |
| Strain | Published study | See supplemental material for details |  |
| Injury of muscle and tendon | Published study | See supplemental material for details |  |
| Tendon rupture and tendinopathy | Published study | See supplemental material for details |  |
| Composite: sprain/strain/tendon injury | - | Composite of all outcomes in this category |  |
| **Dizziness** |  |  |  |
| General sensation/perception signs and symptoms* | CCSR | CCSR SYM015 | Exclude if baseline evidence of perception symptoms or dizziness<sup>†</sup> |
| Dizziness and giddiness | Published study | See supplemental material for details |  |
| Vestibular disorders | Published study | See supplemental material for details |  |
| **Abnormal symptoms** |  |  |  |
| Hyperkalemia | Published study | See supplemental material for details | Outcome or Renal dysfunction<sup>†</sup> |
| Rhabdomyolysis | Sentinel | See supplemental material for details |  |
| Hyponatremia | Published study | See supplemental material for details |  |
| Proteinuria/hematuria | CCSR | CCSR GEN009, GEN010 | Renal dysfunction<sup>†</sup> |
| Nephritis; nephrosis; renal sclerosis | CCSR | CCSR GEN001 |  |
| Acute renal failure | CCSR | CCSR GEN002 |  |
| Cardiac dysrhythmias and conduction disorders | CCSR | CCSR CIR016 CIR017 | Cardiac dysrhythmias or cardiac arrest<sup>†</sup> |
| Cardiac arrest and ventricular fibrillation | CCSR | CCSR CIR018 |  |
| Drug-induced liver injury (DILI) | Published study | See supplemental material for details | Hepatic dysfunction, injury or failure<sup>†</sup> |
| Other specified and unspecified liver disease | CCSR | CCSR DIG019 |  |
| Biliary tract disease | CCSR | CCSR DIG017 |  |
| Acute hepatic failure – narrow definition | Published study | See supplemental material for details |  |
| Hepatic failure – broad definition | CCSR | CCSR DIG018 |  |
| **Hyperglycemic events** |  |  |  |
| Prediabetes* | CCSR | CCSR SYM018 | Exclude if baseline evidence of any outcome or an antidiabetic agent<sup>†</sup> |
| Diabetes | Sentinel | See supplemental material for details |  |
| Hyperglycemia | Sentinel | See supplemental material for details |  |
| Composite: diabetes | - | Composite of all outcomes in this category |  |

Notes: CCSR = Clinical Classifications Software Refined (AHRQ/HCUP for ICD-10-CM diagnoses). Sentinel = validated algorithms and code lists from the FDA Sentinel Initiative. Outcomes in italics with an asterisk (\*) represent the specific adverse drug event (ADE) signals that were detected in a previously conducted signal detection study. Exclusion criteria were applied to restrict analyses to incident events by excluding individuals with baseline evidence of the outcome or related conditions. † = Exclusion criteria were applied to all outcomes in the category.

All diagnosis based outcomes were ascertained from ICD-10-CM codes recorded on inpatient, emergency department, and outpatient claims (diagnosis position unrestricted). Procedure codes were used in two distinct ways. Cardiac valve intervention was defined solely from inpatient ICD-10-PCS and outpatient CPT codes, whereas the composite outcome for nonrheumatic valve disease with recent cardiac imaging required both a qualifying diagnosis code and evidence of recent cardiac imaging procedure. Procedure codes were also used where relevant within the sprains and strains family (e.g., tendon repair surgery). An incident event required the absence of the relevant family or component level exclusion codes in the 12 months preceding the trial start date (inclusive of the trial start date) and the presence of at least one qualifying code during follow-up; only the first outcome occurrence was analyzed. Baseline evidence was established by one inpatient or emergency department claim or by two outpatient claims on distinct dates. Composite outcomes that aggregated related component events within each family were also constructed.

### Baseline Covariates

Baseline covariates were ascertained in the 12-month period preceding each trial start date (inclusive of the trial start date, except where noted). Demographic variables (age, sex, race, dual Medicare–Medicaid eligibility, and disability status) were obtained from the Beneficiary Summary File. Healthcare utilization included counts of inpatient admissions, inpatient days, emergency department visits, unique service dates, and total medical expenditures.

Comorbidities were identified from inpatient, emergency department, and outpatient claims by mapping ICD-10-CM codes to CCSR categories (one inpatient or emergency department claim, or two distinct outpatient claims). The Charlson Comorbidity Index was calculated using a weighted algorithm. Baseline pharmacotherapy utilization was ascertained from Part D claims (excluding the trial start date) and linked to First Databank and AHFS classifications, yielding 417 baseline medication variables across 20 major therapeutic classes. Inpatient procedures were captured with ICD-10-PCS codes (320 CCSR categories); outpatient procedures were identified with CPT/HCPCS codes (245 CCSR categories).

### Statistical Analysis

For each outcome, sequential target trials were constructed among eligible statin naïve beneficiaries. The primary contrast compared initiation of atorvastatin (strategy A1) with initiation of any other new outpatient medication (strategy A2). Strategy A0 (no new medication) was retained to preserve sequential eligibility, but was omitted from the exposure contrast to reduce channeling bias. Trials were retained only when both arms (A1 and A2) contained at least 40 patients. Beneficiaries with baseline evidence of the outcome were excluded from that trial and all subsequent trials. Outcomes occurring on the same day as death were classified as outcomes.

Stabilized inverse probability of treatment weights were estimated by logistic regression, and stabilized inverse probability of censoring weights by Cox regression. Baseline covariates for both models were selected adaptively within each trial by successive exclusion of variables with zero variance, perfect prediction (of treatment or censoring), pairwise correlation > 0.95, or cell frequencies < 20. The product of the stabilized weights was truncated at 10 and applied to data stacked across retained trials.^24–26^

The per protocol effect was estimated as the subdistribution hazard ratio from weighted Fine-Gray models treating death as a competing risk, with robust variance clustered at the patient level. Absolute risks, risk differences, and numbers needed to harm (NNH) were also reported.^27^ Covariate balance and weight distributions were examined as diagnostics.

Prespecified stratified analyses evaluated time windows (days 1–30, 31–91, 92–182) and demographic subgroups (age 65–74 vs ≥ 75 years, sex, and White vs non-White race). A prespecified sensitivity analysis restricted inference to Trials 0 and 1, which achieved superior covariate balance (maximum standardized mean difference < 0.1), thus providing stronger control for measured confounding. These sensitivity analyses change the causal question to initiation of atorvastatin within a day of discharge. A formal dose response analysis was not performed because approximately 81% of atorvastatin initiators received high intensity therapy (40 mg or 80 mg); with so few patients on lower intensity regimens, dose stratified estimates would have been imprecise and of limited interpretive value.

### Statistical Analysis – Outcome Confirmation Procedure

This study sought to confirm or refute a limited set of ADE signals identified in the prior detection analysis.^23^ Each outcome in this internal confirmation study constituted a prespecified hypothesis. Multiplicity was controlled within each outcome because the objective was confirmation of prioritized signals rather than discovery; an across-outcome correction would have been unnecessarily conservative.

For each outcome we examined up to ten analyses organized into three families of tests: (1) a single overall estimate across the full follow-up period, (2) up to three time stratified estimates (to identify possible violations of the proportional hazards assumption), and (3) up to six subgroup estimates by age, sex, and race (to assess potential effect measure modification). Analyses with fewer than 20 events in either treatment strategy were omitted. The Benjamini– Hochberg procedure was applied separately within each family of tests, using the number of qualifying analyses as the number of tests.^28,29^ Adjusted p-values were rendered monotonic. An association survived false discovery rate control at q ≤ 0.05 only if its adjusted p-value was ≤ 0.05 and the subdistribution hazard ratio exceeded 1.0 (sHR > 1.0).

Sensitivity to residual confounding was assessed with three complementary quantitative bias analyses (QBAs), applied only to associations that survived false discovery rate control. The primary method, incorporated into the confirmation algorithm, was probabilistic QBA.^30,31^ For the probabilistic QBA, parameters were sampled 5,000 times from uniform distributions (parameters included: confounder–outcome risk ratio 1.25–3.00; prevalence difference 0.05– 0.25; baseline prevalence in strategy A2 = 0.15). These ranges were chosen to reflect realistic magnitudes of residual confounding that could plausibly persist after extensive measured covariate adjustment in large claims-based pharmacoepidemiologic studies of medication safety (i.e., moderate unmeasured confounders with risk ratios typically between 1.25 and 3.0 and prevalence differences of 5–25 %). The baseline prevalence of 0.15 was selected as a conservative mid-range value consistent with common chronic conditions in older adults. Each observed subdistribution hazard ratio was adjusted, yielding a distribution summarized by its median, 2.5th and 97.5th percentiles, and the proportion of draws remaining above 1.0. This procedure systematically introduces residual confounding of prespecified magnitude into the observed estimates to evaluate whether the associations remain elevated, a critical safeguard in confirmatory analyses where residual bias is a primary threat to validity. To complement the probabilistic QBA, a deterministic QBA examined a fixed grid of the same bias parameters, and E-values were calculated for every stratum specific association under a rare outcome assumption.^30–32^

A prespecified ADE association was confirmed only if it met both of the following criteria: (1) it survived within-outcome Benjamini–Hochberg false discovery rate control at q ≤ 0.05 with sHR > 1.0, and (2) under probabilistic QBA, both the median and the 2.5th percentile of the bias-adjusted subdistribution hazard ratios remained > 1.0. Requiring both the median and the lower percentile of the bias-adjusted distribution to exceed the null ensured that the association remained elevated after integrating over the full prespecified range of residual confounding, rather than relying solely on central tendency. This dual threshold rule prioritized specificity, which is appropriately conservative for confirmatory evaluation.

A post hoc sensitivity analysis examined potential effect measure modification by concomitant high risk antithrombotic therapy, defined as a dispensing of clopidogrel, ticagrelor, prasugrel, warfarin, rivaroxaban, apixaban, dabigatran, edoxaban, enoxaparin, or unfractionated heparin on each trial start date. The rationale for this analysis was grounded in published evidence linking warfarin (and vitamin K antagonists more broadly) to accelerated cardiac valve calcification^33,34^ and the well-established association of high risk antithrombotic agents with hemorrhagic stroke.^35^ Weighted Fine-Gray models that included the product term between atorvastatin initiation and high risk antithrombotic therapy were fit overall, within demographic subgroups, and within successive time windows. From each model the subdistribution hazard ratio for atorvastatin was obtained in the absence of high risk antithrombotic therapy (main effect term), in its presence (linear combination of main effect and product terms), and for the product term itself. Effect modification by high risk antithrombotic therapy was considered present when the product term was statistically significant (p < 0.05).

While some investigators prefer to restrict confirmation to the overall analysis, we elected to confirm associations that met the prespecified dual criteria in time stratified or subgroup analyses when the proportional hazards assumption was violated or when external evidence supported subgroup heterogeneity, thereby prioritizing validity of the effect estimate over a single summary measure. All analyses were conducted in Stata 18.0. The study was reported in accordance with the TARGET guideline (supplemental material **Table S10**).

## RESULTS

### Study Cohort

The study cohort comprised 70,130 statin naïve Medicare fee-for-service beneficiaries aged ≥ 65 years discharged after myocardial or cerebral infarction (see disposition table in supplemental material **Table S4**). During the 14-day trial initiation window, 39,948 initiated atorvastatin (strategy A1; 81% high-intensity, 40 or 80 mg), 19,182 initiated a different new outpatient medication (strategy A2), and 11,000 started no new oral medication (strategy A0). Concomitant high risk antithrombotic therapy, on each trial start date, was more common among atorvastatin initiators (53% A1 versus 44% A2). Outcome specific analyses excluded individuals who failed that trial’s eligibility criteria.

Mean follow-up before inverse probability of censoring weighting was substantially longer under strategy A1 (119 days) than under strategy A2 (52 days), reflecting higher rates of treatment deviation in the comparator arm. Inverse probability of censoring weights were applied to correct for this differential artificial censoring and to recover unbiased per protocol estimates.

The number of sequential trials per outcome ranged from 8 to 11 (mean 10.2). Outcomes with higher baseline prevalence or earlier occurrence after the index (time zero) event generated fewer trials, as a larger fraction of patients were excluded as prevalent cases and the minimum cell size requirement (at least 40 patients in each treatment arm) was reached sooner.

### Baseline Characteristics

Baseline characteristics of the Trial 0 cohort for the acute hemorrhagic cerebrovascular disease outcome (hemorrhagic stroke) are shown in **Table 2**. After exclusion of patients with a baseline bleeding event, the cohort comprised 24,773 individuals who initiated atorvastatin (A1) and 6,298 who initiated a different new medication (A2). Mean age was 76.1 years versus 76.3 years; 49.9% of patients in both arms were female. Racial distribution, dual eligibility, prior year healthcare utilization, Charlson Comorbidity Index, and key comorbidities (cardiac dysrhythmias, Type 2 diabetes, hypertension) were closely comparable. Baseline anticoagulant use and rates of percutaneous coronary intervention were nearly identical. All absolute standardized mean differences (SMDs) for the 573 covariates included in the weight generating model were well below 0.1. The randomly selected A2 medications used to classify discontinuation are presented in supplemental material **Table S2**; the distribution of A1 and A2 medications utilized during follow-up appears in supplemental material **Table S9**.

**Table 2.** Inverse Probability of Treatment Weighted (IPTW) Baseline Characteristics for the Acute hemorrhagic cerebrovascular disease outcome (Trial 0)

| Baseline Characteristics for Acute hemorrhagic cerebrovascular disease |  |  |  |
| --- | --- | --- | --- |
| Trial 0 | Treatment Strategy |  | SMD* |
|  | A1 - Atorvastatin | A2 - Other |  |
| Cohort (N) | 24,773 | 6,298 |  |
| **Demographics** | %/mean(SD)median | %/mean(SD)median |  |
| Age (years, floored) | 76.1 (8.1) 75.0 | 76.3 (8.3) 75.0 | 0.0237 |
| Age (Categorical) |  |  | 0.0119 |
| 65-74 | 49.2% | 48.8% |  |
| 75-84 | 33.1% | 33.1% |  |
| 85+ | 17.7% | 18.1% |  |
| Female sex | 49.9% | 49.9% | 0.0002 |
| Race/Ethnicity |  |  | 0.0075 |
| White | 85.0% | 84.9% |  |
| Black | 6.2% | 6.2% |  |
| Asian | 2.2% | 1.8% |  |
| Hispanic | 4.3% | 4.3% |  |
| **Characteristics upon 1st Medicare Enrollment** |  |  |  |
| Dual Medicare/Medicaid | 15.4% | 15.8% | 0.0121 |
| Disabled | 10.2% | 10.2% | 0.0004 |
| End stage renal disease | 1.0% | 1.0% | 0.0034 |
| **Healthcare Resource Utilization & Costs** |  |  |  |
| Inpatient admission (yes/no) | 11.8% | 12.1% | 0.0096 |
| # Inpatient admissions | 1.2 (0.5) 1.0 | 1.2 (0.5) 1.0 | 0.0109 |
| # Inpatient days | 5.1 (3.6) 4.0 | 5.2 (4.2) 4.0 | 0.0228 |
| ED visit (yes/no) | 39.8% | 40.1% | 0.0061 |
| # Emergency department visits | 0.6 (1.0) 0.0 | 0.6 (1.5) 0.0 | 0.0160 |
| # Unique days w/healthcare encounter | 19.9 (15.6) 16.0 | 20.1 (15.4) 16.0 | 0.0168 |
| Total healthcare costs (USD) | 21,091.2 (16,537.6) 16,655.9 | 21,311.1 (16,895.8) 16,482.4 | 0.0128 |
| **Baseline Comorbidities** |  |  |  |
| Charlson Comorbidity Index (continuous) | 5.6 (2.6) 5.0 | 5.7 (2.6) 5.0 | 0.0300 |
| Essential hypertension | 80.5% | 81.6% | 0.0285 |
| Acute myocardial infarction | 67.3% | 66.6% | 0.0143 |
| Disorders of lipid metabolism | 66.8% | 67.1% | 0.0065 |
| Hypertension with complications and secondary hypertension | 36.5% | 37.3% | 0.0148 |
| Nervous system signs and symptoms | 36.3% | 37.3% | 0.0205 |
| Cardiac dysrhythmias | 33.9% | 33.9% | 0.0008 |
| Diabetes mellitus, Type 2 | 27.7% | 28.3% | 0.0113 |
| Heart failure | 24.8% | 24.8% | 0.0006 |
| **Baseline Pharmacotherapy Utilization** |  |  |  |
| Central Nervous System Agents | 49.6% | 50.7% | 0.0220 |
| Hormones and Synthetic Substitutes | 46.8% | 48.1% | 0.0274 |
| Anti-infective Agents | 40.6% | 41.8% | 0.0253 |
| Gastrointestinal Drugs | 35.3% | 36.2% | 0.0189 |
| Beta Adrenergic Blocking Agents | 28.2% | 29.2% | 0.0211 |
| Adrenals | 25.1% | 25.7% | 0.0146 |
| Selective Beta Adrenergic Blocking Agent | 22.2% | 23.0% | 0.0206 |
| Corticosteroids | 21.5% | 22.2% | 0.0160 |
| Class II Antiarrhythmics | 20.5% | 21.2% | 0.0169 |
| Angiotensin Converting Enzyme Inhibitors | 20.2% | 20.5% | 0.0091 |
| Blood Formation, Coagulation, and Thrombosis | 13.1% | 13.4% | 0.0074 |
| **Baseline Procedures** |  |  |  |
| Electrocardiogram | 94.2% | 94.6% | 0.0186 |
| Diagnostic ultrasound of heart (echocardiogram) | 89.5% | 89.8% | 0.0096 |
| Diagnostic cardiac catheterization, coronary arteriography | 56.8% | 56.4% | 0.0069 |
| Cardiac and coronary fluoroscopy | 55.8% | 55.3% | 0.0105 |
| Percutaneous coronary interventions (PCI) | 44.0% | 42.9% | 0.0231 |
| Percutaneous transluminal coronary angioplasty (PTCA) | 43.4% | 41.7% | 0.0348 |
| Computerized axial tomography (CT) scan head | 42.7% | 43.5% | 0.0162 |

Notes: A1 = new initiation of atorvastatin; A2 = new initiation of a different medication other than atorvastatin. SMD\* = absolute value of the standardized mean difference (A1 vs A2). All demographic, comorbidity, healthcare utilization, pharmacotherapy, and procedure variables were assessed in the 12 months prior to time zero (T0, the index date). ED = emergency department; PCI = percutaneous coronary intervention; PTCA = percutaneous transluminal coronary angioplasty; CT = computed tomography. Values are presented as percentages or mean (SD) median as indicated. This table shows baseline characteristics for the Trial 0 analytic cohort of the acute hemorrhagic cerebrovascular disease outcome only.

Covariate balance after inverse probability of treatment weighting for each trial is summarized in **Table 3** and supplemental material **Table S6**. In Trial 0, the mean SMD of 573 baseline covariates included in the weight generating model was 0.009 (maximum 0.037); in Trial 1 (335 baseline covariates), the corresponding values were 0.019 and 0.095. Both early trials remained below the conventional threshold of 0.1. Beginning with Trial 2, the maximum SMD exceeded 0.1. The larger sample sizes in Trials 0 and 1 permitted inclusion of a greater number of covariates and produced superior covariate balance, providing the rationale for the sensitivity analysis restricted to these two trials.

**Table 3.** Covariate Balance Across Sequential Trials: Acute hemorrhagic cerebrovascular disease.

| **Covariate Balance After Inverse-Probability-of-Treatment Weighting Across Sequential Trials of Atorvastatin Initiation Versus Alternative Medication Initiation** |  |  |  |  |  |  |  |
| --- | --- | --- | --- | --- | --- | --- | --- |
| **Outcome: *Acute hemorrhagic cerebrovascular disease*\*** |  |  |  |  |  |  |  |
| **Trials** | **N** | **N (A1)** | **N (A2)** | **No. of covariates** | **Mean SMD** | **Max SMD** | **Max SMD < 0.1** |
| **0** | **31071** | **24773** | **6298** | **573** | **0.009** | **0.037** | **Yes** |
| **1** | **3155** | **2250** | **905** | **335** | **0.019** | **0.095** | **Yes** |
| 2 | 576 | 338 | 238 | 174 | 0.046 | 0.148 | No |
| 3 | 284 | 134 | 150 | 104 | 0.103 | 0.311 | No |
| 4 | 194 | 95 | 99 | 72 | 0.082 | 0.269 | No |
| 5 | 154 | 60 | 94 | 53 | 0.147 | 0.414 | No |
| 6 | 177 | 59 | 118 | 47 | 0.104 | 0.550 | No |
| 7 | 147 | 54 | 93 | 42 | 0.094 | 0.346 | No |

Notes: Trials = sequential trial number (0–7); N = total number of eligible participants in the trial; N(A1) = number of participants initiating atorvastatin (A1); N(A2) = number of participants initiating a different medication (A2); No. of covariates = number of covariates included in the inverse-probability-of-treatment weighting (IPTW) model for that trial; Mean SMD = mean absolute standardized mean difference across all covariates after IPTW; Max SMD = maximum absolute standardized mean difference across all covariates after IPTW; Max SMD < 0.1 = indicator of whether the maximum SMD is below the conventional threshold of 0.1 for adequate covariate balance (Yes/No). A1 = atorvastatin initiation; A2 = initiation of a different medication.

### Confirmation of Previously Detected ADE Signals

Several associations identified in the earlier high dimensional, hypothesis free signal detection study met the prespecified dual criteria for confirmation: survival of within outcome false discovery rate control at q ≤ 0.05 with a subdistribution hazard ratio greater than 1.0, and retention of an elevated association after probabilistic QBA. Summary results appear in **Table 4**; detailed estimates are presented in **Table 5** and **Figures 2A–2M** and supplemental material **Table S7**. The full results of the post hoc analysis examining potential effect measure modification by concomitant high risk antithrombotic therapy are presented in supplemental material **Table S11**.

**Table 4.**
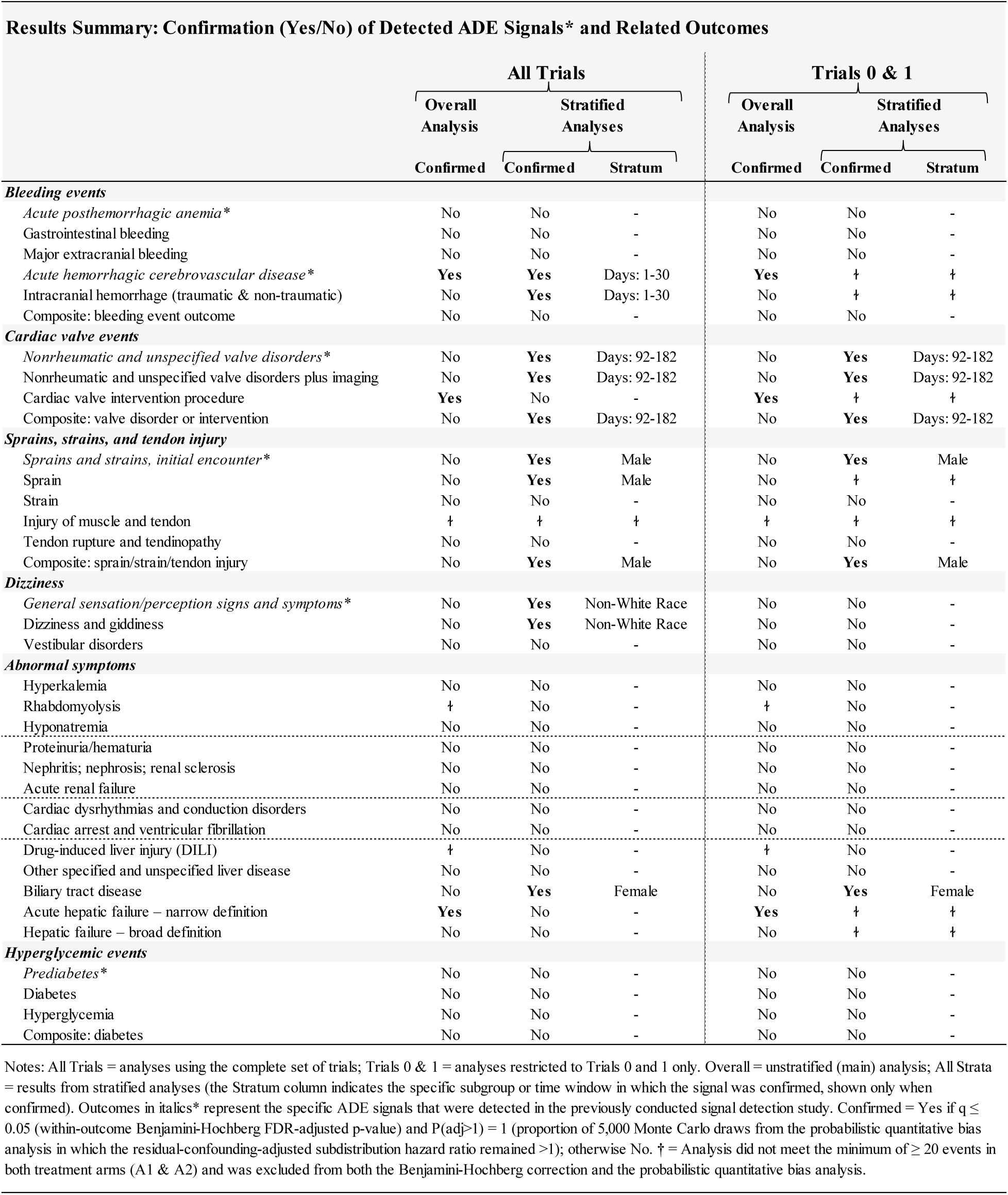
Results Summary: Confirmation (Yes/No) of Detected ADE Signals* and Related Outcomes.

**Table 5.**
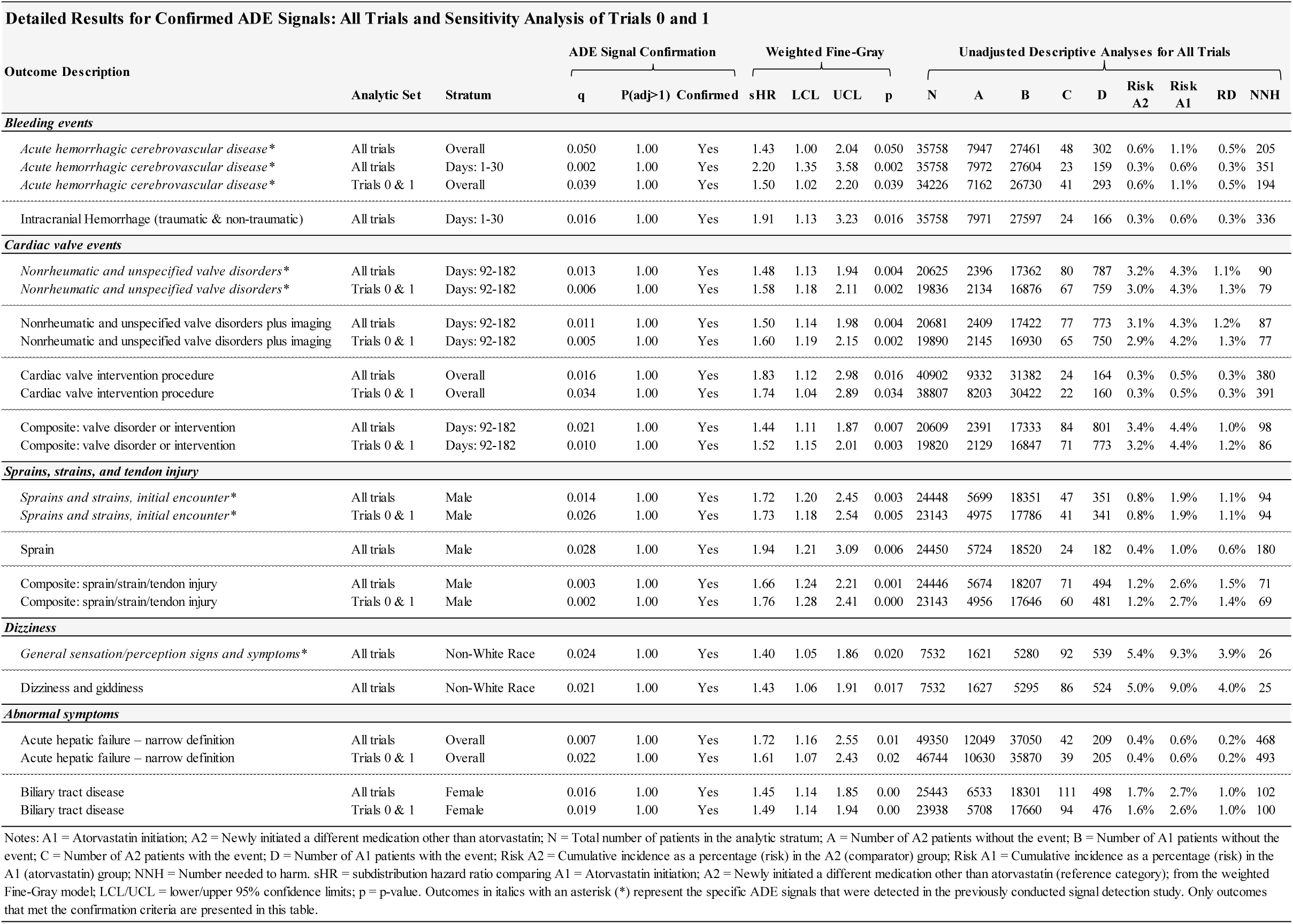
Detailed Results for Confirmed ADE Signals: All Trials and Sensitivity Analysis of Trials 0 and 1.

**Figure 2A.**
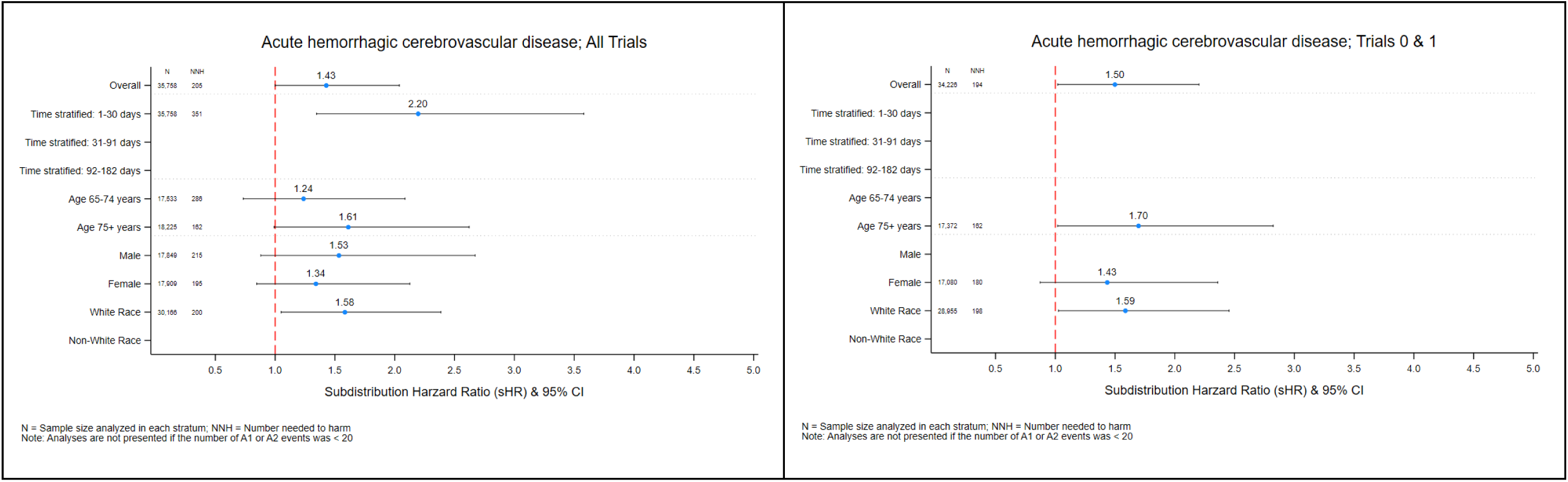
Subdistribution Hazard Ratios for the Confirmed ADE Signal (All Trials and Trials 0 & 1) Outcome: Acute hemorrhagic cerebrovascular disease

**Figure 2B.**
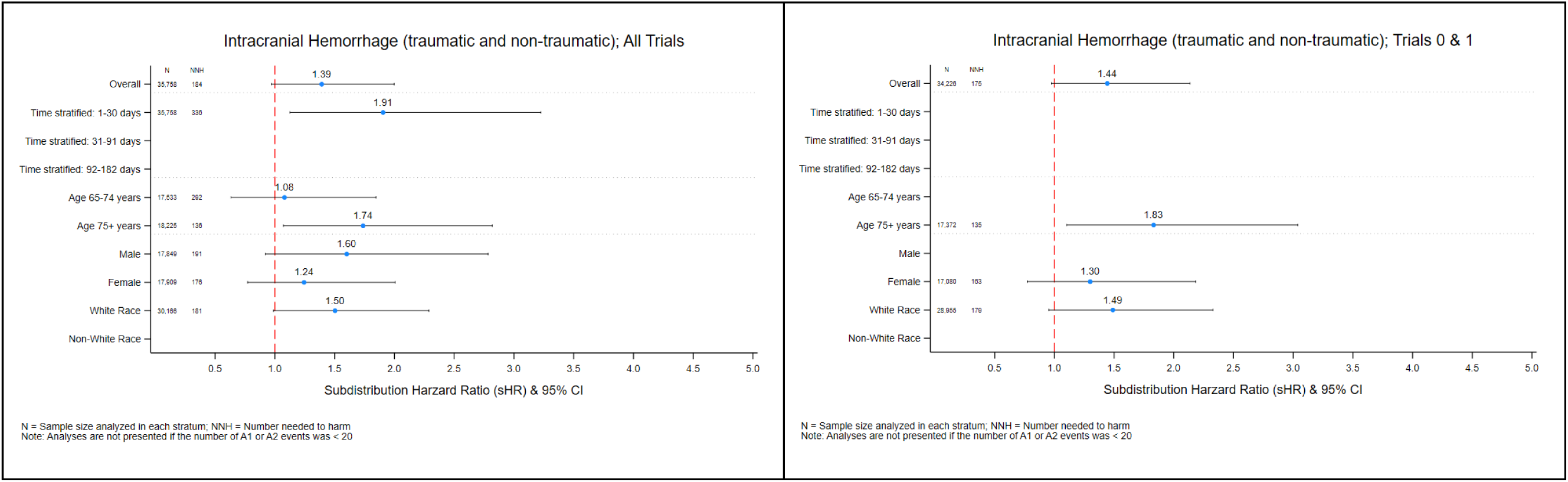
Subdistribution Hazard Ratios for the Confirmed ADE Signal (All Trials and Trials 0 & 1) Outcome: Intracranial Hemorrhage (traumatic & non-traumatic)

**Figure 2C.**
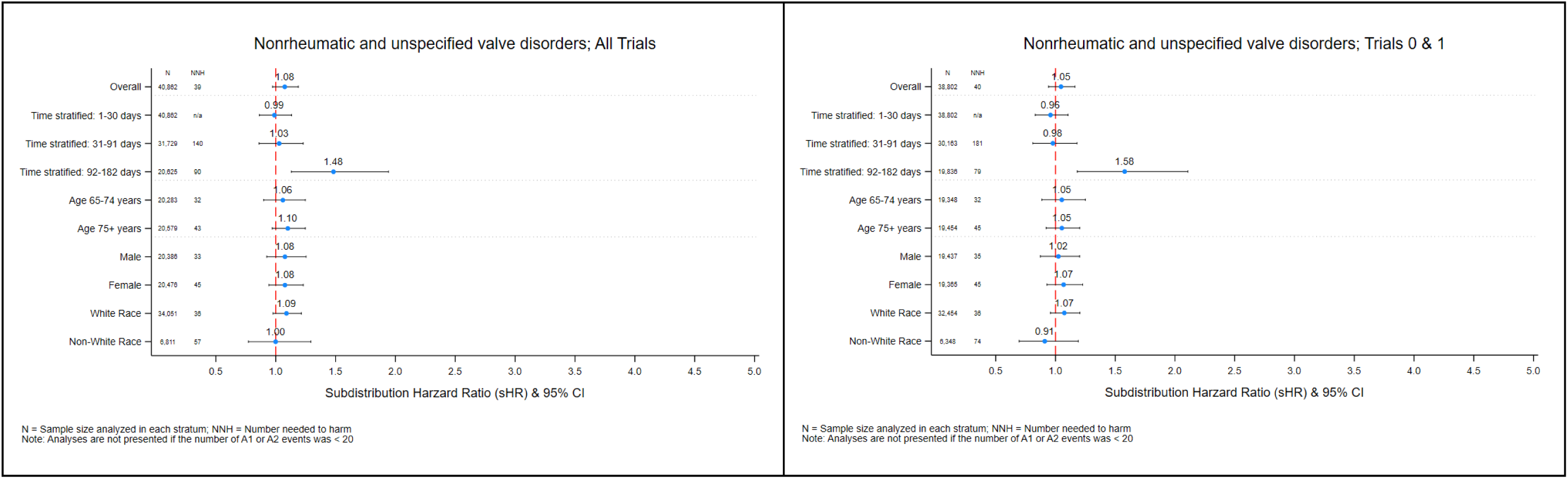
Subdistribution Hazard Ratios for the Confirmed ADE Signal (All Trials and Trials 0 & 1) Outcome: Nonrheumatic and unspecified valve disorders

**Figure 2D.**
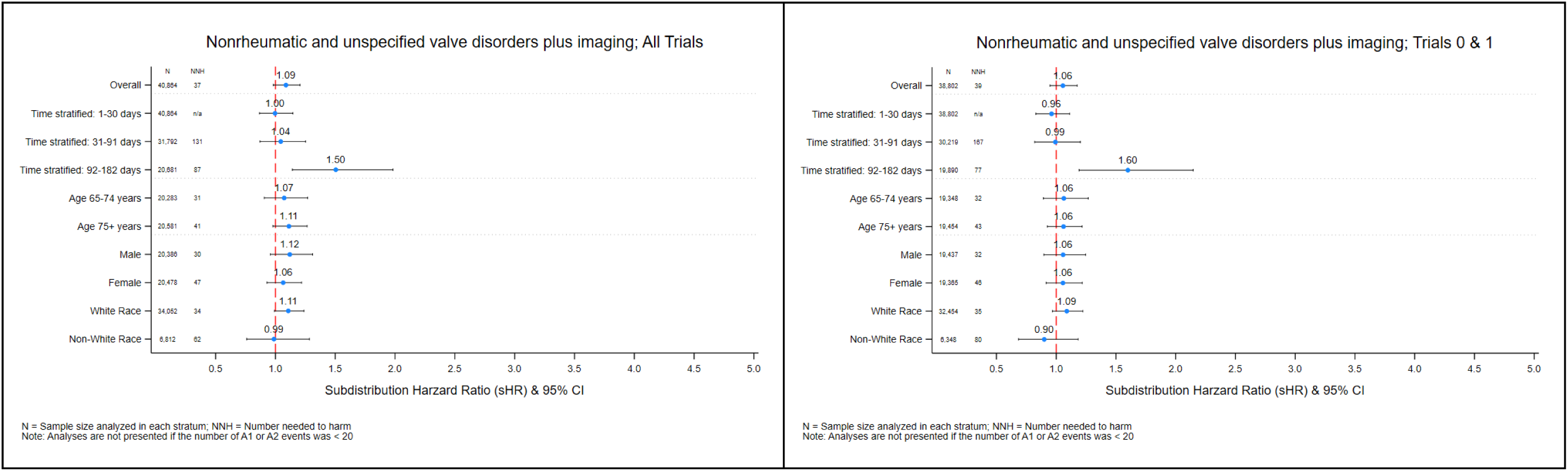
Subdistribution Hazard Ratios for the Confirmed ADE Signal (All Trials and Trials 0 & 1) Outcome: Nonrheumatic and unspecified valve disorders plus imaging

**Figure 2E.**
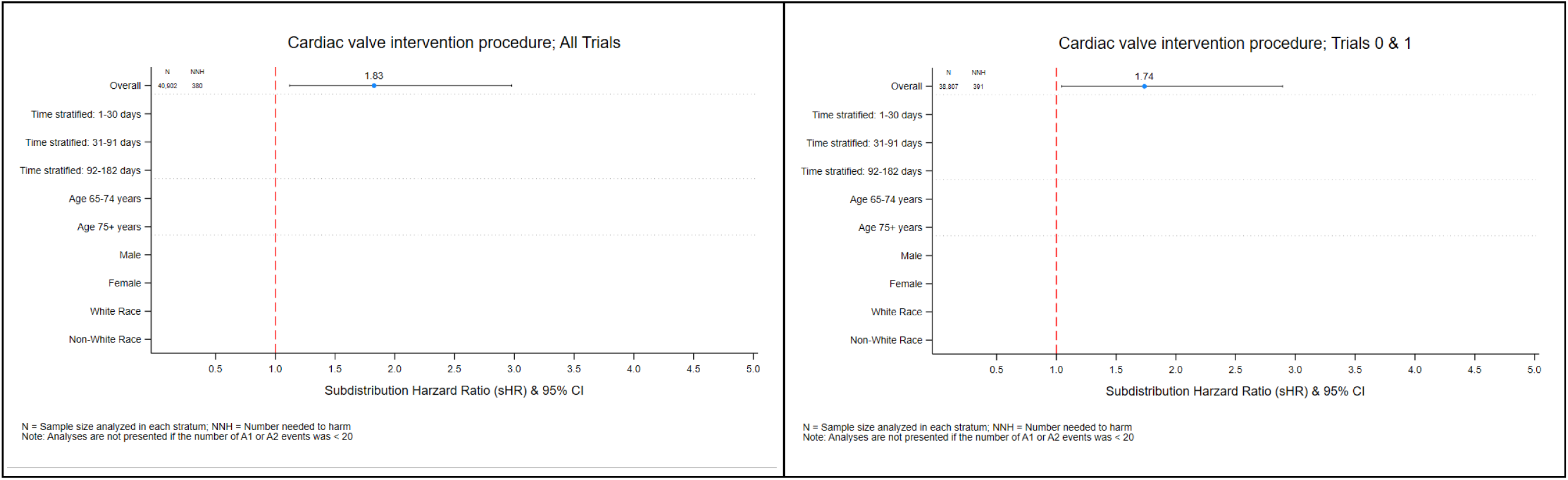
Subdistribution Hazard Ratios for the Confirmed ADE Signal (All Trials and Trials 0 & 1) Outcome: Cardiac valve intervention procedure

**Figure 2F.**
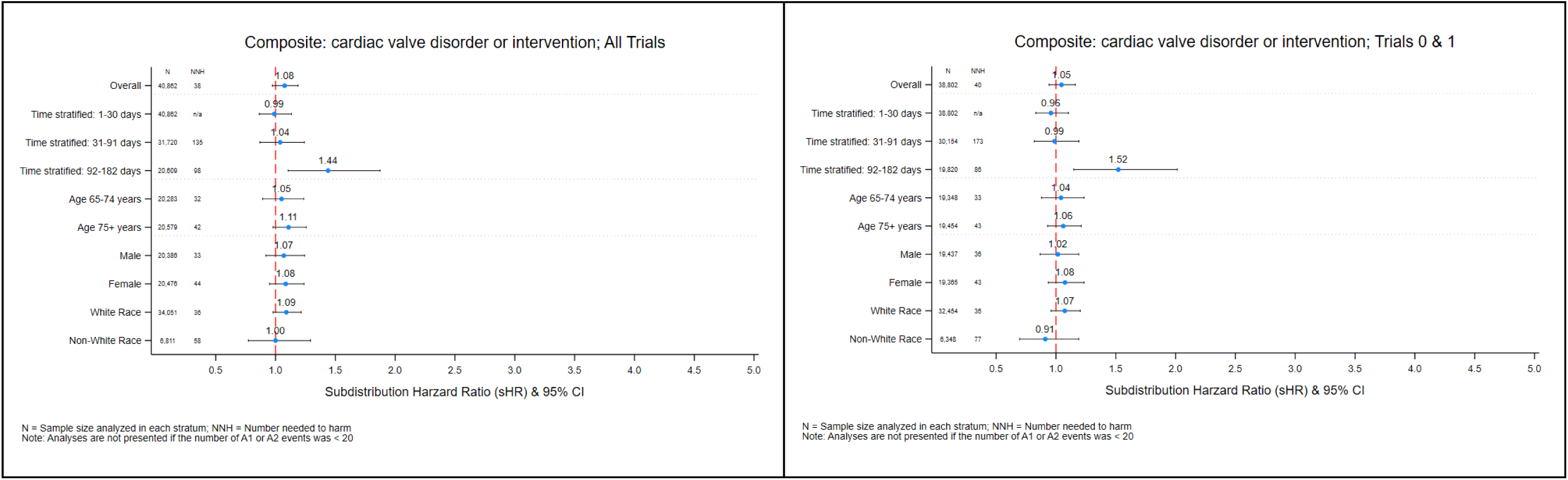
Subdistribution Hazard Ratios for the Confirmed ADE Signal (All Trials and Trials 0 & 1) Outcome: Valve disorder or intervention (composite outcome)

**Figure 2G.**
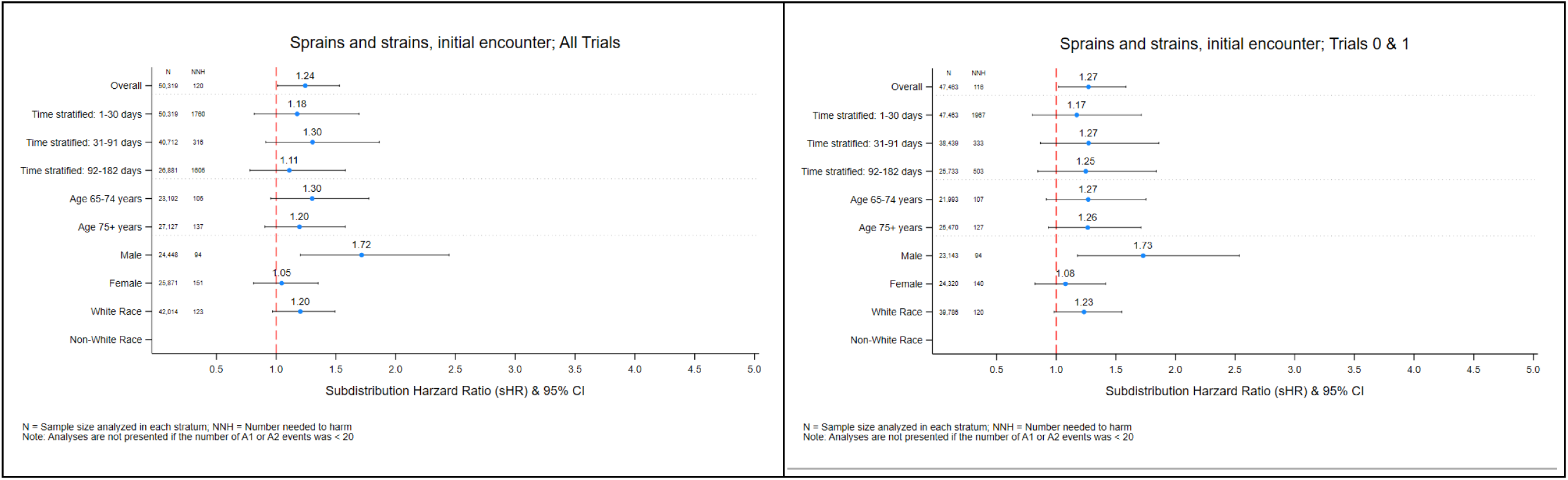
Subdistribution Hazard Ratios for the Confirmed ADE Signal (All Trials and Trials 0 & 1) Outcome: Sprains and strains, initial encounter

**Figure 2H.**
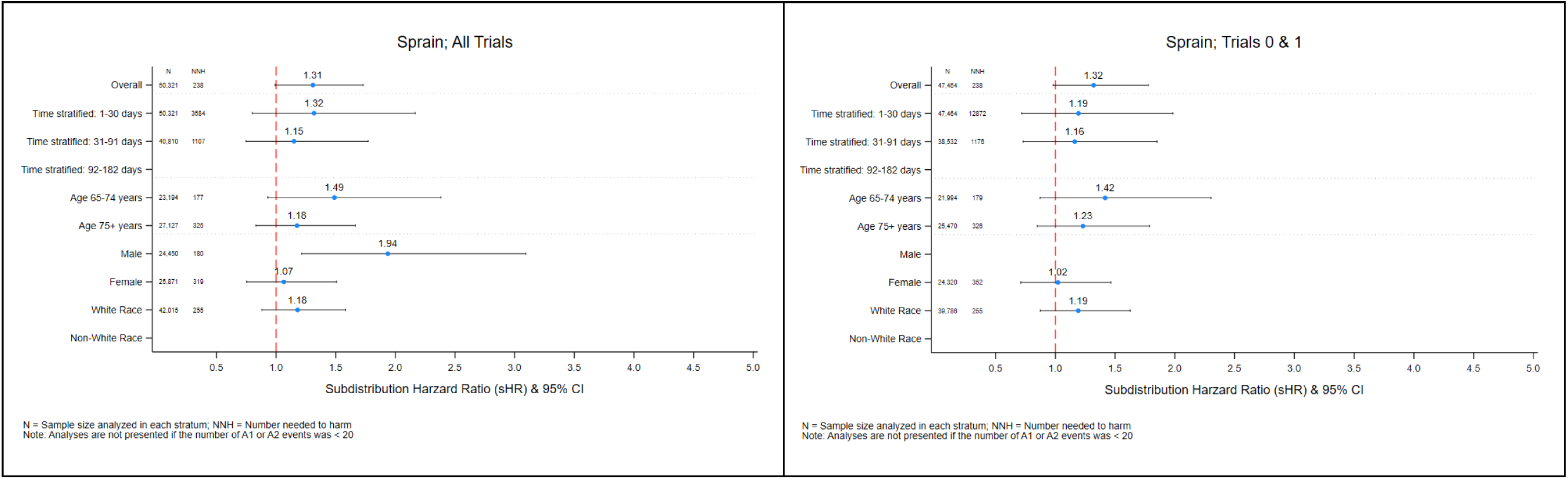
Subdistribution Hazard Ratios for the Confirmed ADE Signal (All Trials and Trials 0 & 1) Outcome: Sprains

**Figure 2I.**
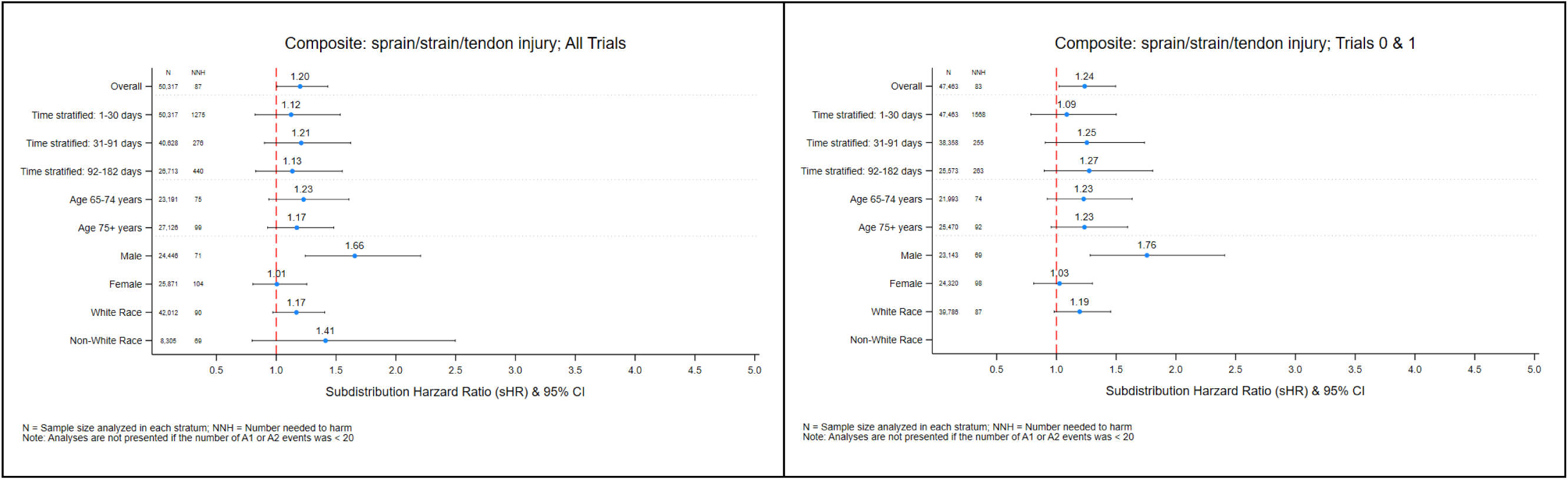
Subdistribution Hazard Ratios for the Confirmed ADE Signal (All Trials and Trials 0 & 1) Outcome: Sprain/strain/tendon injury (composite outcome)

**Figure 2J.**
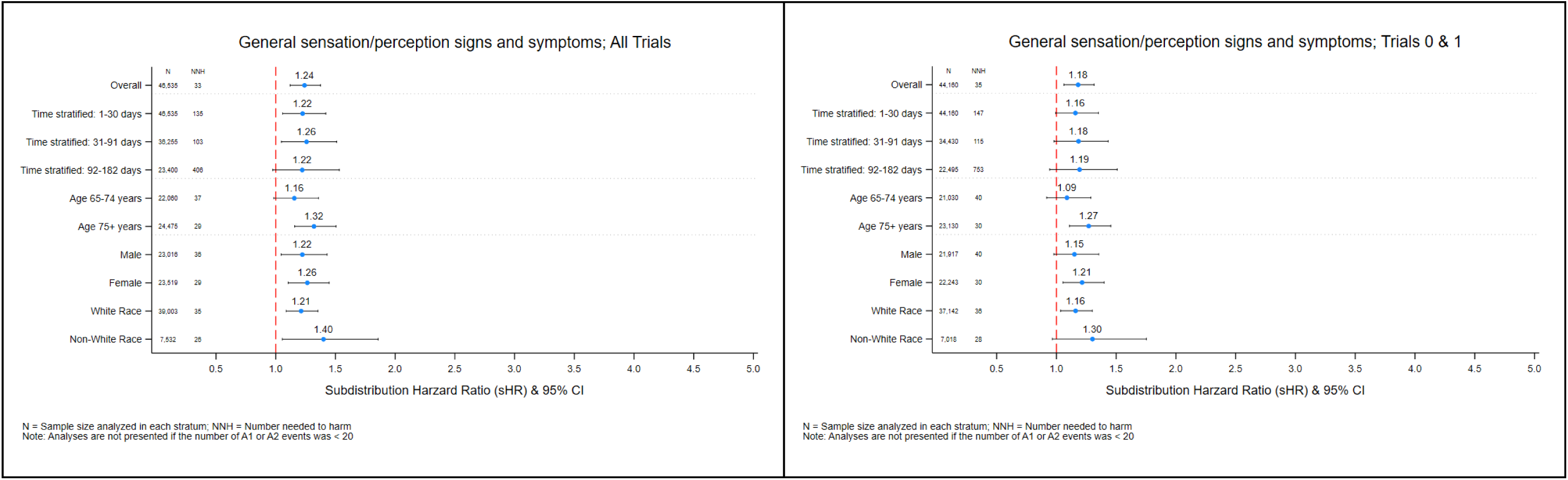
Subdistribution Hazard Ratios for the Confirmed ADE Signal (All Trials and Trials 0 & 1) Outcome: General sensation/perception signs and symptoms

**Figure 2K.**
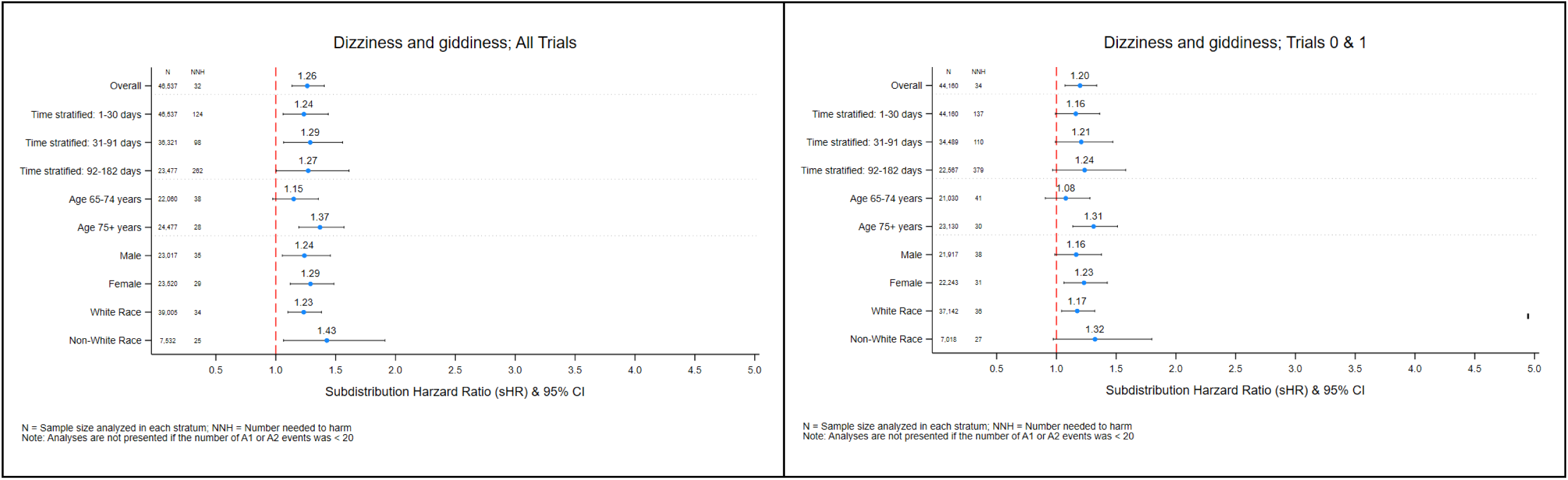
Subdistribution Hazard Ratios for the Confirmed ADE Signal (All Trials and Trials 0 & 1) Outcome: Dizziness and giddiness

**Figure 2L.**
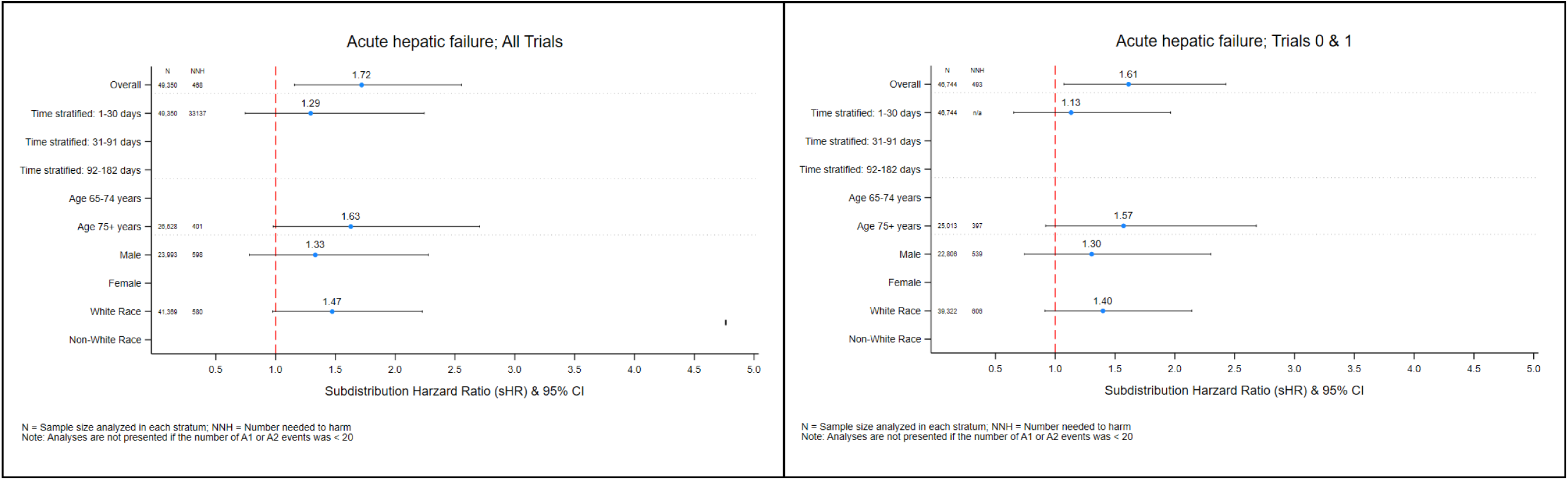
Subdistribution Hazard Ratios for the Confirmed ADE Signal (All Trials and Trials 0 & 1) Outcome: Acute hepatic failure - narrow definition

**Figure 2M.**
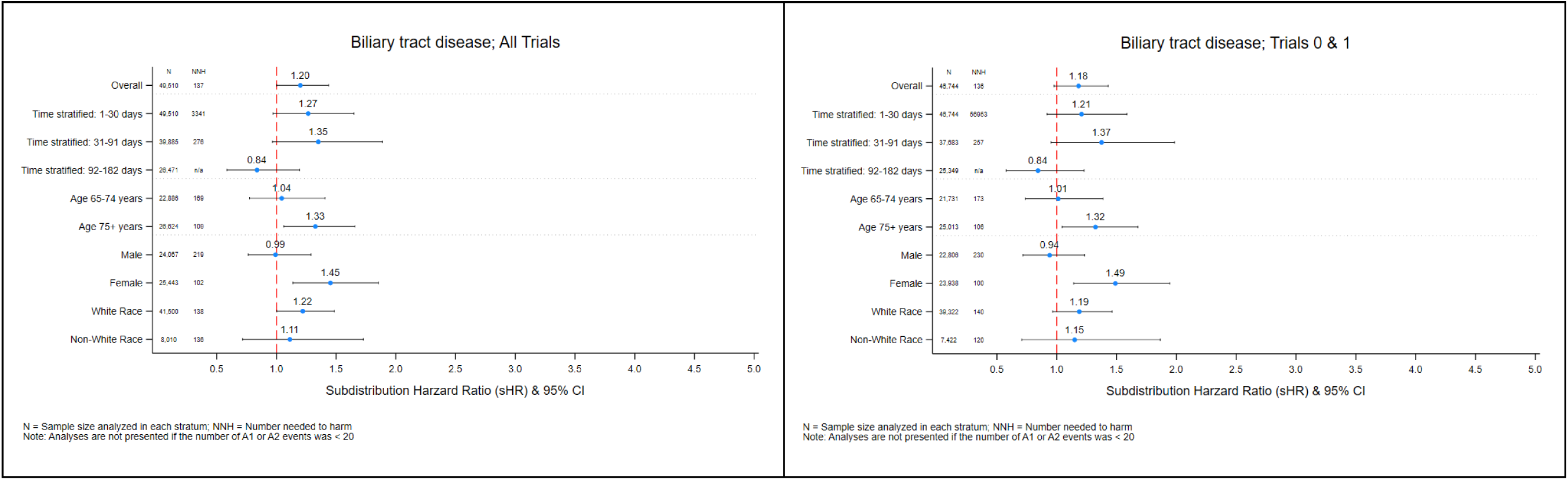
Subdistribution Hazard Ratios for the Confirmed ADE Signal (All Trials and Trials 0 & 1) Outcome: Biliary tract disease

At the highest level of confirmatory support were the early excess risks of acute hemorrhagic cerebrovascular disease and acute hepatic failure. Acute hemorrhagic cerebrovascular disease showed a modest overall association (sHR 1.43, 95% CI 1.00–2.04; NNH 205), corresponding to a 43% higher hazard among patients initiating atorvastatin than among those initiating a different medication, or one excess event per 205 atorvastatin treated patients. This association was substantially stronger in the first 30 days (sHR 2.20, 1.35–3.58; NNH 351) and remained elevated when restricted to Trials 0 and 1 with optimal confounding control (sHR 1.50, 1.02– 2.20; NNH 194). Intracranial hemorrhage (traumatic and non-traumatic) exhibited a similar early elevation (sHR 1.91, 1.13–3.23; NNH 336). Acute hepatic failure (narrow definition) was confirmed overall (sHR 1.72, 1.16–2.55; NNH 468) and persisted in Trials 0 and 1 (sHR 1.61, 1.07–2.43; NNH 493). Both the hemorrhagic and hepatic associations remained robust in the absence of concomitant high risk antithrombotic therapy, indicating that the observed elevations were not explained by concurrent use of these agents.

At the intermediate level of confirmatory support were biliary tract disease and musculoskeletal injuries. Biliary tract disease was confirmed among women (sHR 1.45, 1.14–1.85; NNH 102) with consistent estimates under optimal covariate balance (i.e., Trials 0 and 1). Musculoskeletal injuries (sprains and strains, specific sprain, and the composite of sprain, strain, or tendon injury) were confirmed among men (sHR 1.66–1.94; NNH 69–180) and remained robust in the restricted trial set with optimal covariate balance. Neither association was meaningfully modified by high risk antithrombotic therapy.

At the weaker end of confirmatory support were cardiac valve disorders and interventions, together with sensory symptoms. Nonrheumatic valve disorders (with or without imaging) and the composite of valve disorder or intervention were confirmed in days 92–182 (sHR 1.44–1.60; NNH 77–98). Cardiac valve intervention procedures were confirmed overall (sHR 1.83, 1.12– 2.98; NNH 380). However, the association between atorvastatin and valve disorders was substantially attenuated in the absence of high risk antithrombotic therapy, reducing its confirmatory weight and suggesting that the observed elevation may have been driven by concurrent high risk antithrombotic therapy rather than by atorvastatin itself. Sensory symptoms (general sensation or perception abnormalities and dizziness) were confirmed only among non-White patients and only in the unrestricted set of trials (sHR 1.40–1.43); neither association persisted under the superior covariate balance of Trials 0 and 1.

Two previously detected signals failed the dual confirmation criteria, including acute posthemorrhagic anemia and prediabetes. In these instances, the associations either did not survive within outcome false discovery rate control or proved sensitive to residual confounding. The results presented here reflect only those outcomes and specific analyses that fully satisfied the dual confirmation criteria. Under probabilistic QBA, every confirmed association retained a median bias adjusted subdistribution hazard ratio greater than 1.0 and an empirical lower tail entirely above the null, indicating robustness to residual confounding of the magnitude considered *a priori*. Parallel deterministic QBA and E value calculations yielded consistent findings (supplemental material **Table S8**). Residual confounding of the magnitude evaluated is therefore unlikely to account fully for the confirmed associations. Additional subgroup analyses, for the confirmed outcomes, that reached nominal statistical significance but did not survive multiplicity correction or probabilistic QBA are presented in **Figures 2A–2M** and supplemental material **Table S7**.

## DISCUSSION

This internal confirmatory sequential target trial emulation estimated the per-protocol effect of initiating atorvastatin versus a different prescription medication in the post-myocardial/cerebral infarction setting on a series of outcomes identified from a previous ADE (signal) detection study. Associations that met the dual confirmatory criteria varied in terms of magnitude of associations and robustness to quantitative bias analysis. The strongest confirmatory support was observed for early acute hemorrhagic cerebrovascular disease and for acute hepatic failure. Intermediate support was observed for biliary tract disease and musculoskeletal injuries. Weaker and more conditional evidence was observed for cardiac valve disorders and for sensory symptoms. Prediabetes and acute posthemorrhagic anemia failed to meet the dual confirmatory criteria. This study, together with its antecedent ADE detection counterpart, represents a novel systematic approach to proactive, claims-based pharmacovigilance: a high dimensional, hypothesis free sequence of independent target trial emulations for hundreds of outcomes with signal detection thresholds favoring sensitivity, followed by prespecified internal confirmatory trial emulations that retain the same time zero and source population but tighten eligibility and exclusions, replace heterogeneous outcomes with those that are more homogeneous within the same family, control multiplicity more strictly at the level of each hypothesis, and require that residual confounding of prespecified magnitude not nullify the association.

Of the two associations atop the confirmation gradient, acute hemorrhagic cerebrovascular disease is the more compelling. Intracranial bleeding is comparatively rare but often devastating. In older adults hospitalized for myocardial or cerebral infarction, the greatest contributors are ostensibly underlying vascular disease and use of high risk antithrombotic agents. However, the association between atorvastatin use and acute hemorrhagic cerebrovascular disease cannot be explained away on these bases. For one, the association remained detectable despite extensive adjustment for these as well as many other factors. Furthermore, the association between atorvastatin use and intracerebral hemorrhage was present even in the subgroup of patients not exposed to high-risk antithrombotic therapy. Additionally, the association was robust to various outcome definitions, including when the outcome was reassigned according to the Sentinel definition of intracranial hemorrhage. Of note, a similar hemorrhagic stroke excess was observed in the SPARCL trial, in which patients with recent stroke or TIA were randomized to high intensity atorvastatin (80 mg daily) versus placebo.^36^ Although the present study did not require a particular atorvastatin intensity, it was the case that 81% of atorvastatin initiators received 40 mg or 80 mg. Proposed mechanisms for an atorvastatin effect on intracranial hemorrhage include depletion of cholesterol in cerebral small vessels and pleiotropic effects of statins on platelet function and fibrinolysis,^37–40^ but such mechanisms have not been proven in older, post-infarction populations. Concentration of the excess risk in the first month after initiation makes accumulating surveillance an unlikely explanation for the finding. The relative increase was large (sHR 1.43–2.2) and the absolute excess small (NNH 194–351), the expected profile of a rare, serious event. That excess has to be formally weighed against the established benefit of high intensity statin therapy after infarction (NNT 28–93),^36,41–43^ a balance that may narrow among patients with advanced age, multimorbidity, polypharmacy, and competing risks.

Acute hepatic failure was the other association with strong confirmatory evidence. The signal detection study identified a heterogeneous class of abnormal laboratory findings, including abnormal results of liver function studies (ICD-10-CM R94.5). The present study replaced that class with a narrower definition of acute hepatic failure, among other hepatic dysfunction outcomes, and found that atorvastatin initiation was associated with this acute hepatic failure in the full set of trials and in the earliest trials, where measured confounding was most tightly controlled (**Figure 2L** and supplemental material **Table S7**). Despite a small absolute excess risk (NNH 468–493), this association is clinically important because acute hepatic failure is a serious outcome and the result is consistent with the recognized idiosyncratic hepatotoxicity of statins, including atorvastatin.^44,45^

Adverse associations at an intermediate position on the evidence gradient were of atorvastatin initiation with biliary tract disease and with musculoskeletal injury. On gender-stratified analyses, the confirmation signal for biliary tract disease formally met criteria only among women and that for sprains, strains, and tendon injury only among men. Overall estimates (i.e., for both genders together) were of similar magnitude, but failed to achieve the statistical significance threshold for formal confirmation. At face value, these patterns could reflect effect measure modification (i.e., implying gender-specific effect of atorvastatin). However, we find it more likely that this is a statistical artifact: e.g., higher underlying disease frequency of biliary tract disease among women led to higher event rates, greater statistical precision and thereby attainment of statistical confirmatory criteria in this subgroup but not among men (the latter despite a similar effect estimate). A similar statistical artifact could impact musculoskeletal injury in favor of men.^46–49^

At the weaker end of the gradient were cardiac valve disorders and interventions, together with sensory symptoms (primarily dizziness). Nonrheumatic valve disorders and the composite of valve disorder or intervention were confirmed in the later follow-up window, and valve intervention procedures were confirmed overall. However, the association between atorvastatin and valve disorders (classified by diagnosis codes) was present and strong among users of concomitant high risk antithrombotic therapy, but attenuated and insignificant among nonusers, suggesting effect measure modification. To our knowledge, ours is the first study to suggest potential interaction between atorvastatin and antithrombotic drugs in this regard. Further studies are needed to confirm and, if consistent, elucidate the underlying mechanism. Sensory symptoms were confirmed only among non-White patients and only in the unrestricted trial set; they did not persist under superior covariate balance (in Trial 0 and Trial 1) and therefore occupy a more tentative position on the gradient of support.

Several previously detected ADE signals, including prediabetes and acute posthemorrhagic anemia, failed the dual confirmation criteria. Their attenuation after application of more refined outcome definitions, expanded exclusion criteria, stricter multiplicity control, and formal bias analysis underscores the specificity of the confirmatory procedure.

Strengths of this internal confirmation study derive from the sequential target trial architecture, inverse probability weighting that recovered per-protocol contrasts while treating death as a competing risk, hybrid outcome definitions that preserved continuity with the original CCSR categories while incorporating refined or alternative published outcome algorithms, within-outcome false discovery rate control, and a dual confirmation rule that required both statistical significance under multiplicity correction and robustness to residual confounding. Absolute effect measures (i.e., cumulative incidence and NNH) supplied clinical context that relative measures alone cannot provide. Stratified analyses proved particularly informative: they revealed clear violations of the proportional hazards assumption for several outcomes (most notably the concentration of cerebral hemorrhagic risk in the first 30 days) and illuminated clinically important effect measure modification (musculoskeletal injuries in men and biliary tract disease in women). The prespecified sensitivity analysis restricted to Trials 0 and 1 achieved superior covariate balance and demonstrated that all associations with strong or intermediate support on the gradient remained consistent in both the full set of trials and the optimally balanced early trials. The post hoc analysis of concomitant high risk antithrombotic therapy further clarified interpretation by identifying important effect measure modification for diagnostic valve disorders while confirming that the early hemorrhagic stroke associations were robust in the presence or absence of these agents. With the exception of the post hoc high risk antithrombotic therapy analysis, all analytic decisions adhered to a prespecified protocol, with no post hoc alterations to eligibility, treatment definitions, outcome algorithms, or confirmation thresholds.

Several limitations warrant careful consideration. Despite extensive measured covariate adjustment and formal QBA, residual confounding by unmeasured factors—including low density lipoprotein cholesterol levels, body mass index, frailty, physical activity, dietary patterns, and the precise severity of the index infarction—cannot be excluded. This limitation was offset by ascertaining and adjusting for a comprehensive set of baseline characteristics available in the Medicare claims data that span sociodemographic attributes, healthcare utilization and expenditures, an expansive range of comorbidities and pharmacotherapeutic classes, and diagnostic and interventional procedures, which serve as proxies for substantial portions of the clinical constructs underlying the unmeasured factors.

Claims based outcome ascertainment remains subject to misclassification, unrestricted diagnosis position coding, and potential differential surveillance, particularly for endpoints that rely on imaging or laboratory findings that may be ordered more frequently in patients receiving intensive secondary prevention therapy. Even after disaggregation, certain CCSR categories retain residual clinical heterogeneity. To address this concern, multiple operational definitions were examined within each outcome family; consistency of findings across alternative specifications for hemorrhagic stroke, musculoskeletal injuries, and hepatic dysfunction outcomes strengthens the evidence that the confirmed associations are not artifacts of a single coding algorithm or definition.

The active comparator (treatment strategy A2) comprised a heterogeneous set of newly initiated medications following myocardial or cerebral infarction. As expected, the agents most frequently initiated in both arms were cardiovascular therapies, including clopidogrel, nitroglycerin, beta blockers, and ACE inhibitors. Nonetheless, not every A2 medication was a cardiovascular drug; some patients entered the comparator arm after starting agents such as proton pump inhibitors. Even so, that difference does not imply that patients initiating atorvastatin were sicker or fundamentally different from patients initiating a different medication. On the contrary, patients included in strategies A1 and A2 were nearly exchangeable on measured baseline factors (**Table 2**): age, sex, Charlson comorbidity, prior myocardial infarction, heart failure, cardiac catheterization, and percutaneous coronary intervention all differed by standardized mean differences well below 0.10 (maximum 0.035). In parallel, baseline anticoagulant use was nearly identical (13.1% vs. 13.4%; standardized mean difference 0.0074). This degree of balance argues against material confounding by indication from the composition of A2. In keeping with that baseline similarity, pharmacotherapy utilization during the six months of follow-up (supplemental material **Tables S2** and **S9)** differed only modestly, including apixaban (9.5% vs. 10.5%), amiodarone (6.4% vs. 7.0%), and pantoprazole (11.6% vs. 11.5%). A separate design feature is that strategy A2 could include other statins (rosuvastatin, simvastatin, and pravastatin). Even then, follow-up in both arms was censored at initiation of any statin other than continued atorvastatin among patients already classified as A1. Consequently, if that inclusion induced any bias, the bias would attenuate rather than inflate associations and would therefore be appropriately conservative in a signal confirmation study.

Although concomitant high risk antithrombotic therapy was somewhat more common among atorvastatin initiators during follow-up (e.g., clopidogrel: 46.4% vs 37.9%), a post hoc analysis evaluated potential effect measure modification by high risk antithrombotic therapy. This important sensitivity analysis showed a strong association between initiating atorvastatin, compared with initiating a different medication, and incident cardiac valve disorders among patients who received concomitant high risk antithrombotic therapy (sHR 1.97; 95% CI 1.32 to 2.94). However, among patients who did not receive concomitant high risk antithrombotic therapy, the corresponding association (A1 vs A2) was compatible with no association (sHR 1.06; 95% CI 0.73 to 1.53). That pattern illuminates potential effect measure modification (interaction sHR 1.86; 95% CI 1.08 to 3.21), including a possible joint effect of atorvastatin and high risk antithrombotic therapy on cardiac valve disorders. It is not compatible with confounding by high risk antithrombotic therapy, which would have produced a null association between initiating atorvastatin, compared with initiating a different medication, and cardiac valve disorders among patients who all received concomitant high risk antithrombotic therapy. In contrast, the confirmed associations of atorvastatin initiation with hemorrhagic cerebrovascular disease and with acute hepatic failure remained elevated both in the presence and in the absence of high risk antithrombotic therapy, a pattern indicating that these agents neither confounded nor modified those associations. Taken together, the observed patterns of medication use and the stratified results reduce concern that differential concomitant pharmacotherapy is a major source of residual confounding for the confirmed outcomes.

The probabilistic QBA was dependent on the *a priori* range of bias parameters examined (confounder–outcome risk ratios 1.25–3.00; prevalence differences 0.05–0.25). Although confounding outside this range could in principle nullify some associations, the complete robustness observed within these realistic bounds strengthens confidence in the confirmed findings. Follow-up was limited to 182 days, precluding evaluation of longer term risks; however, most statin-related adverse events occur within the first few months after initiation,^45,47,50^ and the average duration of continuous statin use before discontinuation is approximately six months.^49,51^

The study population was restricted to fee-for-service Medicare beneficiaries aged 65 years or older who had survived hospitalization for myocardial or cerebral infarction and were discharged home; findings may not generalize to Medicare Advantage enrollees, younger patients, commercially insured populations, or individuals initiating atorvastatin for primary prevention. Additionally, approximately 81% of atorvastatin initiators received high intensity therapy (40 mg or 80 mg). Therefore, the observed associations primarily reflect exposure to high intensity regimens; residual variation in risk across dose levels cannot be excluded. Because the confirmatory analyses used the same data source as the preceding detection study, albeit with refined and alternative outcome definitions, expanded exclusion criteria, and stricter, dual confirmation criteria, independent external validation in a different study population is both warranted and planned.

## CONCLUSION

In this internal confirmatory sequential target trial emulation among statin naïve older Medicare beneficiaries after myocardial or cerebral infarction, several previously detected ADE signals associated with atorvastatin initiation met the rigorous dual confirmation criteria and formed a clear gradient of confirmatory support. At the highest level of support were the early excess risks of acute hemorrhagic cerebrovascular disease, intracranial hemorrhage (traumatic and non-traumatic), and acute hepatic failure, all robust to the absence of concomitant high risk antithrombotic therapy and under optimal covariate balance. Intermediate support was observed for biliary tract disease and musculoskeletal injuries within demographically defined strata. At the lower end of the gradient were cardiac valve disorders and interventions, together with sensory symptoms confirmed only among non-White beneficiaries; the cardiac valve associations, in particular, were substantially attenuated once high risk antithrombotic therapy was accounted for.

These results illustrate the value of a two stage pharmacovigilance framework that pairs high dimensional signal detection with prespecified confirmatory evaluation. Although high intensity atorvastatin remains foundational to secondary prevention in a post infarction population (NNT 28–93), the more robust internally confirmed associations—especially those involving early hemorrhagic stroke (NNH 194-336), hepatic dysfunction (NNH 102-493), and musculoskeletal injury (NNH 69-180)—support heightened clinical vigilance in this higher risk older population. External validation in a different study population is warranted to further elucidate and confirm these findings.

## ETHICS STATEMENT

The research program pertaining to this study, titled “Pharmacovigilance 2.0: Elderly, Disabled, and Disadvantaged Populations”, was reviewed and approved by the Institutional Review Board (IRB) at Rutgers University Human Research Protection Program (HRPP) on March 12, 2024. The IRB classified the research as exempt under categories 4(ii) and 4(iii) of the U.S. Code of Federal Regulations, Title 45, Part 46 (45 CFR 46), determining it poses minimal risk. Category 4(ii) applies to research using existing, anonymized data or records where participants cannot be identified, while category 4(iii) covers benign behavioral interventions or interactions with adults where identifiable information is protected, and disclosure poses no significant risk. The principal investigator, Christopher G. Rowan PhD, conducted the study with approval under eIRB protocol #Pro2024000224, supported by the Veritas Research Foundation.

## DATA AVAILABILITY STATEMENT

This study was completed in part using data resources provided by the Institute for Health Data Core at Rutgers University, available at: http://www.ifhcore.rutgers.edu. The data supporting the research initiative, titled *Pharmacovigilance 2.0: Elderly, Disabled, and Disadvantaged Populations* and conducted under the principal investigator Christopher Rowan, were obtained from the Centers for Medicare & Medicaid Services (CMS) under Data Use Agreement number RSCH-2024-70249. These data contain protected health information subject to the Privacy Act and HIPAA regulations. Due to privacy, confidentiality, and contractual restrictions imposed by CMS, the datasets are not publicly available and cannot be shared by the authors. Qualified researchers interested in accessing similar data may apply independently through the Research Data Assistance Center (ResDAC) at https://resdac.org/ and execute their own Data Use Agreement with CMS.

## ACKNOWLEDGEMENTS

We express our sincere gratitude to Jesse Berlin, ScD, Dan Horton, MD, MSCE, and Tobi Gerhard, BSPharm, PhD, FISPE, for their invaluable scientific and methodologic support. We are deeply thankful to Mathew Iozzio, Haoqian Chen PhD, and the Center for Pharmacoepidemiology & Treatment Science at Rutgers University for their partnership on Pharmacovigilance 2.0. We gratefully acknowledge Minh Tran and Shivani Srivastava for their assistance in developing the outcome definitions used in this study. We also appreciate the Centers for Medicare and Medicaid Services (CMS) for providing data access and support for this initiative. Heartfelt thanks go to the Veritas Research Foundation Board of Directors - Scott C. Durbin, MPH, Mathew Allard, MBA, Bradley J. Buecker, MFA, Gregory Lobdell, MA, and LeeAnn Hard, CPA - for their steadfast trust and support, and to U.S. Medicare beneficiaries for trusting the Veritas Research Foundation to conduct independent drug safety research on their behalf.

## TABLE AND FIGURE LEGEND – SUPPLEMENTAL MATERIAL

**Table S1.** Target Trial Protocol

| Protocol Element | Hypothetical Pragmatic Target Trial<br>(For One Outcome) | Pragmatic Target Trial Emulation<br>(Sequential Trial Approach; For One Outcome) |
| --- | --- | --- |
| **Time Zero Event or Index Event** | Discharged home from an inpatient admission for cerebral infarction (ischemic stroke) or myocardial infarction (MI) during the study period (Jan 1, 2017 to December 31, 2019) | Same Time Zero Event & study period*<br>‡ The time zero event or index event will be classified from inpatient Medicare claims data during the study period using: admission date, discharge date, discharge status (home), length of stay ($\geq 3$ days), ICD-10-CM diagnosis codes pertaining to the admission, and primary diagnosis code field |
| **Time Zero** | Discharge date = Time Zero = date of random treatment assignment | Fourteen sequential trials, in consecutive daily intervals, will be conducted. Each trial will have a unique time zero ($TZ_j$; where $j = 0-13$) beginning on the discharge date ($TZ_0$) |
| **Eligibility Criteria** | Established on discharge date:<br>1. Alive & discharged home<br>2. Enrolled in participating practice<br>3. Age $\geq 65$<br>4. No prior statin use<br>5. No statin contraindications<br>6. No history of outcome | Established on or within 12 months of $TZ_j$:<br>1. No death record on or before $TZ_j$<br>2. $\geq 12$ months Medicare A/B/D enrollment<br>3. Age $\geq 65$<br>4. No prior statin dispensing (excluding $TZ_j$)<br>5. No statin contraindication<br>6. No history of outcome |
| **Treatment Assignment** | Random assignment on discharge date; patients are aware of treatment assignment. | For each sequential trial, treatment "assignment" will be consistent with the observed pharmacy dispensing claims (PDCs) data on $TZ_j$. |
| **Treatment Strategies (pragmatic)** | A0 = No new medication initiation*<br>A1 = Newly initiate atorvastatin*<br>A2 = Newly initiate a medication other than atorvastatin*<br>*\* Initiate treatment strategy upon discharge and continue strategy for six months*<br>*Pragmatic = in addition to other newly started and ongoing medications* | A0 = Same†<br>A1 = Same†<br>A2 = Same†<br>*† Initiate treatment strategy on $TZ_j$ and continue strategy for six months*<br>*Pragmatic = in addition to other newly started and ongoing medications* |
| **Follow-up & Censoring** | ITT: Start: Randomization; End: 6 months.<br>Per-protocol:<br>Start: Randomization date<br>End: 6 months or censor (loss-to-follow-up, death, protocol deviation, or end of study period) | ITT: n/a<br>Per-protocol:<br>Start: $TZ$ date<br>End: 6 months or censor (i.e., Medicare disenrollment, end of study period, or treatment strategy deviation)<br>*Death will be a competing risk not a censoring event* |
| **Outcomes** | Evidence of outcome by healthcare provider during 6-month follow-up period | Evidence of outcome as classified using ICD-10-CM codes into HCUP CCSR categories or ICD-10-CM codes from Sentinel code list or published study |
| **Causal Estimands** | Average treatment effects (ATEs) on the risk scale quantified via the predicted average risk, risk difference (RD), and relative risk (RR) for the following causal contrasts between treatment strategies (i.e., A1 vs A2) | Per-protocol ATE on the subdistribution hazard ratio of A1 vs A2 over 182 days post-discharge, accounting for death as a competing risk and adjusted for baseline confounding and censoring due to treatment strategy deviation through inverse probability weighting (IPTW*IPCW). |
| **Statistical Analysis** | ITT: Cox PH regression; KM survival curve<br>Per-protocol: Cox PH, w/IPCW to account for censoring due to loss to follow-up or treatment strategy deviation | ITT: n/a<br>Per-protocol: Weighted Fine-Gray subdistribution hazard ratio (SHR) with death as a competing risk and robust standard errors clustered on patient identifier. |
| Abbreviations: ICD-10-CM: International Classification of Diseases, 10th Revision, Clinical Modification; HCUP: Healthcare Cost and Utilization Project; CCSR: Clinical Classifications Software Refined; ITT: Intention-to-Treat; KM: Kaplan-Meier; Cox PH: Cox Proportional Hazards; IPTW: Inverse Probability of Treatment Weighting; IPCW: Inverse Probability of Censoring Weighting. |  |  |

**Table S2.** Top 20 Randomly Selected Medications to Classify A2 Discontinuation

| **Randomly Selected A2 Medication** | **Frequency** | **Percent** |
| --- | --- | --- |
| CLOPIDOGREL BISULFATE | 2638 | 13.8% |
| ROSUVASTATIN CALCIUM | 1138 | 5.9% |
| METOPROLOL TARTRATE | 1111 | 5.8% |
| NITROGLYCERIN | 977 | 5.1% |
| LISINOPRIL | 790 | 4.1% |
| METOPROLOL SUCCINATE | 784 | 4.1% |
| CARVEDILOL | 731 | 3.8% |
| PRAVASTATIN SODIUM | 635 | 3.3% |
| TICAGRELOR | 605 | 3.2% |
| APIXABAN | 552 | 2.9% |
| FUROSEMIDE | 550 | 2.9% |
| AMLODIPINE BESYLATE | 481 | 2.5% |
| SIMVASTATIN | 455 | 2.4% |
| ISOSORBIDE MONONITRATE | 414 | 2.2% |
| PANTOPRAZOLE SODIUM | 402 | 2.1% |
| AMIODARONE HCL | 306 | 1.6% |
| POTASSIUM CHLORIDE | 264 | 1.4% |
| LOSARTAN POTASSIUM | 247 | 1.3% |
| LEVOFLOXACIN | 209 | 1.1% |
| EZETIMIBE | 192 | 1.0% |
| WARFARIN SODIUM | 185 | 1.0% |
| **Total** | **13666** | **71.2%** |

**Table S3.**
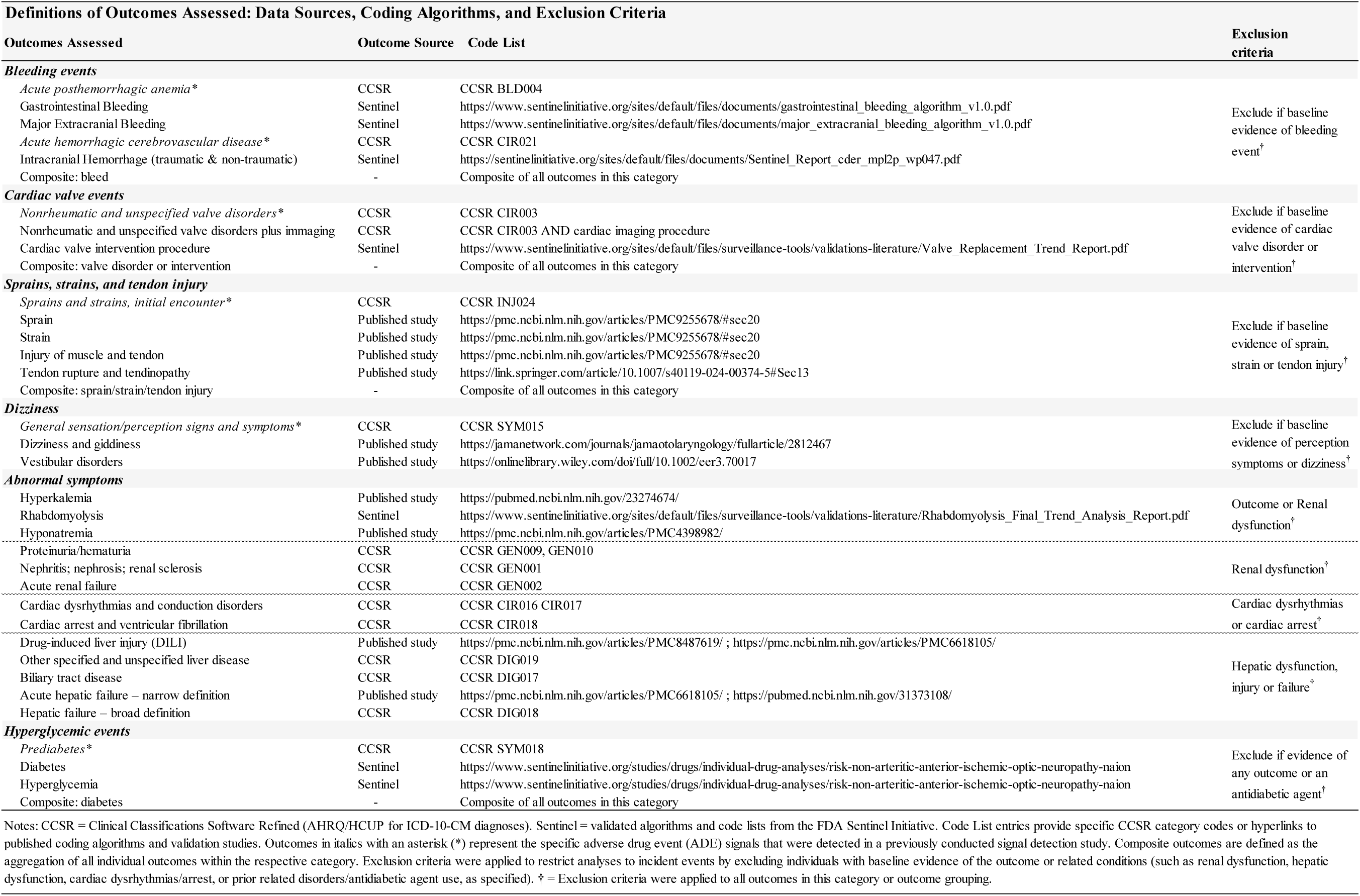
Detailed Outcome Definitions

**Table S4.** Disposition Table

| Eligibility Criteria | N |
| --- | --- |
| Medicare Beneficiary (2017 - 2019) | 20,115,912 |
| + Inpatient Admission | 7,348,255 |
| + Cerebral/myocardial infarction (Time Zero Event) | 694,847 |
| + Age $\geq 65$, LOS $\geq 3$ days & discharged home | 188,071 |
| + Medicare FFS Part A, B, & D<sup>†</sup> | 166,096 |
| + Statin naïve<sup>‡</sup> | 72,270 |
| + No statin contraindication<sup>§</sup> | **70,130** |
| Treatment Strategy<br>A0<br>A1<br>A2<br>N<sub>A0</sub> = 11,000<br>N<sub>A1</sub> = 39,948<br>N<sub>A2</sub> = 19,182 |  |

Abbreviations: N = Number of patients/beneficiaries; LOS = Length of Stay; FFS = Fee-For-Service; Part A = Medicare Hospital Insurance; Part B = Medicare Outpatient Insurance; Part D = Medicare Prescription Drug Coverage Treatment Strategy: A0=Do not initiate a new medication within 14 days post-discharge from Time Zero Event; A1=Initiate atorvastatin within 14 days post-discharge from Time Zero Event; A2=Initiate any other new medication other than atorvastatin within 14 days post-discharge from Time Zero Event. <sup>†</sup> Continuous Medicare Fee-For-Service enrollment in Parts A, B, & D for at least 12 months prior to the Time Zero event <sup>‡</sup> No statin pharmacy dispensing claim or statin related contraindication for at least 12 months prior to the Time Zero event

**Table S5.** Reason for Censoring and Follow-up Time by Treatment Strategy before applying inverse probability of censoring weights

| Reason for End of Follow-up and Distribution of Follow-up Time by Treatment Strategy |  |  |  |  |
| --- | --- | --- | --- | --- |
| Censoring Reason | Treatment Strategy |  |  |  |
|  | A1 | A1 | A2 | A2 |
|  | n | % | n | % |
| Death | 1,578 | 4.0% | 806 | 4.2% |
| Administrative termination (loss of A/B/D coverage) | 464 | 1.2% | 95 | 0.5% |
| Initiated statin (other than atorvastatin) | 2,074 | 5.2% | 6,971 | 36.3% |
| Treatment strategy switch | 5,602 | 14.0% | 2,890 | 15.1% |
| Discontinuation of assigned treatment strategy | 7,484 | 18.7% | 5,167 | 26.9% |
| End of study period before 182 days | 4,632 | 11.6% | 842 | 4.4% |
| Uncensored (completed 182 days of follow-up) | 18,114 | 45.3% | 2411 | 12.6% |
| **Total** | **39,948** | **100.0%** | **19,182** | **100.0%** |

| Time to First Censoring Event | Treatment Strategy |  |
| --- | --- | --- |
|  | A1 | A2 |
| N | 39,948 | 19,182 |
| Mean (SD) | 119 (67) | 52 (64) |
| Median (IQR) | 148 (60–182) | 32 (0–69) |

**Table S6.** Covariate Balance After Inverse-Probability-of-Treatment Weighting Across Sequential Trials of Atorvastatin Initiation Versus Alternative Medication Initiation

|  |  |  |  |  |  |  |  |  |
| --- | --- | --- | --- | --- | --- | --- | --- | --- |
| Confirmed Outcomes | Trials | N | N (A1) | N (A2) | No. of covariates | Mean SMD | Max SMD | Max SMD > 0.1 |
| *Acute hemorrhagic cerebrovascular disease\** | 0 | 31071 | 24773 | 6298 | 573 | 0.009 | 0.037 | No |
| *Acute hemorrhagic cerebrovascular disease\** | 1 | 3155 | 2250 | 905 | 335 | 0.019 | 0.095 | No |
| *Acute hemorrhagic cerebrovascular disease\** | 2 | 576 | 338 | 238 | 174 | 0.046 | 0.148 | Yes |
| *Acute hemorrhagic cerebrovascular disease\** | 3 | 284 | 134 | 150 | 104 | 0.103 | 0.311 | Yes |
| *Acute hemorrhagic cerebrovascular disease\** | 4 | 194 | 95 | 99 | 72 | 0.082 | 0.269 | Yes |
| *Acute hemorrhagic cerebrovascular disease\** | 5 | 154 | 60 | 94 | 53 | 0.147 | 0.414 | Yes |
| *Acute hemorrhagic cerebrovascular disease\** | 6 | 177 | 59 | 118 | 47 | 0.104 | 0.550 | Yes |
| *Acute hemorrhagic cerebrovascular disease\** | 7 | 147 | 54 | 93 | 42 | 0.094 | 0.346 | Yes |
| *Nonrheumatic and unspecified valve disorders\** | 0 | 35030 | 27925 | 7105 | 666 | 0.008 | 0.035 | No |
| *Nonrheumatic and unspecified valve disorders\** | 1 | 3772 | 2654 | 1118 | 396 | 0.015 | 0.050 | No |
| *Nonrheumatic and unspecified valve disorders\** | 2 | 684 | 417 | 267 | 200 | 0.050 | 0.175 | Yes |
| *Nonrheumatic and unspecified valve disorders\** | 3 | 331 | 155 | 176 | 125 | 0.100 | 0.294 | Yes |
| *Nonrheumatic and unspecified valve disorders\** | 4 | 221 | 94 | 127 | 89 | 0.077 | 0.236 | Yes |
| *Nonrheumatic and unspecified valve disorders\** | 5 | 186 | 70 | 116 | 68 | 0.084 | 0.245 | Yes |
| *Nonrheumatic and unspecified valve disorders\** | 6 | 215 | 68 | 147 | 55 | 0.119 | 0.307 | Yes |
| *Nonrheumatic and unspecified valve disorders\** | 7 | 173 | 63 | 110 | 54 | 0.127 | 0.566 | Yes |
| *Nonrheumatic and unspecified valve disorders\** | 8 | 138 | 45 | 93 | 30 | 0.079 | 0.278 | Yes |
| *Nonrheumatic and unspecified valve disorders\** | 9 | 112 | 42 | 70 | 24 | 0.093 | 0.522 | Yes |

**Table S6 (Continued) Covariate Balance After Inverse-Probability-of-Treatment Weighting Across Sequential Trials of Atorvastatin Initiation Versus Alternative Medication Initiation**
|  |  |  |  |  |  |  |  |  |
| --- | --- | --- | --- | --- | --- | --- | --- | --- |
| Confirmed Outcomes | Trials | N | N (A1) | N (A2) | No. of covariates | Mean SMD | Max SMD | Max SMD > 0.1 |
| Nonrheumatic and unspecified valve disorders plus imaging | 0 | 35030 | 27925 | 7105 | 666 | 0.008 | 0.035 | No |
| Nonrheumatic and unspecified valve disorders plus imaging | 1 | 3772 | 2654 | 1118 | 396 | 0.015 | 0.050 | No |
| Nonrheumatic and unspecified valve disorders plus imaging | 2 | 684 | 417 | 267 | 200 | 0.050 | 0.175 | Yes |
| Nonrheumatic and unspecified valve disorders plus imaging | 3 | 331 | 155 | 176 | 125 | 0.100 | 0.294 | Yes |
| Nonrheumatic and unspecified valve disorders plus imaging | 4 | 221 | 94 | 127 | 89 | 0.077 | 0.236 | Yes |
| Nonrheumatic and unspecified valve disorders plus imaging | 5 | 186 | 70 | 116 | 68 | 0.084 | 0.245 | Yes |
| Nonrheumatic and unspecified valve disorders plus imaging | 6 | 216 | 68 | 148 | 55 | 0.112 | 0.296 | Yes |
| Nonrheumatic and unspecified valve disorders plus imaging | 7 | 173 | 63 | 110 | 54 | 0.127 | 0.566 | Yes |
| Nonrheumatic and unspecified valve disorders plus imaging | 8 | 139 | 45 | 94 | 30 | 0.081 | 0.286 | Yes |
| Nonrheumatic and unspecified valve disorders plus imaging | 9 | 112 | 42 | 70 | 24 | 0.093 | 0.522 | Yes |
| Cardiac valve intervention procedure | 0 | 35030 | 27925 | 7105 | 666 | 0.008 | 0.035 | No |
| Cardiac valve intervention procedure | 1 | 3777 | 2657 | 1120 | 396 | 0.015 | 0.050 | No |
| Cardiac valve intervention procedure | 2 | 688 | 418 | 270 | 202 | 0.051 | 0.151 | Yes |
| Cardiac valve intervention procedure | 3 | 336 | 156 | 180 | 126 | 0.097 | 0.294 | Yes |
| Cardiac valve intervention procedure | 4 | 224 | 94 | 130 | 89 | 0.085 | 0.267 | Yes |
| Cardiac valve intervention procedure | 5 | 193 | 71 | 122 | 69 | 0.142 | 0.391 | Yes |
| Cardiac valve intervention procedure | 6 | 221 | 70 | 151 | 57 | 0.098 | 0.280 | Yes |
| Cardiac valve intervention procedure | 7 | 174 | 64 | 110 | 55 | 0.113 | 0.549 | Yes |
| Cardiac valve intervention procedure | 8 | 145 | 47 | 98 | 32 | 0.085 | 0.249 | Yes |
| Cardiac valve intervention procedure | 9 | 114 | 44 | 70 | 28 | 0.097 | 0.486 | Yes |
| Composite: valve | 0 | 35030 | 27925 | 7105 | 666 | 0.008 | 0.035 | No |
| Composite: valve | 1 | 3772 | 2654 | 1118 | 396 | 0.015 | 0.050 | No |
| Composite: valve | 2 | 684 | 417 | 267 | 200 | 0.050 | 0.175 | Yes |
| Composite: valve | 3 | 331 | 155 | 176 | 125 | 0.100 | 0.294 | Yes |
| Composite: valve | 4 | 221 | 94 | 127 | 89 | 0.077 | 0.236 | Yes |
| Composite: valve | 5 | 186 | 70 | 116 | 68 | 0.084 | 0.245 | Yes |
| Composite: valve | 6 | 215 | 68 | 147 | 55 | 0.119 | 0.307 | Yes |
| Composite: valve | 7 | 173 | 63 | 110 | 54 | 0.127 | 0.566 | Yes |
| Composite: valve | 8 | 138 | 45 | 93 | 30 | 0.079 | 0.278 | Yes |
| Composite: valve | 9 | 112 | 42 | 70 | 24 | 0.093 | 0.522 | Yes |

**Table S6 (Continued) Covariate Balance After Inverse-Probability-of-Treatment Weighting Across Sequential Trials of Atorvastatin Initiation Versus Alternative Medication Initiation**
|  |  |  |  |  |  |  |  |  |
| --- | --- | --- | --- | --- | --- | --- | --- | --- |
| Confirmed Outcomes | Trials | N | N (A1) | N (A2) | No. of covariates | Mean SMD | Max SMD | Max SMD > 0.1 |
| *Sprains and strains, initial encounter\** | 0 | 42839 | 33268 | 9571 | 720 | 0.006 | 0.029 | No |
| *Sprains and strains, initial encounter\** | 1 | 4624 | 3194 | 1430 | 434 | 0.013 | 0.042 | No |
| *Sprains and strains, initial encounter\** | 2 | 867 | 510 | 357 | 240 | 0.040 | 0.149 | Yes |
| *Sprains and strains, initial encounter\** | 3 | 468 | 210 | 258 | 171 | 0.082 | 0.275 | Yes |
| *Sprains and strains, initial encounter\** | 4 | 292 | 131 | 161 | 123 | 0.070 | 0.241 | Yes |
| *Sprains and strains, initial encounter\** | 5 | 266 | 100 | 166 | 107 | 0.135 | 0.346 | Yes |
| *Sprains and strains, initial encounter\** | 6 | 263 | 78 | 185 | 73 | 0.106 | 0.394 | Yes |
| *Sprains and strains, initial encounter\** | 7 | 219 | 81 | 138 | 82 | 0.136 | 0.416 | Yes |
| *Sprains and strains, initial encounter\** | 8 | 175 | 50 | 125 | 36 | 0.102 | 0.376 | Yes |
| *Sprains and strains, initial encounter\** | 9 | 133 | 50 | 83 | 42 | 0.154 | 0.727 | Yes |
| *Sprains and strains, initial encounter\** | 10 | 173 | 41 | 132 | 20 | 0.127 | 0.491 | Yes |
| Sprain | 0 | 42839 | 33268 | 9571 | 720 | 0.006 | 0.029 | No |
| Sprain | 1 | 4625 | 3194 | 1431 | 434 | 0.013 | 0.043 | No |
| Sprain | 2 | 868 | 510 | 358 | 240 | 0.040 | 0.148 | Yes |
| Sprain | 3 | 468 | 210 | 258 | 171 | 0.082 | 0.275 | Yes |
| Sprain | 4 | 292 | 131 | 161 | 123 | 0.070 | 0.241 | Yes |
| Sprain | 5 | 266 | 100 | 166 | 107 | 0.135 | 0.346 | Yes |
| Sprain | 6 | 263 | 78 | 185 | 73 | 0.106 | 0.394 | Yes |
| Sprain | 7 | 219 | 81 | 138 | 82 | 0.136 | 0.416 | Yes |
| Sprain | 8 | 175 | 50 | 125 | 36 | 0.102 | 0.376 | Yes |
| Sprain | 9 | 133 | 50 | 83 | 42 | 0.154 | 0.727 | Yes |
| Sprain | 10 | 173 | 41 | 132 | 20 | 0.127 | 0.491 | Yes |
| Composite: sprain/strain/tendon injury | 0 | 42839 | 33268 | 9571 | 720 | 0.006 | 0.029 | No |
| Composite: sprain/strain/tendon injury | 1 | 4624 | 3194 | 1430 | 434 | 0.013 | 0.042 | No |
| Composite: sprain/strain/tendon injury | 2 | 867 | 510 | 357 | 240 | 0.040 | 0.149 | Yes |
| Composite: sprain/strain/tendon injury | 3 | 468 | 210 | 258 | 171 | 0.082 | 0.275 | Yes |
| Composite: sprain/strain/tendon injury | 4 | 292 | 131 | 161 | 123 | 0.070 | 0.241 | Yes |
| Composite: sprain/strain/tendon injury | 5 | 266 | 100 | 166 | 107 | 0.135 | 0.346 | Yes |
| Composite: sprain/strain/tendon injury | 6 | 263 | 78 | 185 | 73 | 0.106 | 0.394 | Yes |
| Composite: sprain/strain/tendon injury | 7 | 219 | 81 | 138 | 82 | 0.136 | 0.416 | Yes |
| Composite: sprain/strain/tendon injury | 8 | 175 | 50 | 125 | 36 | 0.102 | 0.376 | Yes |
| Composite: sprain/strain/tendon injury | 9 | 133 | 50 | 83 | 42 | 0.154 | 0.727 | Yes |
| Composite: sprain/strain/tendon injury | 10 | 171 | 40 | 131 | 20 | 0.129 | 0.484 | Yes |

**Table S6 (Continued) Covariate Balance After Inverse-Probability-of-Treatment Weighting Across Sequential Trials of Atorvastatin Initiation Versus Alternative Medication Initiation**
|  |  |  |  |  |  |  |  |  |
| --- | --- | --- | --- | --- | --- | --- | --- | --- |
| Confirmed Outcomes | Trials | N | N (A1) | N (A2) | No. of covariates | Mean SMD | Max SMD | Max SMD > 0.1 |
| *General sensation/perception signs and symptoms\** | 0 | 39956 | 31040 | 8916 | 710 | 0.006 | 0.034 | **No** |
| *General sensation/perception signs and symptoms\** | 1 | 4204 | 2896 | 1308 | 425 | 0.015 | 0.068 | **No** |
| *General sensation/perception signs and symptoms\** | 2 | 784 | 459 | 325 | 225 | 0.045 | 0.240 | Yes |
| *General sensation/perception signs and symptoms\** | 3 | 406 | 185 | 221 | 150 | 0.082 | 0.293 | Yes |
| *General sensation/perception signs and symptoms\** | 4 | 256 | 114 | 142 | 104 | 0.098 | 0.264 | Yes |
| *General sensation/perception signs and symptoms\** | 5 | 231 | 91 | 140 | 95 | 0.150 | 0.387 | Yes |
| *General sensation/perception signs and symptoms\** | 6 | 228 | 66 | 162 | 56 | 0.115 | 0.393 | Yes |
| *General sensation/perception signs and symptoms\** | 7 | 198 | 72 | 126 | 74 | 0.117 | 0.609 | Yes |
| *General sensation/perception signs and symptoms\** | 8 | 159 | 47 | 112 | 31 | 0.086 | 0.214 | Yes |
| *General sensation/perception signs and symptoms\** | 9 | 113 | 40 | 73 | 18 | 0.148 | 0.409 | Yes |
| Dizziness and giddiness | 0 | 39956 | 31040 | 8916 | 710 | 0.006 | 0.034 | **No** |
| Dizziness and giddiness | 1 | 4204 | 2896 | 1308 | 425 | 0.015 | 0.068 | **No** |
| Dizziness and giddiness | 2 | 784 | 459 | 325 | 225 | 0.045 | 0.240 | Yes |
| Dizziness and giddiness | 3 | 407 | 185 | 222 | 150 | 0.081 | 0.305 | Yes |
| Dizziness and giddiness | 4 | 256 | 114 | 142 | 104 | 0.098 | 0.264 | Yes |
| Dizziness and giddiness | 5 | 231 | 91 | 140 | 95 | 0.150 | 0.387 | Yes |
| Dizziness and giddiness | 6 | 228 | 66 | 162 | 56 | 0.115 | 0.393 | Yes |
| Dizziness and giddiness | 7 | 199 | 72 | 127 | 74 | 0.116 | 0.604 | Yes |
| Dizziness and giddiness | 8 | 159 | 47 | 112 | 31 | 0.086 | 0.214 | Yes |
| Dizziness and giddiness | 9 | 113 | 40 | 73 | 18 | 0.148 | 0.409 | Yes |

**Table S6 (Continued) Covariate Balance After Inverse-Probability-of-Treatment Weighting Across Sequential Trials of Atorvastatin Initiation Versus Alternative Medication Initiation**
|  |  |  |  |  |  |  |  |  |
| --- | --- | --- | --- | --- | --- | --- | --- | --- |
| Confirmed Outcomes | Trials | N | N (A1) | N (A2) | No. of covariates | Mean SMD | Max SMD | Max SMD > 0.1 |
| Acute hepatic failure – narrow definition | 0 | 42240 | 32944 | 9296 | 701 | 0.007 | 0.037 | No |
| Acute hepatic failure – narrow definition | 1 | 4504 | 3131 | 1373 | 425 | 0.013 | 0.053 | No |
| Acute hepatic failure – narrow definition | 2 | 846 | 500 | 346 | 232 | 0.033 | 0.103 | Yes |
| Acute hepatic failure – narrow definition | 3 | 443 | 204 | 239 | 166 | 0.070 | 0.218 | Yes |
| Acute hepatic failure – narrow definition | 4 | 281 | 126 | 155 | 112 | 0.097 | 0.286 | Yes |
| Acute hepatic failure – narrow definition | 5 | 252 | 94 | 158 | 95 | 0.078 | 0.226 | Yes |
| Acute hepatic failure – narrow definition | 6 | 263 | 79 | 184 | 71 | 0.112 | 0.478 | Yes |
| Acute hepatic failure – narrow definition | 7 | 216 | 79 | 137 | 81 | 0.129 | 0.415 | Yes |
| Acute hepatic failure – narrow definition | 8 | 175 | 53 | 122 | 44 | 0.107 | 0.513 | Yes |
| Acute hepatic failure – narrow definition | 9 | 130 | 49 | 81 | 39 | 0.166 | 0.525 | Yes |
| Biliary tract disease | 0 | 42240 | 32944 | 9296 | 701 | 0.007 | 0.037 | No |
| Biliary tract disease | 1 | 4504 | 3131 | 1373 | 425 | 0.013 | 0.053 | No |
| Biliary tract disease | 2 | 844 | 498 | 346 | 232 | 0.036 | 0.118 | Yes |
| Biliary tract disease | 3 | 442 | 203 | 239 | 165 | 0.067 | 0.234 | Yes |
| Biliary tract disease | 4 | 281 | 126 | 155 | 112 | 0.097 | 0.286 | Yes |
| Biliary tract disease | 5 | 252 | 94 | 158 | 95 | 0.078 | 0.226 | Yes |
| Biliary tract disease | 6 | 262 | 79 | 183 | 71 | 0.108 | 0.478 | Yes |
| Biliary tract disease | 7 | 215 | 79 | 136 | 81 | 0.130 | 0.414 | Yes |
| Biliary tract disease | 8 | 175 | 53 | 122 | 44 | 0.107 | 0.513 | Yes |
| Biliary tract disease | 9 | 131 | 49 | 82 | 39 | 0.167 | 0.503 | Yes |
| Biliary tract disease | 10 | 164 | 40 | 124 | 16 | 0.130 | 0.457 | Yes |

Notes: Trials = sequential trial number (0–7); N = total number of eligible participants in the trial; N(A1) = number of participants initiating atorvastatin (A1); N(A2) = number of participants initiating a different medication (A2); No. of covariates = number of covariates included in the inverse-probability-of-treatment weighting (IPTW) model for that trial; Mean SMD = mean absolute standardized mean difference across all covariates after IPTW; Max SMD = maximum absolute standardized mean difference across all covariates after IPTW; Max SMD > 0.1 = indicator of whether the maximum SMD exceeds the conventional threshold of 0.1 for adequate covariate balance (Yes/No). A1 = atorvastatin initiation; A2 = initiation of a different medication.

**Table S7.** Detailed Results for Confirmed ADE Signals

| Detailed Results for Confirmed ADE Signals |  |  |  |  |  |  |  |  |  |  |  |  |  |  |  |  |  |
| --- | --- | --- | --- | --- | --- | --- | --- | --- | --- | --- | --- | --- | --- | --- | --- | --- | --- |
| Category : Bleeding Events |  |  |  |  |  |  |  |  |  |  |  |  |  |  |  |  |  |
| Outcome : Acute hemorrhagic cerebrovascular disease |  |  |  |  |  |  |  |  |  |  |  |  |  |  |  |  |  |
| Analysis Set | Stratum | ADE Signal Confirmation |  |  | Weighted Fine-Gray |  |  |  | Unadjusted Descriptive Analyses for All Trials |  |  |  |  |  |  |  |  |
|  |  | q | P(adj>1) | Confirmed | sHR | LCL | UCL | p | N | A | B | C | D | Risk A2 | Risk A1 | RD | NNH |
| All Trials | Overall | 0.050 | 1.00 | Yes | 1.43 | 1.00 | 2.04 | 0.050 | 35758 | 7947 | 27461 | 48 | 302 | 0.60% | 1.09% | 0.50% | 205 |
| All Trials | Time stratified: 1-30 days | 0.002 | 1.00 | Yes | 2.20 | 1.35 | 3.58 | 0.002 | 35758 | 7972 | 27604 | 23 | 159 | 0.29% | 0.57% | 0.30% | 351 |
| All Trials | Time stratified: 31-91 days | † | † | † | 0.63 | 0.36 | 1.12 | 0.113 | 29007 | 5519 | 23383 | 19 | 86 | 0.34% | 0.37% | 0.00% | 4281 |
| All Trials | Time stratified: 92-182 days | † | † | † | 1.96 | 0.81 | 4.74 | 0.136 | 19904 | 2414 | 17427 | * | 57 | * | 0.33% | * | * |
| All Trials | Age 65-74 years | 0.424 | 0.84 | No | 1.24 | 0.73 | 2.09 | 0.424 | 17533 | 3430 | 13942 | 22 | 139 | 0.64% | 0.99% | 0.30% | 286 |
| All Trials | Age 75+ years | 0.137 | 1.00 | No | 1.61 | 0.99 | 2.62 | 0.055 | 18225 | 4517 | 13519 | 26 | 163 | 0.57% | 1.19% | 0.60% | 162 |
| All Trials | Male | 0.219 | 1.00 | No | 1.53 | 0.88 | 2.67 | 0.132 | 17849 | 3709 | 13969 | 22 | 149 | 0.59% | 1.06% | 0.50% | 215 |
| All Trials | Female | 0.266 | 0.99 | No | 1.34 | 0.85 | 2.13 | 0.213 | 17909 | 4238 | 13492 | 26 | 153 | 0.61% | 1.12% | 0.50% | 195 |
| All Trials | White Race | 0.142 | 1.00 | No | 1.58 | 1.05 | 2.39 | 0.028 | 30166 | 6823 | 23064 | 37 | 242 | 0.54% | 1.04% | 0.50% | 200 |
| All Trials | Non-White Race | † | † | † | 1.00 | 0.49 | 2.05 | 0.995 | 5592 | 1124 | 4397 | 11 | 60 | 0.97% | 1.35% | 0.40% | 265 |
| Trials 0 & 1 | Overall | 0.039 | 1.00 | Yes | 1.50 | 1.02 | 2.20 | 0.039 | 34226 | 7162 | 26730 | 41 | 293 | 0.57% | 1.08% | 0.50% | 194 |
| Trials 0 & 1 | Time stratified: 1-30 days | † | † | † | 2.71 | 1.59 | 4.62 | 0.000 | 34226 | 7185 | 26869 | 18 | 154 | 0.25% | 0.57% | 0.30% | 313 |
| Trials 0 & 1 | Time stratified: 31-91 days | † | † | † | 0.60 | 0.33 | 1.07 | 0.085 | 27803 | 4944 | 22756 | 18 | 85 | 0.36% | 0.37% | 0.00% | 10660 |
| Trials 0 & 1 | Time stratified: 92-182 days | † | † | † | 2.06 | 0.78 | 5.43 | 0.144 | 19229 | 2180 | 16990 | * | 54 | * | 0.32% | * | * |
| Trials 0 & 1 | Age 65-74 years | † | † | † | 1.30 | 0.72 | 2.32 | 0.385 | 16854 | 3092 | 13612 | 17 | 133 | 0.55% | 0.97% | 0.40% | 238 |
| Trials 0 & 1 | Age 75+ years | 0.063 | 1.00 | No | 1.70 | 1.02 | 2.82 | 0.042 | 17372 | 4070 | 13118 | 24 | 160 | 0.59% | 1.21% | 0.60% | 162 |
| Trials 0 & 1 | Male | † | † | † | 1.58 | 0.86 | 2.88 | 0.137 | 17146 | 3357 | 13627 | 19 | 143 | 0.56% | 1.04% | 0.50% | 210 |
| Trials 0 & 1 | Female | 0.155 | 1.00 | No | 1.43 | 0.87 | 2.36 | 0.155 | 17080 | 3805 | 13103 | 22 | 150 | 0.57% | 1.13% | 0.60% | 180 |
| Trials 0 & 1 | White Race | 0.114 | 1.00 | No | 1.59 | 1.03 | 2.45 | 0.038 | 28955 | 6162 | 22524 | 33 | 236 | 0.53% | 1.04% | 0.50% | 198 |
| Trials 0 & 1 | Non-White Race | † | † | † | 1.20 | 0.53 | 2.74 | 0.658 | 5271 | 1000 | 4206 | * | 57 | * | 1.34% | * | * |

**Table S7 (Continued) Detailed Results for Confirmed ADE Signals**
| Detailed Results for Confirmed ADE Signals |  |  |  |  |  |  |  |  |  |  |  |  |  |  |  |  |  |
| --- | --- | --- | --- | --- | --- | --- | --- | --- | --- | --- | --- | --- | --- | --- | --- | --- | --- |
| Category : Bleeding Events |  |  |  |  |  |  |  |  |  |  |  |  |  |  |  |  |  |
| Outcome : Intracranial Hemorrhage (traumatic & non-traumatic) |  |  |  |  |  |  |  |  |  |  |  |  |  |  |  |  |  |
| Analysis Set | Stratum | ADE Signal Confirmation |  |  | Weighted Fine-Gray |  |  |  | Unadjusted Descriptive Analyses for All Trials |  |  |  |  |  |  |  |  |
|  |  | q | P(adj>1) | Confirmed | sHR | LCL | UCL | p | N | A | B | C | D | Risk A2 | Risk A1 | RD | NNH |
| All Trials | Overall | 0.073 | 1.00 | No | 1.39 | 0.97 | 2.00 | 0.073 | 35758 | 7946 | 27442 | 49 | 321 | 0.61% | 1.16% | 0.50% | 184 |
| All Trials | Time stratified: 1-30 days | 0.016 | 1.00 | Yes | 1.91 | 1.13 | 3.23 | 0.016 | 35758 | 7971 | 27597 | 24 | 166 | 0.30% | 0.60% | 0.30% | 336 |
| All Trials | Time stratified: 31-91 days | † | † | † | 0.69 | 0.39 | 1.21 | 0.197 | 29000 | 5518 | 23371 | 19 | 92 | 0.34% | 0.39% | 0.00% | 2042 |
| All Trials | Time stratified: 92-182 days | † | † | † | 2.13 | 0.88 | 5.13 | 0.092 | 19892 | 2414 | 17409 | * | 63 | * | 0.36% | * | * |
| All Trials | Age 65-74 years | 0.778 | 0.27 | No | 1.08 | 0.63 | 1.85 | 0.778 | 17533 | 3429 | 13939 | 23 | 142 | 0.67% | 1.01% | 0.30% | 292 |
| All Trials | Age 75+ years | 0.126 | 1.00 | No | 1.74 | 1.07 | 2.82 | 0.025 | 18225 | 4517 | 13503 | 26 | 179 | 0.57% | 1.31% | 0.70% | 136 |
| All Trials | Male | 0.159 | 1.00 | No | 1.60 | 0.92 | 2.78 | 0.095 | 17849 | 3709 | 13961 | 22 | 157 | 0.59% | 1.11% | 0.50% | 191 |
| All Trials | Female | 0.462 | 0.86 | No | 1.24 | 0.77 | 2.00 | 0.369 | 17909 | 4237 | 13481 | 27 | 164 | 0.63% | 1.20% | 0.60% | 176 |
| All Trials | White Race | 0.143 | 1.00 | No | 1.50 | 0.99 | 2.29 | 0.057 | 30166 | 6822 | 23048 | 38 | 258 | 0.55% | 1.11% | 0.60% | 181 |
| All Trials | Non-White Race | † | † | † | 1.04 | 0.51 | 2.14 | 0.904 | 5592 | 1124 | 4394 | 11 | 63 | 0.97% | 1.41% | 0.40% | 225 |
| Trials 0 & 1 | Overall | 0.067 | 1.00 | No | 1.44 | 0.98 | 2.14 | 0.067 | 34226 | 7161 | 26711 | 42 | 312 | 0.58% | 1.15% | 0.60% | 175 |
| Trials 0 & 1 | Time stratified: 1-30 days | † | † | † | 2.21 | 1.21 | 4.03 | 0.010 | 34226 | 7184 | 26862 | 19 | 161 | 0.26% | 0.60% | 0.30% | 301 |
| Trials 0 & 1 | Time stratified: 31-91 days | † | † | † | 0.65 | 0.36 | 1.17 | 0.153 | 27796 | 4943 | 22744 | 18 | 91 | 0.36% | 0.40% | 0.00% | 2803 |
| Trials 0 & 1 | Time stratified: 92-182 days | † | † | † | 2.25 | 0.86 | 5.90 | 0.100 | 19217 | 2180 | 16972 | * | 60 | * | 0.35% | * | * |
| Trials 0 & 1 | Age 65-74 years | † | † | † | 1.10 | 0.61 | 1.99 | 0.749 | 16854 | 3091 | 13609 | 18 | 136 | 0.58% | 0.99% | 0.40% | 244 |
| Trials 0 & 1 | Age 75+ years | 0.057 | 1.00 | No | 1.83 | 1.10 | 3.04 | 0.019 | 17372 | 4070 | 13102 | 24 | 176 | 0.59% | 1.33% | 0.70% | 135 |
| Trials 0 & 1 | Male | † | † | † | 1.65 | 0.91 | 3.00 | 0.102 | 17146 | 3357 | 13619 | 19 | 151 | 0.56% | 1.10% | 0.50% | 187 |
| Trials 0 & 1 | Female | 0.321 | 0.96 | No | 1.30 | 0.77 | 2.18 | 0.321 | 17080 | 3804 | 13092 | 23 | 161 | 0.60% | 1.21% | 0.60% | 163 |
| Trials 0 & 1 | White Race | 0.120 | 1.00 | No | 1.49 | 0.95 | 2.33 | 0.080 | 28955 | 6161 | 22508 | 34 | 252 | 0.55% | 1.11% | 0.60% | 179 |
| Trials 0 & 1 | Non-White Race | † | † | † | 1.26 | 0.56 | 2.87 | 0.576 | 5271 | 1000 | 4203 | * | 60 | * | 1.41% | * | * |
| Notes: A1 = Atorvastatin initiation; A2 = Newly initiated a different medication other than atorvastatin; N = Total number of patients in the analytic stratum; A = Number of A2 patients without the event; B = Number of A1 patients without the event; C = Number of A2 patients with the event; D = Number of A1 patients with the event; Risk A1 = Cumulative incidence (risk) in the A1 (atorvastatin) group; Risk A2 = Cumulative incidence (risk) in the A2 (comparator) group; RD = Risk difference; NNH = Number needed to harm. ADE Signal Confirmation metrics: q = Within-outcome Benjamini-Hochberg false-discovery-rate adjusted p-value; P(adj>1) = Proportion of 5,000 Monte Carlo draws from the probabilistic quantitative bias analysis in which the residual-confounding-adjusted subdistribution hazard ratio remained greater than 1. Confirmed = 1 if q ≤ 0.05 and P(adj>1) = 1; otherwise 0. Only analyses with at least 20 events in both treatment arms (C ≥ 20 and D ≥ 20) were included in the ADE Signal Confirmation analyses. Analyses that did not meet this threshold were excluded from both the Benjamini-Hochberg correction and the probabilistic quantitative bias analysis (denoted by †). * = Not reportable due to cell counts ≤ 10 per CMS policies. |  |  |  |  |  |  |  |  |  |  |  |  |  |  |  |  |  |

**Table S7 (Continued) Detailed Results for Confirmed ADE Signals**
| Detailed Results for Confirmed ADE Signals |  |  |  |  |  |  |  |  |  |  |  |  |  |  |  |  |  |
| --- | --- | --- | --- | --- | --- | --- | --- | --- | --- | --- | --- | --- | --- | --- | --- | --- | --- |
| Category : Cardiac valve events |  |  |  |  |  |  |  |  |  |  |  |  |  |  |  |  |  |
| Outcome : Nonrheumatic and unspecified valve disorders |  |  |  |  |  |  |  |  |  |  |  |  |  |  |  |  |  |
| Analysis Set | Stratum | ADE Signal Confirmation |  |  | Weighted Fine-Gray |  |  |  | Unadjusted Descriptive Analyses for All Trials |  |  |  |  |  |  |  |  |
|  |  | q | P(adj>1) | Confirmed | sHR | LCL | UCL | p | N | A | B | C | D | Risk A2 | Risk A1 | RD | NNH |
| All Trials | Overall | 0.153 | 0.28 | No | 1.08 | 0.97 | 1.19 | 0.153 | 40862 | 8622 | 28328 | 707 | 3205 | 7.58% | 10.16% | 2.60% | 39 |
| All Trials | Time stratified: 1-30 days | 0.871 | 0.00 | No | 0.99 | 0.86 | 1.13 | 0.871 | 40862 | 8924 | 30218 | 405 | 1315 | 4.34% | 4.17% | -0.20% | - |
| All Trials | Time stratified: 31-91 days | 1.133 | 0.02 | No | 1.03 | 0.86 | 1.23 | 0.756 | 31729 | 5943 | 24461 | 222 | 1103 | 3.60% | 4.31% | 0.70% | 140 |
| All Trials | Time stratified: 92-182 days | 0.013 | 1.00 | Yes | 1.48 | 1.13 | 1.94 | 0.004 | 20625 | 2396 | 17362 | 80 | 787 | 3.23% | 4.34% | 1.10% | 90 |
| All Trials | Age 65-74 years | 0.588 | 0.17 | No | 1.06 | 0.90 | 1.25 | 0.490 | 20283 | 3878 | 14634 | 257 | 1514 | 6.22% | 9.38% | 3.20% | 32 |
| All Trials | Age 75+ years | 0.402 | 0.38 | No | 1.10 | 0.97 | 1.25 | 0.134 | 20579 | 4744 | 13694 | 450 | 1691 | 8.66% | 10.99% | 2.30% | 43 |
| All Trials | Male | 0.506 | 0.26 | No | 1.08 | 0.93 | 1.25 | 0.337 | 20386 | 4067 | 14420 | 302 | 1597 | 6.91% | 9.97% | 3.10% | 33 |
| All Trials | Female | 0.544 | 0.26 | No | 1.08 | 0.94 | 1.23 | 0.272 | 20476 | 4555 | 13908 | 405 | 1608 | 8.17% | 10.36% | 2.20% | 45 |
| All Trials | White Race | 0.716 | 0.34 | No | 1.09 | 0.98 | 1.21 | 0.119 | 34051 | 7275 | 23459 | 600 | 2717 | 7.62% | 10.38% | 2.80% | 36 |
| All Trials | Non-White Race | 0.996 | 0.00 | No | 1.00 | 0.77 | 1.29 | 0.996 | 6811 | 1347 | 4869 | 107 | 488 | 7.36% | 9.11% | 1.80% | 57 |
| Trials 0 & 1 | Overall | 0.399 | 0.10 | No | 1.05 | 0.94 | 1.16 | 0.399 | 38802 | 7597 | 27483 | 626 | 3096 | 7.61% | 10.12% | 2.50% | 40 |
| Trials 0 & 1 | Time stratified: 1-30 days | 0.832 | 0.00 | No | 0.96 | 0.83 | 1.10 | 0.555 | 38802 | 7865 | 29304 | 358 | 1275 | 4.35% | 4.17% | -0.20% | - |
| Trials 0 & 1 | Time stratified: 31-91 days | 0.815 | 0.00 | No | 0.98 | 0.81 | 1.18 | 0.815 | 30163 | 5184 | 23716 | 201 | 1062 | 3.73% | 4.29% | 0.60% | 181 |
| Trials 0 & 1 | Time stratified: 92-182 days | 0.006 | 1.00 | Yes | 1.58 | 1.18 | 2.11 | 0.002 | 19836 | 2134 | 16876 | 67 | 759 | 3.04% | 4.30% | 1.30% | 79 |
| Trials 0 & 1 | Age 65-74 years | 0.668 | 0.13 | No | 1.05 | 0.89 | 1.25 | 0.557 | 19348 | 3430 | 14222 | 229 | 1467 | 6.26% | 9.35% | 3.10% | 32 |
| Trials 0 & 1 | Age 75+ years | 0.889 | 0.12 | No | 1.05 | 0.92 | 1.20 | 0.445 | 19454 | 4167 | 13261 | 397 | 1629 | 8.70% | 10.94% | 2.20% | 45 |
| Trials 0 & 1 | Male | 0.768 | 0.01 | No | 1.02 | 0.87 | 1.20 | 0.768 | 19437 | 3584 | 14037 | 272 | 1544 | 7.05% | 9.91% | 2.90% | 35 |
| Trials 0 & 1 | Female | 1.070 | 0.21 | No | 1.07 | 0.93 | 1.23 | 0.357 | 19365 | 4013 | 13446 | 354 | 1552 | 8.11% | 10.35% | 2.20% | 45 |
| Trials 0 & 1 | White Race | 1.316 | 0.24 | No | 1.07 | 0.96 | 1.20 | 0.219 | 32454 | 6446 | 22848 | 529 | 2631 | 7.58% | 10.33% | 2.70% | 36 |
| Trials 0 & 1 | Non-White Race | 0.741 | 0.00 | No | 0.91 | 0.70 | 1.19 | 0.494 | 6348 | 1151 | 4635 | 97 | 465 | 7.77% | 9.12% | 1.30% | 74 |

**Table S7 (Continued) Detailed Results for Confirmed ADE Signals**
| Detailed Results for Confirmed ADE Signals |  |  |  |  |  |  |  |  |  |  |  |  |  |  |  |  |  |
| --- | --- | --- | --- | --- | --- | --- | --- | --- | --- | --- | --- | --- | --- | --- | --- | --- | --- |
| Category : Cardiac valve events |  |  |  |  |  |  |  |  |  |  |  |  |  |  |  |  |  |
| Outcome : Nonrheumatic and unspecified valve disorders plus imaging |  |  |  |  |  |  |  |  |  |  |  |  |  |  |  |  |  |
| Analysis Set | Stratum | ADE Signal Confirmation |  |  | Weighted Fine-Gray |  |  |  | Unadjusted Descriptive Analyses for All Trials |  |  |  |  |  |  |  |  |
|  |  | q | P(adj>1) | Confirmed | sHR | LCL | UCL | p | N | A | B | C | D | Risk A2 | Risk A1 | RD | NNH |
| All Trials | Overall | 0.107 | 0.31 | No | 1.09 | 0.98 | 1.21 | 0.107 | 40864 | 8659 | 28412 | 672 | 3121 | 7.20% | 9.90% | 2.70% | 37 |
| All Trials | Time stratified: 1-30 days | 0.960 | 0.00 | No | 1.00 | 0.87 | 1.15 | 0.960 | 40864 | 8950 | 30266 | 381 | 1267 | 4.08% | 4.02% | -0.10% |  |
| All Trials | Time stratified: 31-91 days | 0.961 | 0.08 | No | 1.04 | 0.87 | 1.25 | 0.641 | 31792 | 5972 | 24525 | 214 | 1081 | 3.46% | 4.22% | 0.80% | 131 |
| All Trials | Time stratified: 92-182 days | 0.011 | 1.00 | Yes | 1.50 | 1.14 | 1.98 | 0.004 | 20681 | 2409 | 17422 | 77 | 773 | 3.10% | 4.25% | 1.20% | 87 |
| All Trials | Age 65-74 years | 0.497 | 0.24 | No | 1.07 | 0.91 | 1.27 | 0.414 | 20283 | 3890 | 14668 | 245 | 1480 | 5.93% | 9.17% | 3.20% | 31 |
| All Trials | Age 75+ years | 0.313 | 0.43 | No | 1.11 | 0.98 | 1.27 | 0.104 | 20581 | 4769 | 13744 | 427 | 1641 | 8.22% | 10.67% | 2.40% | 41 |
| All Trials | Male | 0.314 | 0.48 | No | 1.12 | 0.96 | 1.31 | 0.157 | 20386 | 4087 | 14447 | 282 | 1570 | 6.45% | 9.80% | 3.30% | 30 |
| All Trials | Female | 0.548 | 0.18 | No | 1.06 | 0.93 | 1.22 | 0.365 | 20478 | 4572 | 13965 | 390 | 1551 | 7.86% | 10.00% | 2.10% | 47 |
| All Trials | White Race | 0.434 | 0.43 | No | 1.11 | 0.99 | 1.24 | 0.072 | 34052 | 7308 | 23524 | 568 | 2652 | 7.21% | 10.13% | 2.90% | 34 |
| All Trials | Non-White Race | 0.925 | 0.00 | No | 0.99 | 0.76 | 1.28 | 0.925 | 6812 | 1351 | 4888 | 104 | 469 | 7.15% | 8.75% | 1.60% | 62 |
| Trials 0 & 1 | Overall | 0.323 | 0.14 | No | 1.06 | 0.95 | 1.18 | 0.323 | 38802 | 7625 | 27561 | 598 | 3018 | 7.27% | 9.87% | 2.60% | 39 |
| Trials 0 & 1 | Time stratified: 1-30 days | 0.890 | 0.00 | No | 0.96 | 0.83 | 1.11 | 0.594 | 38802 | 7884 | 29351 | 339 | 1228 | 4.12% | 4.02% | -0.10% |  |
| Trials 0 & 1 | Time stratified: 31-91 days | 0.941 | 0.00 | No | 0.99 | 0.82 | 1.20 | 0.941 | 30219 | 5206 | 23779 | 194 | 1040 | 3.59% | 4.19% | 0.60% | 167 |
| Trials 0 & 1 | Time stratified: 92-182 days | 0.005 | 1.00 | Yes | 1.60 | 1.19 | 2.15 | 0.002 | 19890 | 2145 | 16930 | 65 | 750 | 2.94% | 4.24% | 1.30% | 77 |
| Trials 0 & 1 | Age 65-74 years | 0.587 | 0.19 | No | 1.06 | 0.89 | 1.27 | 0.489 | 19348 | 3439 | 14254 | 220 | 1435 | 6.01% | 9.15% | 3.10% | 32 |
| Trials 0 & 1 | Age 75+ years | 1.173 | 0.16 | No | 1.06 | 0.93 | 1.22 | 0.391 | 19454 | 4186 | 13307 | 378 | 1583 | 8.28% | 10.63% | 2.30% | 43 |
| Trials 0 & 1 | Male | 0.507 | 0.15 | No | 1.06 | 0.90 | 1.25 | 0.507 | 19437 | 3599 | 14063 | 257 | 1518 | 6.66% | 9.74% | 3.10% | 32 |
| Trials 0 & 1 | Female | 0.899 | 0.14 | No | 1.06 | 0.92 | 1.22 | 0.449 | 19365 | 4026 | 13498 | 341 | 1500 | 7.81% | 10.00% | 2.20% | 46 |
| Trials 0 & 1 | White Race | 0.940 | 0.33 | No | 1.09 | 0.97 | 1.22 | 0.157 | 32454 | 6471 | 22909 | 504 | 2570 | 7.23% | 10.09% | 2.90% | 35 |
| Trials 0 & 1 | Non-White Race | 0.679 | 0.00 | No | 0.90 | 0.68 | 1.18 | 0.453 | 6348 | 1154 | 4652 | 94 | 448 | 7.53% | 8.78% | 1.30% | 80 |
| Notes: A1 = Atorvastatin initiation; A2 = Newly initiated a different medication other than atorvastatin; N = Total number of patients in the analytic stratum; A = Number of A2 patients without the event; B = Number of A1 patients without the event; C = Number of A2 patients with the event; D = Number of A1 patients with the event; Risk A1 = Cumulative incidence (risk) in the A1 (atorvastatin) group; Risk A2 = Cumulative incidence (risk) in the A2 (comparator) group; RD = Risk difference; NNH = Number needed to harm. ADE Signal Confirmation metrics: q = Within-outcome Benjamini-Hochberg false-discovery-rate adjusted p-value; P(adj>1) = Proportion of 5,000 Monte Carlo draws from the probabilistic quantitative bias analysis in which the residual-confounding-adjusted subdistribution hazard ratio remained greater than 1. Confirmed = 1 if q ≤ 0.05 and P(adj>1) = 1; otherwise 0. Only analyses with at least 20 events in both treatment arms (C ≥ 20 and D ≥ 20) were included in the ADE Signal Confirmation analyses. Analyses that did not meet this threshold were excluded from both the Benjamini-Hochberg correction and the probabilistic quantitative bias analysis (denoted by †). * = Not reportable due to cell counts ≤ 10 per CMS policies. |  |  |  |  |  |  |  |  |  |  |  |  |  |  |  |  |  |

**Table S7 (Continued) Detailed Results for Confirmed ADE Signals**
| Detailed Results for Confirmed ADE Signals |  |  |  |  |  |  |  |  |  |  |  |  |  |  |  |  |  |
| --- | --- | --- | --- | --- | --- | --- | --- | --- | --- | --- | --- | --- | --- | --- | --- | --- | --- |
| Category : Cardiac valve events |  |  |  |  |  |  |  |  |  |  |  |  |  |  |  |  |  |
| Outcome : Cardiac valve intervention procedure |  |  |  |  |  |  |  |  |  |  |  |  |  |  |  |  |  |
| Analysis Set | Stratum | ADE Signal Confirmation |  |  | Weighted Fine-Gray |  |  |  | Unadjusted Descriptive Analyses for All Trials |  |  |  |  |  |  |  |  |
|  |  | q | P(adj>1) | Confirmed | sHR | LCL | UCL | p | N | A | B | C | D | Risk A2 | Risk A1 | RD | NNH |
| All Trials | Overall | 0.016 | 1.00 | Yes | 1.83 | 1.12 | 2.98 | 0.016 | 40902 | 9332 | 31382 | 24 | 164 | 0.26% | 0.52% | 0.30% | 380 |
| All Trials | Time stratified: 1-30 days | † | † | † | 1.93 | 1.00 | 3.74 | 0.051 | 40902 | 9344 | 31495 | 12 | 51 | 0.13% | 0.16% | 0.00% | 2993 |
| All Trials | Time stratified: 31-91 days | † | † | † | 3.29 | 1.13 | 9.62 | 0.029 | 33204 | 6491 | 26644 | * | 63 | * | 0.24% | * | * |
| All Trials | Time stratified: 92-182 days | † | † | † | 1.43 | 0.54 | 3.77 | 0.471 | 22577 | 2752 | 19769 | * | 50 | * | 0.25% | * | * |
| All Trials | Age 65-74 years | † | † | † | 2.25 | 0.97 | 5.21 | 0.058 | 20300 | 4139 | 16072 | * | 82 | * | 0.51% | * | * |
| All Trials | Age 75+ years | † | † | † | 1.60 | 0.88 | 2.92 | 0.124 | 20602 | 5193 | 15310 | 17 | 82 | 0.33% | 0.53% | 0.20% | 484 |
| All Trials | Male | † | † | † | 2.24 | 1.08 | 4.64 | 0.031 | 20400 | 4364 | 15931 | * | 95 | * | 0.59% | * | * |
| All Trials | Female | † | † | † | 1.48 | 0.77 | 2.87 | 0.241 | 20502 | 4968 | 15451 | 14 | 69 | 0.28% | 0.44% | 0.20% | 611 |
| All Trials | White Race | † | † | † | 2.23 | 1.30 | 3.84 | 0.004 | 34087 | 7882 | 26039 | 18 | 148 | 0.23% | 0.57% | 0.30% | 296 |
| All Trials | Non-White Race | † | † | † | 0.74 | 0.25 | 2.22 | 0.591 | 6815 | 1450 | 5343 | * | 16 | * | 0.30% | * | * |
| Trials 0 & 1 | Overall | 0.034 | 1.00 | Yes | 1.74 | 1.04 | 2.89 | 0.034 | 38807 | 8203 | 30422 | 22 | 160 | 0.27% | 0.52% | 0.30% | 391 |
| Trials 0 & 1 | Time stratified: 1-30 days | † | † | † | 1.82 | 0.92 | 3.62 | 0.086 | 38807 | 8214 | 30531 | 11 | 51 | 0.13% | 0.17% | 0.00% | 3028 |
| Trials 0 & 1 | Time stratified: 31-91 days | † | † | † | 3.69 | 1.06 | 12.86 | 0.040 | 31536 | 5655 | 25817 | * | 59 | * | 0.23% | * | * |
| Trials 0 & 1 | Time stratified: 92-182 days | † | † | † | 1.33 | 0.50 | 3.49 | 0.569 | 21696 | 2445 | 19195 | * | 50 | * | 0.26% | * | * |
| Trials 0 & 1 | Age 65-74 years | † | † | † | 2.28 | 0.92 | 5.65 | 0.074 | 19349 | 3653 | 15610 | * | 80 | * | 0.51% | * | * |
| Trials 0 & 1 | Age 75+ years | † | † | † | 1.47 | 0.79 | 2.73 | 0.222 | 19458 | 4550 | 14812 | 16 | 80 | 0.35% | 0.54% | 0.20% | 535 |
| Trials 0 & 1 | Male | † | † | † | 2.07 | 0.97 | 4.44 | 0.060 | 19440 | 3847 | 15492 | * | 92 | * | 0.59% | * | * |
| Trials 0 & 1 | Female | † | † | † | 1.44 | 0.72 | 2.88 | 0.296 | 19367 | 4356 | 14930 | 13 | 68 | 0.30% | 0.45% | 0.20% | 642 |
| Trials 0 & 1 | White Race | † | † | † | 2.05 | 1.17 | 3.58 | 0.012 | 32458 | 6959 | 25337 | 17 | 145 | 0.24% | 0.57% | 0.30% | 307 |
| Trials 0 & 1 | Non-White Race | † | † | † | 0.77 | 0.23 | 2.61 | 0.677 | 6349 | 1244 | 5085 | * | 15 | * | 0.29% | * | * |
| Notes: A1 = Atorvastatin initiation; A2 = Newly initiated a different medication other than atorvastatin; N = Total number of patients in the analytic stratum; A = Number of A2 patients without the event; B = Number of A1 patients without the event; C = Number of A2 patients with the event; D = Number of A1 patients with the event; Risk A1 = Cumulative incidence (risk) in the A1 (atorvastatin) group; Risk A2 = Cumulative incidence (risk) in the A2 (comparator) group; RD = Risk difference; NNH = Number needed to harm. ADE Signal Confirmation metrics: q = Within-outcome Benjamini-Hochberg false-discovery-rate adjusted p-value; P(adj>1) = Proportion of 5,000 Monte Carlo draws from the probabilistic quantitative bias analysis in which the residual-confounding-adjusted subdistribution hazard ratio remained greater than 1. Confirmed = 1 if q ≤ 0.05 and P(adj>1) = 1; otherwise 0. Only analyses with at least 20 events in both treatment arms (C ≥ 20 and D ≥ 20) were included in the ADE Signal Confirmation analyses. Analyses that did not meet this threshold were excluded from both the Benjamini-Hochberg correction and the probabilistic quantitative bias analysis (denoted by †). * = Not reportable due to cell counts ≤ 10 per CMS policies. |  |  |  |  |  |  |  |  |  |  |  |  |  |  |  |  |  |

**Table S7 (Continued) Detailed Results for Confirmed ADE Signals**
| Detailed Results for Confirmed ADE Signals |  |  |  |  |  |  |  |  |  |  |  |  |  |  |  |  |  |
| --- | --- | --- | --- | --- | --- | --- | --- | --- | --- | --- | --- | --- | --- | --- | --- | --- | --- |
| Category : Cardiac valve events |  |  |  |  |  |  |  |  |  |  |  |  |  |  |  |  |  |
| Outcome : Composite: valve disorder or intervention |  |  |  |  |  |  |  |  |  |  |  |  |  |  |  |  |  |
| Analysis Set | Stratum | ADE Signal Confirmation |  |  | Weighted Fine-Gray |  |  |  | Unadjusted Descriptive Analyses for All Trials |  |  |  |  |  |  |  |  |
|  |  | q | P(adj>1) | Confirmed | sHR | LCL | UCL | p | N | A | B | C | D | Risk A2 | Risk A1 | RD | NNH |
| All Trials | Overall | 0.149 | 0.26 | No | 1.08 | 0.97 | 1.19 | 0.149 | 40862 | 8614 | 28295 | 715 | 3238 | 7.66% | 10.27% | 2.60% | 38 |
| All Trials | Time stratified: 1-30 days | 0.882 | 0.00 | No | 0.99 | 0.86 | 1.13 | 0.882 | 40862 | 8921 | 30210 | 408 | 1323 | 4.37% | 4.20% | -0.20% |  |
| All Trials | Time stratified: 31-91 days | 1.010 | 0.06 | No | 1.04 | 0.87 | 1.24 | 0.674 | 31720 | 5940 | 24443 | 223 | 1114 | 3.62% | 4.36% | 0.70% | 135 |
| All Trials | Time stratified: 92-182 days | 0.021 | 1.00 | Yes | 1.44 | 1.11 | 1.87 | 0.007 | 20609 | 2391 | 17333 | 84 | 801 | 3.39% | 4.42% | 1.00% | 98 |
| All Trials | Age 65-74 years | 0.665 | 0.11 | No | 1.05 | 0.89 | 1.24 | 0.554 | 20283 | 3874 | 14624 | 261 | 1524 | 6.31% | 9.44% | 3.10% | 32 |
| All Trials | Age 75+ years | 0.631 | 0.43 | No | 1.11 | 0.98 | 1.26 | 0.105 | 20579 | 4740 | 13671 | 454 | 1714 | 8.74% | 11.14% | 2.40% | 42 |
| All Trials | Male | 0.576 | 0.22 | No | 1.07 | 0.92 | 1.24 | 0.384 | 20386 | 4062 | 14409 | 307 | 1608 | 7.03% | 10.04% | 3.00% | 33 |
| All Trials | Female | 0.456 | 0.30 | No | 1.08 | 0.95 | 1.24 | 0.228 | 20476 | 4552 | 13886 | 408 | 1630 | 8.23% | 10.51% | 2.30% | 44 |
| All Trials | White Race | 0.347 | 0.32 | No | 1.09 | 0.98 | 1.21 | 0.116 | 34051 | 7268 | 23428 | 607 | 2748 | 7.71% | 10.50% | 2.80% | 36 |
| All Trials | Non-White Race | 0.997 | 0.00 | No | 1.00 | 0.77 | 1.29 | 0.997 | 6811 | 1346 | 4867 | 108 | 490 | 7.43% | 9.15% | 1.70% | 58 |
| Trials 0 & 1 | Overall | 0.399 | 0.09 | No | 1.05 | 0.94 | 1.16 | 0.399 | 38802 | 7589 | 27450 | 634 | 3129 | 7.71% | 10.23% | 2.50% | 40 |
| Trials 0 & 1 | Time stratified: 1-30 days | 0.836 | 0.00 | No | 0.96 | 0.83 | 1.10 | 0.557 | 38802 | 7862 | 29296 | 361 | 1283 | 4.39% | 4.20% | -0.20% |  |
| Trials 0 & 1 | Time stratified: 31-91 days | 0.899 | 0.00 | No | 0.99 | 0.82 | 1.19 | 0.899 | 30154 | 5181 | 23698 | 202 | 1073 | 3.75% | 4.33% | 0.60% | 173 |
| Trials 0 & 1 | Time stratified: 92-182 days | 0.010 | 1.00 | Yes | 1.52 | 1.15 | 2.01 | 0.003 | 19820 | 2129 | 16847 | 71 | 773 | 3.23% | 4.39% | 1.20% | 86 |
| Trials 0 & 1 | Age 65-74 years | 0.764 | 0.08 | No | 1.04 | 0.88 | 1.24 | 0.636 | 19348 | 3426 | 14212 | 233 | 1477 | 6.37% | 9.41% | 3.00% | 33 |
| Trials 0 & 1 | Age 75+ years | 0.762 | 0.17 | No | 1.06 | 0.93 | 1.21 | 0.381 | 19454 | 4163 | 13238 | 401 | 1652 | 8.79% | 11.09% | 2.30% | 43 |
| Trials 0 & 1 | Male | 0.847 | 0.00 | No | 1.02 | 0.87 | 1.19 | 0.847 | 19437 | 3579 | 14026 | 277 | 1555 | 7.18% | 9.98% | 2.80% | 36 |
| Trials 0 & 1 | Female | 0.922 | 0.25 | No | 1.08 | 0.94 | 1.24 | 0.307 | 19365 | 4010 | 13424 | 357 | 1574 | 8.17% | 10.49% | 2.30% | 43 |
| Trials 0 & 1 | White Race | 1.318 | 0.23 | No | 1.07 | 0.96 | 1.20 | 0.220 | 32454 | 6439 | 22817 | 536 | 2662 | 7.68% | 10.45% | 2.80% | 36 |
| Trials 0 & 1 | Non-White Race | 0.739 | 0.00 | No | 0.91 | 0.70 | 1.19 | 0.493 | 6348 | 1150 | 4633 | 98 | 467 | 7.85% | 9.16% | 1.30% | 77 |
| Notes: A1 = Atorvastatin initiation; A2 = Newly initiated a different medication other than atorvastatin; N = Total number of patients in the analytic stratum; A = Number of A2 patients without the event; B = Number of A1 patients without the event; C = Number of A2 patients with the event; D = Number of A1 patients with the event; Risk A1 = Cumulative incidence (risk) in the A1 (atorvastatin) group; Risk A2 = Cumulative incidence (risk) in the A2 (comparator) group; RD = Risk difference; NNH = Number needed to harm. ADE Signal Confirmation metrics: q = Within-outcome Benjamini-Hochberg false-discovery-rate adjusted p-value; P(adj>1) = Proportion of 5,000 Monte Carlo draws from the probabilistic quantitative bias analysis in which the residual-confounding-adjusted subdistribution hazard ratio remained greater than 1. Confirmed = 1 if q ≤ 0.05 and P(adj>1) = 1; otherwise 0. Only analyses with at least 20 events in both treatment arms (C ≥ 20 and D ≥ 20) were included in the ADE Signal Confirmation analyses. Analyses that did not meet this threshold were excluded from both the Benjamini-Hochberg correction and the probabilistic quantitative bias analysis (denoted by †). * = Not reportable due to cell counts ≤ 10 per CMS policies. |  |  |  |  |  |  |  |  |  |  |  |  |  |  |  |  |  |

**Table S7 (Continued) Detailed Results for Confirmed ADE Signals**
| Detailed Results for Confirmed ADE Signals |  |  |  |  |  |  |  |  |  |  |  |  |  |  |  |  |  |
| --- | --- | --- | --- | --- | --- | --- | --- | --- | --- | --- | --- | --- | --- | --- | --- | --- | --- |
| Category : Sprains, strains, and tendon injury |  |  |  |  |  |  |  |  |  |  |  |  |  |  |  |  |  |
| Outcome : Sprains and strains, initial encounter |  |  |  |  |  |  |  |  |  |  |  |  |  |  |  |  |  |
| Analysis Set | Stratum | ADE Signal Confirmation |  |  | Weighted Fine-Gray |  |  |  | Unadjusted Descriptive Analyses for All Trials |  |  |  |  |  |  |  |  |
|  |  | q | P(adj>1) | Confirmed | sHR | LCL | UCL | p | N | A | B | C | D | Risk A2 | Risk A1 | RD | NNH |
| All Trials | Overall | 0.039 | 0.87 | No | 1.24 | 1.01 | 1.53 | 0.039 | 50319 | 12459 | 36959 | 147 | 754 | 1.17% | 2.00% | 0.80% | 120 |
| All Trials | Time stratified: 1-30 days | 0.576 | 0.69 | No | 1.18 | 0.82 | 1.69 | 0.384 | 50319 | 12555 | 37539 | 51 | 174 | 0.40% | 0.46% | 0.10% | 1760 |
| All Trials | Time stratified: 31-91 days | 0.431 | 0.96 | No | 1.30 | 0.91 | 1.86 | 0.144 | 40712 | 8848 | 31530 | 51 | 283 | 0.57% | 0.89% | 0.30% | 316 |
| All Trials | Time stratified: 92-182 days | 0.559 | 0.43 | No | 1.11 | 0.78 | 1.58 | 0.559 | 26881 | 3648 | 22891 | 45 | 297 | 1.22% | 1.28% | 0.10% | 1605 |
| All Trials | Age 65-74 years | 0.160 | 0.95 | No | 1.30 | 0.95 | 1.77 | 0.096 | 23192 | 4987 | 17770 | 57 | 378 | 1.13% | 2.08% | 1.00% | 105 |
| All Trials | Age 75+ years | 0.259 | 0.76 | No | 1.20 | 0.91 | 1.58 | 0.207 | 27127 | 7472 | 19189 | 90 | 376 | 1.19% | 1.92% | 0.70% | 137 |
| **All Trials** | **Male** | **0.014** | **1.00** | **Yes** | **1.72** | **1.20** | **2.45** | **0.003** | **24448** | **5699** | **18351** | **47** | **351** | **0.82%** | **1.88%** | **1.10%** | **94** |
| All Trials | Female | 0.726 | 0.10 | No | 1.05 | 0.81 | 1.35 | 0.726 | 25871 | 6760 | 18608 | 100 | 403 | 1.46% | 2.12% | 0.70% | 151 |
| All Trials | White Race | 0.234 | 0.77 | No | 1.20 | 0.97 | 1.49 | 0.093 | 42014 | 10563 | 30681 | 131 | 639 | 1.22% | 2.04% | 0.80% | 123 |
| All Trials | Non-White Race | † | † | † | 1.51 | 0.78 | 2.93 | 0.225 | 8305 | 1896 | 6278 | 16 | 115 | 0.84% | 1.80% | 1.00% | 104 |
| Trials 0 & 1 | Overall | 0.034 | 0.91 | No | 1.27 | 1.02 | 1.58 | 0.034 | 47463 | 10874 | 35727 | 127 | 735 | 1.15% | 2.02% | 0.90% | 116 |
| Trials 0 & 1 | Time stratified: 1-30 days | 0.414 | 0.68 | No | 1.17 | 0.80 | 1.71 | 0.414 | 47463 | 10955 | 36291 | 46 | 171 | 0.42% | 0.47% | 0.10% | 1967 |
| Trials 0 & 1 | Time stratified: 31-91 days | 0.654 | 0.91 | No | 1.27 | 0.87 | 1.86 | 0.218 | 38439 | 7652 | 30470 | 45 | 272 | 0.58% | 0.88% | 0.30% | 333 |
| Trials 0 & 1 | Time stratified: 92-182 days | 0.398 | 0.88 | No | 1.25 | 0.85 | 1.84 | 0.265 | 25733 | 3234 | 22171 | 36 | 292 | 1.10% | 1.30% | 0.20% | 503 |
| Trials 0 & 1 | Age 65-74 years | 0.189 | 0.90 | No | 1.27 | 0.92 | 1.75 | 0.151 | 21993 | 4352 | 17217 | 52 | 372 | 1.18% | 2.11% | 0.90% | 107 |
| Trials 0 & 1 | Age 75+ years | 0.216 | 0.90 | No | 1.26 | 0.93 | 1.71 | 0.129 | 25470 | 6522 | 18510 | 75 | 363 | 1.14% | 1.92% | 0.80% | 127 |
| **Trials 0 & 1** | **Male** | **0.026** | **1.00** | **Yes** | **1.73** | **1.18** | **2.54** | **0.005** | **23143** | **4975** | **17786** | **41** | **341** | **0.82%** | **1.88%** | **1.10%** | **94** |
| Trials 0 & 1 | Female | 0.592 | 0.26 | No | 1.08 | 0.82 | 1.41 | 0.592 | 24320 | 5899 | 17941 | 86 | 394 | 1.44% | 2.15% | 0.70% | 140 |
| Trials 0 & 1 | White Race | 0.181 | 0.84 | No | 1.23 | 0.98 | 1.55 | 0.072 | 39786 | 9263 | 29785 | 114 | 624 | 1.22% | 2.05% | 0.80% | 120 |
| Trials 0 & 1 | Non-White Race | † | † | † | 1.50 | 0.73 | 3.11 | 0.272 | 7677 | 1611 | 5942 | 13 | 111 | 0.80% | 1.83% | 1.00% | 97 |

**Table S7 (Continued) Detailed Results for Confirmed ADE Signals**
| Detailed Results for Confirmed ADE Signals |  |  |  |  |  |  |  |  |  |  |  |  |  |  |  |  |  |
| --- | --- | --- | --- | --- | --- | --- | --- | --- | --- | --- | --- | --- | --- | --- | --- | --- | --- |
| Category : Sprains, strains, and tendon injury |  |  |  |  |  |  |  |  |  |  |  |  |  |  |  |  |  |
| Outcome : Sprain |  |  |  |  |  |  |  |  |  |  |  |  |  |  |  |  |  |
| Analysis Set | Stratum | ADE Signal Confirmation |  |  | Weighted Fine-Gray |  |  |  | Unadjusted Descriptive Analyses for All Trials |  |  |  |  |  |  |  |  |
|  |  | q | P(adj>1) | Confirmed | sHR | LCL | UCL | p | N | A | B | C | D | Risk A2 | Risk A1 | RD | NNH |
| All Trials | Overall | 0.059 | 0.96 | No | 1.31 | 0.99 | 1.73 | 0.059 | 50321 | 12533 | 37330 | 75 | 383 | 0.59% | 1.02% | 0.40% | 238 |
| All Trials | Time stratified: 1-30 days | 0.552 | 0.97 | No | 1.32 | 0.80 | 2.17 | 0.276 | 50321 | 12581 | 37622 | 27 | 91 | 0.21% | 0.24% | 0.00% | 3684 |
| All Trials | Time stratified: 31-91 days | 0.525 | 0.61 | No | 1.15 | 0.75 | 1.77 | 0.525 | 40810 | 8894 | 31750 | 30 | 136 | 0.34% | 0.43% | 0.10% | 1107 |
| All Trials | Time stratified: 92-182 days | † | † | † | 1.37 | 0.80 | 2.34 | 0.252 | 27098 | 3698 | 23226 | 18 | 156 | 0.48% | 0.67% | 0.20% | 547 |
| All Trials | Age 65-74 years | 0.244 | 1.00 | No | 1.49 | 0.93 | 2.38 | 0.098 | 23194 | 5020 | 17952 | 26 | 196 | 0.52% | 1.08% | 0.60% | 177 |
| All Trials | Age 75+ years | 0.452 | 0.70 | No | 1.18 | 0.83 | 1.66 | 0.361 | 27127 | 7513 | 19378 | 49 | 187 | 0.65% | 0.96% | 0.30% | 325 |
| All Trials | Male | 0.028 | 1.00 | Yes | 1.94 | 1.21 | 3.09 | 0.006 | 24450 | 5724 | 18520 | 24 | 182 | 0.42% | 0.97% | 0.60% | 180 |
| All Trials | Female | 0.723 | 0.18 | No | 1.07 | 0.75 | 1.51 | 0.723 | 25871 | 6809 | 18810 | 51 | 201 | 0.74% | 1.06% | 0.30% | 319 |
| All Trials | White Race | 0.445 | 0.70 | No | 1.18 | 0.88 | 1.58 | 0.267 | 42015 | 10626 | 30995 | 69 | 325 | 0.65% | 1.04% | 0.40% | 255 |
| All Trials | Non-White Race | † | † | † | 2.94 | 1.21 | 7.15 | 0.018 | 8306 | 1907 | 6335 | * | 58 | * | 0.91% | * | * |
| Trials 0 & 1 | Overall | 0.067 | 0.97 | No | 1.32 | 0.98 | 1.78 | 0.067 | 47464 | 10936 | 36090 | 66 | 372 | 0.60% | 1.02% | 0.40% | 238 |
| Trials 0 & 1 | Time stratified: 1-30 days | 0.993 | 0.74 | No | 1.19 | 0.72 | 1.98 | 0.496 | 47464 | 10976 | 36373 | 26 | 89 | 0.24% | 0.24% | 0.00% | 12872 |
| Trials 0 & 1 | Time stratified: 31-91 days | 0.527 | 0.65 | No | 1.16 | 0.73 | 1.85 | 0.527 | 38532 | 7692 | 30684 | 26 | 130 | 0.34% | 0.42% | 0.10% | 1176 |
| Trials 0 & 1 | Time stratified: 92-182 days | † | † | † | 1.61 | 0.87 | 2.96 | 0.126 | 25942 | 3276 | 22499 | 14 | 153 | 0.43% | 0.68% | 0.20% | 400 |
| Trials 0 & 1 | Age 65-74 years | 0.641 | 1.00 | No | 1.42 | 0.87 | 2.30 | 0.160 | 21994 | 4381 | 17395 | 24 | 194 | 0.54% | 1.10% | 0.60% | 179 |
| Trials 0 & 1 | Age 75+ years | 0.368 | 0.85 | No | 1.23 | 0.85 | 1.79 | 0.276 | 25470 | 6555 | 18695 | 42 | 178 | 0.64% | 0.94% | 0.30% | 326 |
| Trials 0 & 1 | Male | † | † | † | 2.28 | 1.34 | 3.85 | 0.002 | 23144 | 4998 | 17951 | 19 | 176 | 0.38% | 0.97% | 0.60% | 169 |
| Trials 0 & 1 | Female | 0.908 | 0.01 | No | 1.02 | 0.71 | 1.47 | 0.908 | 24320 | 5938 | 18139 | 47 | 196 | 0.79% | 1.07% | 0.30% | 352 |
| Trials 0 & 1 | White Race | 0.539 | 0.73 | No | 1.19 | 0.87 | 1.63 | 0.270 | 39786 | 9316 | 30092 | 61 | 317 | 0.65% | 1.04% | 0.40% | 255 |
| Trials 0 & 1 | Non-White Race | † | † | † | 3.05 | 1.15 | 8.06 | 0.024 | 7678 | 1620 | 5998 | * | 55 | * | 0.91% | * | * |
| Notes: A1 = Atorvastatin initiation; A2 = Newly initiated a different medication other than atorvastatin; N = Total number of patients in the analytic stratum; A = Number of A2 patients without the event; B = Number of A1 patients without the event; C = Number of A2 patients with the event; D = Number of A1 patients with the event; Risk A1 = Cumulative incidence (risk) in the A1 (atorvastatin) group; Risk A2 = Cumulative incidence (risk) in the A2 (comparator) group; RD = Risk difference; NNH = Number needed to harm. ADE Signal Confirmation metrics: q = Within-outcome Benjamini-Hochberg false-discovery-rate adjusted p-value; P(adj>1) = Proportion of 5,000 Monte Carlo draws from the probabilistic quantitative bias analysis in which the residual-confounding-adjusted subdistribution hazard ratio remained greater than 1. Confirmed = 1 if q ≤ 0.05 and P(adj>1) = 1; otherwise 0. Only analyses with at least 20 events in both treatment arms (C ≥ 20 and D ≥ 20) were included in the ADE Signal Confirmation analyses. Analyses that did not meet this threshold were excluded from both the Benjamini-Hochberg correction and the probabilistic quantitative bias analysis (denoted by †). * = Not reportable due to cell counts ≤ 10 per CMS policies. |  |  |  |  |  |  |  |  |  |  |  |  |  |  |  |  |  |

**Table S7 (Continued) Detailed Results for Confirmed ADE Signals**
| Detailed Results for Confirmed ADE Signals |  |  |  |  |  |  |  |  |  |  |  |  |  |  |  |  |  |
| --- | --- | --- | --- | --- | --- | --- | --- | --- | --- | --- | --- | --- | --- | --- | --- | --- | --- |
| Category : Sprains, strains, and tendon injury |  |  |  |  |  |  |  |  |  |  |  |  |  |  |  |  |  |
| Outcome : Composite: sprain/strain/tendon injury |  |  |  |  |  |  |  |  |  |  |  |  |  |  |  |  |  |
| Analysis Set | Stratum | ADE Signal Confirmation |  |  | Weighted Fine-Gray |  |  |  | Unadjusted Descriptive Analyses for All Trials |  |  |  |  |  |  |  |  |
|  |  | q | P(adj>1) | Confirmed | sHR | LCL | UCL | p | N | A | B | C | D | Risk A2 | Risk A1 | RD | NNH |
| All Trials | Overall | 0.043 | 0.76 | No | 1.20 | 1.01 | 1.43 | 0.043 | 50317 | 12397 | 36656 | 208 | 1056 | 1.65% | 2.80% | 1.20% | 87 |
| All Trials | Time stratified: 1-30 days | 0.459 | 0.50 | No | 1.12 | 0.82 | 1.54 | 0.459 | 50317 | 12533 | 37467 | 72 | 245 | 0.57% | 0.65% | 0.10% | 1275 |
| All Trials | Time stratified: 31-91 days | 0.620 | 0.79 | No | 1.21 | 0.90 | 1.62 | 0.207 | 40628 | 8802 | 31354 | 78 | 394 | 0.88% | 1.24% | 0.40% | 276 |
| All Trials | Time stratified: 92-182 days | 0.647 | 0.54 | No | 1.13 | 0.83 | 1.55 | 0.431 | 26713 | 3608 | 22630 | 58 | 417 | 1.58% | 1.81% | 0.20% | 440 |
| All Trials | Age 65-74 years | 0.267 | 0.84 | No | 1.23 | 0.94 | 1.61 | 0.134 | 23191 | 4966 | 17625 | 78 | 522 | 1.55% | 2.88% | 1.30% | 75 |
| All Trials | Age 75+ years | 0.276 | 0.68 | No | 1.17 | 0.93 | 1.48 | 0.184 | 27126 | 7431 | 19031 | 130 | 534 | 1.72% | 2.73% | 1.00% | 99 |
| All Trials | Male | 0.003 | 1.00 | Yes | 1.66 | 1.24 | 2.21 | 0.001 | 24446 | 5674 | 18207 | 71 | 494 | 1.24% | 2.64% | 1.40% | 71 |
| All Trials | Female | 0.962 | 0.00 | No | 1.01 | 0.81 | 1.26 | 0.962 | 25871 | 6723 | 18449 | 137 | 562 | 2.00% | 2.96% | 1.00% | 104 |
| All Trials | White Race | 0.294 | 0.67 | No | 1.17 | 0.97 | 1.40 | 0.098 | 42012 | 10505 | 30422 | 188 | 897 | 1.76% | 2.86% | 1.10% | 90 |
| All Trials | Non-White Race | 0.283 | 1.00 | No | 1.41 | 0.80 | 2.50 | 0.235 | 8305 | 1892 | 6234 | 20 | 159 | 1.05% | 2.49% | 1.40% | 69 |
| Trials 0 & 1 | Overall | 0.029 | 0.85 | No | 1.24 | 1.02 | 1.50 | 0.029 | 47463 | 10823 | 35432 | 178 | 1030 | 1.62% | 2.82% | 1.20% | 83 |
| Trials 0 & 1 | Time stratified: 1-30 days | 0.616 | 0.30 | No | 1.09 | 0.79 | 1.50 | 0.616 | 47463 | 10935 | 36220 | 66 | 242 | 0.60% | 0.66% | 0.10% | 1568 |
| Trials 0 & 1 | Time stratified: 31-91 days | 0.517 | 0.89 | No | 1.25 | 0.91 | 1.74 | 0.172 | 38358 | 7615 | 30298 | 65 | 380 | 0.85% | 1.24% | 0.40% | 255 |
| Trials 0 & 1 | Time stratified: 92-182 days | 0.262 | 0.92 | No | 1.27 | 0.90 | 1.81 | 0.174 | 25573 | 3201 | 21917 | 47 | 408 | 1.45% | 1.83% | 0.40% | 263 |
| Trials 0 & 1 | Age 65-74 years | 0.197 | 0.84 | No | 1.23 | 0.92 | 1.63 | 0.158 | 21993 | 4335 | 17077 | 69 | 512 | 1.57% | 2.91% | 1.30% | 74 |
| Trials 0 & 1 | Age 75+ years | 0.179 | 0.85 | No | 1.23 | 0.96 | 1.60 | 0.108 | 25470 | 6488 | 18355 | 109 | 518 | 1.65% | 2.74% | 1.10% | 92 |
| Trials 0 & 1 | Male | 0.002 | 1.00 | Yes | 1.76 | 1.28 | 2.41 | 0.000 | 23143 | 4956 | 17646 | 60 | 481 | 1.20% | 2.65% | 1.50% | 69 |
| Trials 0 & 1 | Female | 0.831 | 0.02 | No | 1.03 | 0.81 | 1.30 | 0.831 | 24320 | 5867 | 17786 | 118 | 549 | 1.97% | 2.99% | 1.00% | 98 |
| Trials 0 & 1 | White Race | 0.194 | 0.75 | No | 1.19 | 0.98 | 1.45 | 0.078 | 39786 | 9214 | 29532 | 163 | 877 | 1.74% | 2.88% | 1.10% | 87 |
| Trials 0 & 1 | Non-White Race | † | † | † | 1.54 | 0.80 | 2.97 | 0.195 | 7677 | 1609 | 5900 | 15 | 153 | 0.92% | 2.53% | 1.60% | 62 |

**Table S7 (Continued) Detailed Results for Confirmed ADE Signals**
| Detailed Results for Confirmed ADE Signals |  |  |  |  |  |  |  |  |  |  |  |  |  |  |  |  |  |
| --- | --- | --- | --- | --- | --- | --- | --- | --- | --- | --- | --- | --- | --- | --- | --- | --- | --- |
| Category : Dizziness |  |  |  |  |  |  |  |  |  |  |  |  |  |  |  |  |  |
| Outcome : General sensation/perception signs and symptoms |  |  |  |  |  |  |  |  |  |  |  |  |  |  |  |  |  |
| Analysis Set | Stratum | ADE Signal Confirmation |  |  | Weighted Fine-Gray |  |  |  | Unadjusted Descriptive Analyses for All Trials |  |  |  |  |  |  |  |  |
|  |  | q | P(adj>1) | Confirmed | sHR | LCL | UCL | p | N | A | B | C | D | Risk A2 | Risk A1 | RD | NNH |
| All Trials | Overall | 0.000 | 0.86 | No | 1.24 | 1.12 | 1.37 | 0.000 | 46535 | 10866 | 31939 | 659 | 3071 | 5.72% | 8.77% | 3.10% | 33 |
| All Trials | Time stratified: 1-30 days | 0.021 | 0.82 | No | 1.22 | 1.06 | 1.42 | 0.007 | 46535 | 11179 | 33699 | 346 | 1311 | 3.00% | 3.74% | 0.70% | 135 |
| All Trials | Time stratified: 31-91 days | 0.021 | 0.89 | No | 1.26 | 1.05 | 1.51 | 0.014 | 36255 | 7604 | 27422 | 205 | 1024 | 2.63% | 3.60% | 1.00% | 103 |
| All Trials | Time stratified: 92-182 days | 0.083 | 0.81 | No | 1.22 | 0.97 | 1.53 | 0.083 | 23400 | 3074 | 19482 | 108 | 736 | 3.39% | 3.64% | 0.20% | 406 |
| All Trials | Age 65-74 years | 0.079 | 0.62 | No | 1.16 | 0.98 | 1.36 | 0.079 | 22060 | 4512 | 15845 | 269 | 1434 | 5.63% | 8.30% | 2.70% | 37 |
| All Trials | Age 75+ years | 0.000 | 0.97 | No | 1.32 | 1.16 | 1.50 | 0.000 | 24475 | 6354 | 16094 | 390 | 1637 | 5.78% | 9.23% | 3.40% | 29 |
| All Trials | Male | 0.018 | 0.83 | No | 1.22 | 1.05 | 1.43 | 0.012 | 23016 | 5076 | 16218 | 288 | 1434 | 5.37% | 8.12% | 2.80% | 36 |
| All Trials | Female | 0.001 | 0.91 | No | 1.26 | 1.10 | 1.45 | 0.001 | 23519 | 5790 | 15721 | 371 | 1637 | 6.02% | 9.43% | 3.40% | 29 |
| All Trials | White Race | 0.002 | 0.79 | No | 1.21 | 1.09 | 1.35 | 0.001 | 39003 | 9245 | 26659 | 567 | 2532 | 5.78% | 8.67% | 2.90% | 35 |
| All Trials | Non-White Race | 0.024 | 1.00 | Yes | 1.40 | 1.05 | 1.86 | 0.020 | 7532 | 1621 | 5280 | 92 | 539 | 5.37% | 9.26% | 3.90% | 26 |
| Trials 0 & 1 | Overall | 0.002 | 0.70 | No | 1.18 | 1.06 | 1.31 | 0.002 | 44160 | 9625 | 30965 | 599 | 2971 | 5.86% | 8.75% | 2.90% | 35 |
| Trials 0 & 1 | Time stratified: 1-30 days | 0.185 | 0.63 | No | 1.16 | 0.99 | 1.35 | 0.062 | 44160 | 9911 | 32666 | 313 | 1270 | 3.06% | 3.74% | 0.70% | 147 |
| Trials 0 & 1 | Time stratified: 31-91 days | 0.122 | 0.73 | No | 1.18 | 0.98 | 1.43 | 0.082 | 34430 | 6676 | 26581 | 186 | 987 | 2.71% | 3.58% | 0.90% | 115 |
| Trials 0 & 1 | Time stratified: 92-182 days | 0.140 | 0.73 | No | 1.19 | 0.94 | 1.51 | 0.140 | 22495 | 2755 | 18926 | 100 | 714 | 3.50% | 3.64% | 0.10% | 753 |
| Trials 0 & 1 | Age 65-74 years | 0.330 | 0.32 | No | 1.09 | 0.92 | 1.29 | 0.330 | 21030 | 3991 | 15404 | 245 | 1390 | 5.78% | 8.28% | 2.50% | 40 |
| Trials 0 & 1 | Age 75+ years | 0.003 | 0.92 | No | 1.27 | 1.11 | 1.45 | 0.001 | 23130 | 5634 | 15561 | 354 | 1581 | 5.91% | 9.22% | 3.30% | 30 |
| Trials 0 & 1 | Male | 0.108 | 0.62 | No | 1.15 | 0.98 | 1.35 | 0.090 | 21917 | 4486 | 15770 | 266 | 1395 | 5.60% | 8.13% | 2.50% | 40 |
| Trials 0 & 1 | Female | 0.021 | 0.82 | No | 1.21 | 1.05 | 1.40 | 0.007 | 22243 | 5139 | 15195 | 333 | 1576 | 6.09% | 9.40% | 3.30% | 30 |
| Trials 0 & 1 | White Race | 0.021 | 0.63 | No | 1.16 | 1.04 | 1.30 | 0.011 | 37142 | 8224 | 25940 | 516 | 2462 | 5.90% | 8.67% | 2.80% | 36 |
| Trials 0 & 1 | Non-White Race | 0.125 | 0.96 | No | 1.30 | 0.97 | 1.75 | 0.083 | 7018 | 1401 | 5025 | 83 | 509 | 5.59% | 9.20% | 3.60% | 28 |
| Notes: A1 = Atorvastatin initiation; A2 = Newly initiated a different medication other than atorvastatin; N = Total number of patients in the analytic stratum; A = Number of A2 patients without the event; B = Number of A1 patients without the event; C = Number of A2 patients with the event; D = Number of A1 patients with the event; Risk A1 = Cumulative incidence (risk) in the A1 (atorvastatin) group; Risk A2 = Cumulative incidence (risk) in the A2 (comparator) group; RD = Risk difference; NNH = Number needed to harm. ADE Signal Confirmation metrics: q = Within-outcome Benjamini-Hochberg false-discovery-rate adjusted p-value; P(adj>1) = Proportion of 5,000 Monte Carlo draws from the probabilistic quantitative bias analysis in which the residual-confounding-adjusted subdistribution hazard ratio remained greater than 1. Confirmed = 1 if q ≤ 0.05 and P(adj>1) = 1; otherwise 0. Only analyses with at least 20 events in both treatment arms (C ≥ 20 and D ≥ 20) were included in the ADE Signal Confirmation analyses. Analyses that did not meet this threshold were excluded from both the Benjamini-Hochberg correction and the probabilistic quantitative bias analysis (denoted by †). * = Not reportable due to cell counts ≤ 10 per CMS policies. |  |  |  |  |  |  |  |  |  |  |  |  |  |  |  |  |  |

**Table S7 (Continued) Detailed Results for Confirmed ADE Signals**
| Detailed Results for Confirmed ADE Signals |  |  |  |  |  |  |  |  |  |  |  |  |  |  |  |  |  |
| --- | --- | --- | --- | --- | --- | --- | --- | --- | --- | --- | --- | --- | --- | --- | --- | --- | --- |
| Category : Dizziness |  |  |  |  |  |  |  |  |  |  |  |  |  |  |  |  |  |
| Outcome : Dizziness and giddiness |  |  |  |  |  |  |  |  |  |  |  |  |  |  |  |  |  |
| Analysis Set | Stratum | ADE Signal Confirmation |  |  | Weighted Fine-Gray |  |  |  | Unadjusted Descriptive Analyses for All Trials |  |  |  |  |  |  |  |  |
|  |  | q | P(adj>1) | Confirmed | sHR | LCL | UCL | p | N | A | B | C | D | Risk A2 | Risk A1 | RD | NNH |
| All Trials | Overall | 0.000 | 0.90 | No | 1.26 | 1.14 | 1.40 | 0.000 | 46537 | 10915 | 32064 | 612 | 2946 | 5.31% | 8.41% | 3.10% | 32 |
| All Trials | Time stratified: 1-30 days | 0.020 | 0.85 | No | 1.24 | 1.06 | 1.44 | 0.007 | 46537 | 11203 | 33744 | 324 | 1266 | 2.81% | 3.62% | 0.80% | 124 |
| All Trials | Time stratified: 31-91 days | 0.014 | 0.94 | No | 1.29 | 1.06 | 1.56 | 0.009 | 36321 | 7642 | 27508 | 190 | 981 | 2.43% | 3.44% | 1.00% | 98 |
| All Trials | Time stratified: 92-182 days | 0.048 | 0.92 | No | 1.27 | 1.00 | 1.61 | 0.048 | 23477 | 3099 | 19581 | 98 | 699 | 3.07% | 3.45% | 0.40% | 262 |
| All Trials | Age 65-74 years | 0.097 | 0.62 | No | 1.15 | 0.97 | 1.36 | 0.097 | 22060 | 4523 | 15892 | 258 | 1387 | 5.40% | 8.03% | 2.60% | 38 |
| All Trials | Age 75+ years | 0.000 | 1.00 | No | 1.37 | 1.19 | 1.57 | 0.000 | 24477 | 6392 | 16172 | 354 | 1559 | 5.25% | 8.79% | 3.50% | 28 |
| All Trials | Male | 0.014 | 0.86 | No | 1.24 | 1.05 | 1.46 | 0.009 | 23017 | 5095 | 16261 | 270 | 1391 | 5.03% | 7.88% | 2.80% | 35 |
| All Trials | Female | 0.001 | 0.94 | No | 1.29 | 1.12 | 1.49 | 0.000 | 23520 | 5820 | 15803 | 342 | 1555 | 5.55% | 8.96% | 3.40% | 29 |
| All Trials | White Race | 0.001 | 0.84 | No | 1.23 | 1.10 | 1.38 | 0.000 | 39005 | 9288 | 26769 | 526 | 2422 | 5.36% | 8.30% | 2.90% | 34 |
| All Trials | Non-White Race | 0.021 | 1.00 | Yes | 1.43 | 1.06 | 1.91 | 0.017 | 7532 | 1627 | 5295 | 86 | 524 | 5.02% | 9.00% | 4.00% | 25 |
| Trials 0 & 1 | Overall | 0.001 | 0.75 | No | 1.20 | 1.07 | 1.34 | 0.001 | 44160 | 9665 | 31087 | 559 | 2849 | 5.47% | 8.40% | 2.90% | 34 |
| Trials 0 & 1 | Time stratified: 1-30 days | 0.097 | 0.65 | No | 1.16 | 0.99 | 1.36 | 0.064 | 44160 | 9929 | 32709 | 295 | 1227 | 2.89% | 3.62% | 0.70% | 137 |
| Trials 0 & 1 | Time stratified: 31-91 days | 0.185 | 0.79 | No | 1.21 | 0.99 | 1.47 | 0.062 | 34489 | 6707 | 26664 | 173 | 945 | 2.51% | 3.42% | 0.90% | 110 |
| Trials 0 & 1 | Time stratified: 92-182 days | 0.090 | 0.85 | No | 1.24 | 0.97 | 1.58 | 0.090 | 22567 | 2777 | 19022 | 91 | 677 | 3.17% | 3.44% | 0.30% | 379 |
| Trials 0 & 1 | Age 65-74 years | 0.394 | 0.27 | No | 1.08 | 0.91 | 1.28 | 0.394 | 21030 | 4001 | 15451 | 235 | 1343 | 5.55% | 8.00% | 2.40% | 41 |
| Trials 0 & 1 | Age 75+ years | 0.001 | 0.97 | No | 1.31 | 1.14 | 1.51 | 0.000 | 23130 | 5664 | 15636 | 324 | 1506 | 5.41% | 8.79% | 3.40% | 30 |
| Trials 0 & 1 | Male | 0.111 | 0.67 | No | 1.16 | 0.99 | 1.38 | 0.074 | 21917 | 4502 | 15812 | 250 | 1353 | 5.26% | 7.88% | 2.60% | 38 |
| Trials 0 & 1 | Female | 0.017 | 0.84 | No | 1.23 | 1.06 | 1.43 | 0.006 | 22243 | 5163 | 15275 | 309 | 1496 | 5.65% | 8.92% | 3.30% | 31 |
| Trials 0 & 1 | White Race | 0.015 | 0.69 | No | 1.17 | 1.04 | 1.32 | 0.008 | 37142 | 8259 | 26048 | 481 | 2354 | 5.50% | 8.29% | 2.80% | 36 |
| Trials 0 & 1 | Non-White Race | 0.089 | 0.97 | No | 1.32 | 0.97 | 1.80 | 0.074 | 7018 | 1406 | 5039 | 78 | 495 | 5.26% | 8.94% | 3.70% | 27 |

**Table S7 (Continued) Detailed Results for Confirmed ADE Signals**
| Detailed Results for Confirmed ADE Signals |  |  |  |  |  |  |  |  |  |  |  |  |  |  |  |  |  |
| --- | --- | --- | --- | --- | --- | --- | --- | --- | --- | --- | --- | --- | --- | --- | --- | --- | --- |
| Category : Abnormal symptoms |  |  |  |  |  |  |  |  |  |  |  |  |  |  |  |  |  |
| Outcome : Acute hepatic failure |  |  |  |  |  |  |  |  |  |  |  |  |  |  |  |  |  |
| Analysis Set | Stratum | ADE Signal Confirmation |  |  | Weighted Fine-Gray |  |  |  | Unadjusted Descriptive Analyses for All Trials |  |  |  |  |  |  |  |  |
|  |  | q | P(adj>1) | Confirmed | sHR | LCL | UCL | p | N | A | B | C | D | Risk A2 | Risk A1 | RD | NNH |
| All Trials | Overall | 0.007 | 1.00 | Yes | 1.72 | 1.16 | 2.55 | 0.007 | 49350 | 12049 | 37050 | 42 | 209 | 0.35% | 0.56% | 0.20% | 468 |
| All Trials | Time stratified: 1-30 days | 0.359 | 0.94 | No | 1.29 | 0.75 | 2.24 | 0.359 | 49350 | 12068 | 37187 | 23 | 72 | 0.19% | 0.19% | 0.00% | 33137 |
| All Trials | Time stratified: 31-91 days | † | † | † | 3.21 | 1.56 | 6.59 | 0.001 | 40066 | 8521 | 31457 | 11 | 77 | 0.13% | 0.24% | 0.10% | 868 |
| All Trials | Time stratified: 92-182 days | † | † | † | 1.14 | 0.49 | 2.63 | 0.760 | 26858 | 3592 | 23198 | * | 60 | * | 0.26% | * | * |
| All Trials | Age 65-74 years | † | † | † | 1.91 | 1.02 | 3.56 | 0.042 | 22822 | 4836 | 17882 | 15 | 89 | 0.31% | 0.50% | 0.20% | 538 |
| All Trials | Age 75+ years | 0.179 | 1.00 | No | 1.63 | 0.98 | 2.71 | 0.060 | 26528 | 7213 | 19168 | 27 | 120 | 0.37% | 0.62% | 0.20% | 401 |
| All Trials | Male | 0.293 | 0.99 | No | 1.33 | 0.78 | 2.28 | 0.293 | 23993 | 5489 | 18373 | 23 | 108 | 0.42% | 0.58% | 0.20% | 598 |
| All Trials | Female | † | † | † | 2.27 | 1.29 | 4.02 | 0.005 | 25357 | 6560 | 18677 | 19 | 101 | 0.29% | 0.54% | 0.20% | 402 |
| All Trials | White Race | 0.099 | 1.00 | No | 1.47 | 0.98 | 2.23 | 0.066 | 41369 | 10274 | 30881 | 40 | 174 | 0.39% | 0.56% | 0.20% | 580 |
| All Trials | Non-White Race | † | † | † | 5.87 | 1.32 | 26.13 | 0.020 | 7981 | 1775 | 6169 | * | 35 | * | 0.56% | * | * |
| Trials 0 & 1 | Overall | 0.022 | 1.00 | Yes | 1.61 | 1.07 | 2.43 | 0.022 | 46744 | 10630 | 35870 | 39 | 205 | 0.37% | 0.57% | 0.20% | 493 |
| Trials 0 & 1 | Time stratified: 1-30 days | 0.658 | 0.53 | No | 1.13 | 0.65 | 1.96 | 0.658 | 46744 | 10646 | 36004 | 23 | 71 | 0.22% | 0.20% | 0.00% |  |
| Trials 0 & 1 | Time stratified: 31-91 days | † | † | † | 3.50 | 1.59 | 7.73 | 0.002 | 37975 | 7451 | 30439 | * | 76 | * | 0.25% | * | * |
| Trials 0 & 1 | Time stratified: 92-182 days | † | † | † | 1.11 | 0.46 | 2.67 | 0.809 | 25769 | 3207 | 22497 | * | 58 | * | 0.26% | * | * |
| Trials 0 & 1 | Age 65-74 years | † | † | † | 1.71 | 0.91 | 3.18 | 0.093 | 21731 | 4275 | 17353 | 15 | 88 | 0.35% | 0.50% | 0.20% | 646 |
| Trials 0 & 1 | Age 75+ years | 0.293 | 1.00 | No | 1.57 | 0.92 | 2.68 | 0.098 | 25013 | 6355 | 18517 | 24 | 117 | 0.38% | 0.63% | 0.30% | 397 |
| Trials 0 & 1 | Male | 0.358 | 0.96 | No | 1.30 | 0.74 | 2.30 | 0.358 | 22806 | 4847 | 17832 | 20 | 107 | 0.41% | 0.60% | 0.20% | 539 |
| Trials 0 & 1 | Female | † | † | † | 2.01 | 1.14 | 3.56 | 0.016 | 23938 | 5783 | 18038 | 19 | 98 | 0.33% | 0.54% | 0.20% | 470 |
| Trials 0 & 1 | White Race | 0.185 | 1.00 | No | 1.40 | 0.91 | 2.14 | 0.124 | 39322 | 9104 | 30009 | 37 | 172 | 0.40% | 0.57% | 0.20% | 606 |
| Trials 0 & 1 | Non-White Race | † | † | † | 5.07 | 1.14 | 22.61 | 0.033 | 7422 | 1526 | 5861 | * | 33 | * | 0.56% | * | * |
| Notes: A1 = Atorvastatin initiation; A2 = Newly initiated a different medication other than atorvastatin; N = Total number of patients in the analytic stratum; A = Number of A2 patients without the event; B = Number of A1 patients without the event; C = Number of A2 patients with the event; D = Number of A1 patients with the event; Risk A1 = Cumulative incidence (risk) in the A1 (atorvastatin) group; Risk A2 = Cumulative incidence (risk) in the A2 (comparator) group; RD = Risk difference; NNH = Number needed to harm. ADE Signal Confirmation metrics: q = Within-outcome Benjamini-Hochberg false-discovery-rate adjusted p-value; P(adj>1) = Proportion of 5,000 Monte Carlo draws from the probabilistic quantitative bias analysis in which the residual-confounding-adjusted subdistribution hazard ratio remained greater than 1. Confirmed = 1 if q ≤ 0.05 and P(adj>1) = 1; otherwise 0. Only analyses with at least 20 events in both treatment arms (C ≥ 20 and D ≥ 20) were included in the ADE Signal Confirmation analyses. Analyses that did not meet this threshold were excluded from both the Benjamini-Hochberg correction and the probabilistic quantitative bias analysis (denoted by †). * = Not reportable due to cell counts ≤ 10 per CMS policies. |  |  |  |  |  |  |  |  |  |  |  |  |  |  |  |  |  |

**Table S7 (Continued) Detailed Results for Confirmed ADE Signals**
| Detailed Results for Confirmed ADE Signals |  |  |  |  |  |  |  |  |  |  |  |  |  |  |  |  |  |
| --- | --- | --- | --- | --- | --- | --- | --- | --- | --- | --- | --- | --- | --- | --- | --- | --- | --- |
| Category : Abnormal symptoms |  |  |  |  |  |  |  |  |  |  |  |  |  |  |  |  |  |
| Outcome : Biliary tract disease |  |  |  |  |  |  |  |  |  |  |  |  |  |  |  |  |  |
| Analysis Set | Stratum | ADE Signal Confirmation |  |  | Weighted Fine-Gray |  |  |  | Unadjusted Descriptive Analyses for All Trials |  |  |  |  |  |  |  |  |
|  |  | q | P(adj>1) | Confirmed | sHR | LCL | UCL | p | N | A | B | C | D | Risk A2 | Risk A1 | RD | NNH |
| All Trials | Overall | 0.047 | 0.76 | No | 1.20 | 1.00 | 1.44 | 0.047 | 49510 | 11995 | 36355 | 219 | 941 | 1.79% | 2.52% | 0.70% | 137 |
| All Trials | Time stratified: 1-30 days | 0.120 | 0.91 | No | 1.27 | 0.97 | 1.65 | 0.080 | 49510 | 12105 | 36952 | 109 | 344 | 0.89% | 0.92% | 0.00% | 3341 |
| All Trials | Time stratified: 31-91 days | 0.235 | 0.99 | No | 1.35 | 0.97 | 1.89 | 0.078 | 39885 | 8508 | 30975 | 62 | 340 | 0.72% | 1.09% | 0.40% | 276 |
| All Trials | Time stratified: 92-182 days | 0.325 | 0.00 | No | 0.84 | 0.59 | 1.19 | 0.325 | 26471 | 3535 | 22631 | 48 | 257 | 1.34% | 1.12% | -0.20% | - |
| All Trials | Age 65-74 years | 0.932 | 0.08 | No | 1.04 | 0.77 | 1.41 | 0.777 | 22886 | 4819 | 17605 | 76 | 386 | 1.55% | 2.15% | 0.60% | 169 |
| All Trials | Age 75+ years | 0.040 | 0.98 | No | 1.33 | 1.06 | 1.66 | 0.013 | 26624 | 7176 | 18750 | 143 | 555 | 1.95% | 2.87% | 0.90% | 109 |
| All Trials | Male | 0.954 | 0.00 | No | 0.99 | 0.76 | 1.29 | 0.954 | 24067 | 5462 | 18054 | 108 | 443 | 1.94% | 2.39% | 0.50% | 219 |
| All Trials | Female | 0.016 | 1.00 | Yes | 1.45 | 1.14 | 1.85 | 0.003 | 25443 | 6533 | 18301 | 111 | 498 | 1.67% | 2.65% | 1.00% | 102 |
| All Trials | White Race | 0.094 | 0.82 | No | 1.22 | 1.00 | 1.48 | 0.047 | 41500 | 10232 | 30314 | 183 | 771 | 1.76% | 2.48% | 0.70% | 138 |
| All Trials | Non-White Race | 0.952 | 0.44 | No | 1.11 | 0.72 | 1.73 | 0.635 | 8010 | 1763 | 6041 | 36 | 170 | 2.00% | 2.74% | 0.70% | 136 |
| Trials 0 & 1 | Overall | 0.085 | 0.71 | No | 1.18 | 0.98 | 1.43 | 0.085 | 46744 | 10479 | 35167 | 190 | 908 | 1.78% | 2.52% | 0.70% | 136 |
| Trials 0 & 1 | Time stratified: 1-30 days | 0.265 | 0.78 | No | 1.21 | 0.92 | 1.59 | 0.177 | 46744 | 10571 | 35743 | 98 | 332 | 0.92% | 0.92% | 0.00% | 56953 |
| Trials 0 & 1 | Time stratified: 31-91 days | 0.268 | 1.00 | No | 1.37 | 0.95 | 1.99 | 0.089 | 37683 | 7358 | 29948 | 51 | 326 | 0.69% | 1.08% | 0.40% | 257 |
| Trials 0 & 1 | Time stratified: 92-182 days | 0.376 | 0.00 | No | 0.84 | 0.58 | 1.23 | 0.376 | 25349 | 3127 | 21931 | 41 | 250 | 1.29% | 1.13% | -0.20% |  |
| Trials 0 & 1 | Age 65-74 years | 0.940 | 0.00 | No | 1.01 | 0.74 | 1.39 | 0.940 | 21731 | 4223 | 17068 | 67 | 373 | 1.56% | 2.14% | 0.60% | 173 |
| Trials 0 & 1 | Age 75+ years | 0.061 | 0.98 | No | 1.32 | 1.04 | 1.68 | 0.020 | 25013 | 6256 | 18099 | 123 | 535 | 1.93% | 2.87% | 0.90% | 106 |
| Trials 0 & 1 | Male | 0.791 | 0.00 | No | 0.94 | 0.72 | 1.23 | 0.659 | 22806 | 4771 | 17507 | 96 | 432 | 1.97% | 2.41% | 0.40% | 230 |
| Trials 0 & 1 | Female | 0.019 | 1.00 | Yes | 1.49 | 1.14 | 1.94 | 0.003 | 23938 | 5708 | 17660 | 94 | 476 | 1.62% | 2.62% | 1.00% | 100 |
| Trials 0 & 1 | White Race | 0.204 | 0.74 | No | 1.19 | 0.97 | 1.46 | 0.102 | 39322 | 8979 | 29430 | 162 | 751 | 1.77% | 2.49% | 0.70% | 140 |
| Trials 0 & 1 | Non-White Race | 0.858 | 0.60 | No | 1.15 | 0.71 | 1.87 | 0.572 | 7422 | 1500 | 5737 | 28 | 157 | 1.83% | 2.66% | 0.80% | 120 |

Notes: A1 = Atorvastatin initiation; A2 = Newly initiated a different medication other than atorvastatin; N = Total number of patients in the analytic stratum; A = Number of A2 patients without the event; B = Number of A1 patients without the event; C = Number of A2 patients with the event; D = Number of A1 patients with the event; Risk A1 = Cumulative incidence (risk) in the A1 (atorvastatin) group; Risk A2 = Cumulative incidence (risk) in the A2 (comparator) group; RD = Risk difference; NNH = Number needed to harm. ADE Signal Confirmation metrics: q = Within-outcome Benjamini-Hochberg false-discovery-rate adjusted p-value; P(adj>1) = Proportion of 5,000 Monte Carlo draws from the probabilistic quantitative bias analysis in which the residual-confounding-adjusted subdistribution hazard ratio remained greater than 1. Confirmed = 1 if q ≤ 0.05 and P(adj>1) = 1; otherwise 0. Only analyses with at least 20 events in both treatment arms (C ≥ 20 and D ≥ 20) were included in the ADE Signal Confirmation analyses. Analyses that did not meet this threshold were excluded from both the Benjamini-Hochberg correction and the probabilistic quantitative bias analysis (denoted by †). \* = Not reportable due to cell counts ≤ 10 per CMS policies.

**Table S8.** Quantitative bias analysis assessing robustness of selected associations to residual confounding

|  |  |  |  |  |  |  |  |  |  |  |  |  |  |  |  |
| --- | --- | --- | --- | --- | --- | --- | --- | --- | --- | --- | --- | --- | --- | --- | --- |
| Confirmed Outcomes |  |  | Weighted Fine-Gray |  |  |  | E-Value |  | Probabilistic bias-adjusted |  |  |  | Deterministic bias-adjusted |  |  |
| Outcome | Analytic Set | Stratum | sHR | LCL | UCL | p | Estimate | CI | sHR-adj | 2.5<sup>th</sup>-adj | 97.5<sup>th</sup>-adj | P(adj>1) | sHR-adj | min | max |
| Bleeding events |  |  |  |  |  |  |  |  |  |  |  |  |  |  |  |
| Acute hemorrhagic cerebrovascular disease | All trials | Overall | 1.43 | 1.00 | 2.04 | 0.050 | 2.21 | 1.01 | 1.27 | 1.08 | 1.39 | 1.00 | 1.31 | 1.03 | 1.41 |
| Acute hemorrhagic cerebrovascular disease | All trials | Time stratified: 1-30 days | 2.20 | 1.35 | 3.58 | 0.002 | 3.82 | 2.03 | 1.95 | 1.66 | 2.13 | 1.00 | 2.01 | 1.59 | 2.17 |
| Acute hemorrhagic cerebrovascular disease | Trials 0 & 1 | Overall | 1.50 | 1.02 | 2.20 | 0.039 | 2.36 | 1.16 | 1.33 | 1.14 | 1.46 | 1.00 | 1.37 | 1.08 | 1.48 |
| Intracranial Hemorrhage (traumatic & non-traumatic) | All trials | Time stratified: 1-30 days | 1.91 | 1.13 | 3.23 | 0.016 | 3.22 | 1.50 | 1.70 | 1.44 | 1.85 | 1.00 | 1.74 | 1.38 | 1.88 |
| Cardiac valve events |  |  |  |  |  |  |  |  |  |  |  |  |  |  |  |
| Nonrheumatic and unspecified valve disorders | All trials | Time stratified: 92-182 days | 1.48 | 1.13 | 1.94 | 0.004 | 2.33 | 1.52 | 1.32 | 1.12 | 1.44 | 1.00 | 1.36 | 1.07 | 1.46 |
| Nonrheumatic and unspecified valve disorders | Trials 0 & 1 | Time stratified: 92-182 days | 1.58 | 1.18 | 2.11 | 0.002 | 2.53 | 1.65 | 1.41 | 1.19 | 1.54 | 1.00 | 1.44 | 1.14 | 1.56 |
| Nonrheumatic valve disease | All trials | Time stratified: 92-182 days | 1.50 | 1.14 | 1.98 | 0.004 | 2.38 | 1.54 | 1.34 | 1.14 | 1.46 | 1.00 | 1.38 | 1.09 | 1.49 |
| Nonrheumatic valve disease | Trials 0 & 1 | Time stratified: 92-182 days | 1.60 | 1.19 | 2.15 | 0.002 | 2.58 | 1.67 | 1.42 | 1.21 | 1.56 | 1.00 | 1.46 | 1.16 | 1.58 |
| Cardiac valve intervention procedure | All trials | Overall | 1.83 | 1.12 | 2.98 | 0.016 | 3.06 | 1.49 | 1.62 | 1.38 | 1.78 | 1.00 | 1.67 | 1.32 | 1.80 |
| Cardiac valve intervention procedure | Trials 0 & 1 | Overall | 1.74 | 1.04 | 2.89 | 0.034 | 2.87 | 1.25 | 1.54 | 1.31 | 1.69 | 1.00 | 1.59 | 1.25 | 1.72 |
| Composite: valve disorder or intervention | All trials | Time stratified: 92-182 days | 1.44 | 1.11 | 1.87 | 0.007 | 2.23 | 1.45 | 1.28 | 1.09 | 1.40 | 1.00 | 1.32 | 1.04 | 1.42 |
| Composite: valve disorder or intervention | Trials 0 & 1 | Time stratified: 92-182 days | 1.52 | 1.15 | 2.01 | 0.003 | 2.41 | 1.56 | 1.36 | 1.15 | 1.48 | 1.00 | 1.39 | 1.10 | 1.50 |
| Sprains, strains, and tendon injury |  |  |  |  |  |  |  |  |  |  |  |  |  |  |  |
| Sprains and strains, initial encounter | All trials | Male | 1.72 | 1.20 | 2.45 | 0.003 | 2.82 | 1.70 | 1.53 | 1.30 | 1.67 | 1.00 | 1.57 | 1.24 | 1.69 |
| Sprains and strains, initial encounter | Trials 0 & 1 | Male | 1.73 | 1.18 | 2.54 | 0.005 | 2.85 | 1.64 | 1.54 | 1.31 | 1.68 | 1.00 | 1.58 | 1.25 | 1.71 |
| Sprain | All trials | Male | 1.94 | 1.21 | 3.09 | 0.006 | 3.28 | 1.72 | 1.72 | 1.46 | 1.88 | 1.00 | 1.77 | 1.40 | 1.91 |
| Composite: sprain/strain/tendon injury | All trials | Male | 1.66 | 1.24 | 2.21 | 0.001 | 2.70 | 1.79 | 1.47 | 1.25 | 1.61 | 1.00 | 1.52 | 1.20 | 1.64 |
| Composite: sprain/strain/tendon injury | Trials 0 & 1 | Male | 1.76 | 1.28 | 2.41 | 0.000 | 2.91 | 1.89 | 1.56 | 1.33 | 1.71 | 1.00 | 1.61 | 1.27 | 1.74 |
| Dizziness |  |  |  |  |  |  |  |  |  |  |  |  |  |  |  |
| General sensation/perception signs and symptoms | All trials | Non-White Race | 1.40 | 1.05 | 1.86 | 0.020 | 2.15 | 1.30 | 1.25 | 1.06 | 1.36 | 1.00 | 1.28 | 1.01 | 1.38 |
| Dizziness and giddiness | All trials | Non-White Race | 1.43 | 1.06 | 1.91 | 0.017 | 2.21 | 1.33 | 1.27 | 1.08 | 1.39 | 1.00 | 1.30 | 1.03 | 1.41 |
| Abnormal symptoms |  |  |  |  |  |  |  |  |  |  |  |  |  |  |  |
| Acute hepatic failure – narrow definition | All trials | Overall | 1.72 | 1.16 | 2.55 | 0.007 | 2.83 | 1.59 | 1.53 | 1.30 | 1.67 | 1.00 | 1.57 | 1.24 | 1.70 |
| Acute hepatic failure – narrow definition | Trials 0 & 1 | Overall | 1.61 | 1.07 | 2.43 | 0.022 | 2.61 | 1.35 | 1.44 | 1.22 | 1.57 | 1.00 | 1.48 | 1.16 | 1.59 |
| Biliary tract disease | All trials | Female | 1.45 | 1.14 | 1.85 | 0.003 | 2.26 | 1.53 | 1.29 | 1.10 | 1.41 | 1.00 | 1.33 | 1.05 | 1.43 |
| Biliary tract disease | Trials 0 & 1 | Female | 1.49 | 1.14 | 1.94 | 0.003 | 2.35 | 1.55 | 1.32 | 1.12 | 1.45 | 1.00 | 1.36 | 1.08 | 1.47 |

Notes: Weighted Fine-Gray columns show the observed subdistribution hazard ratio (sHR) with lower and upper 95% confidence limits (LCL, UCL) and p-value from the inverse-probability-weighted Fine-Gray model. E-Value Estimate is the minimum risk-ratio association an unmeasured confounder would need with both exposure and outcome to fully explain away the observed point estimate; E-Value CI is the corresponding value required to explain away the confidence interval. Probabilistic bias-adjusted columns report the median adjusted sHR (sHR-adj), the 2.5th and 97.5th percentiles of the distribution of adjusted sHRs obtained by sampling bias parameters from their prior distributions, and P(adj>1), the proportion of simulated bias-adjusted sHRs that remained greater than 1. Deterministic bias-adjusted columns report the median adjusted sHR (sHR-adj) and the minimum-maximum adjusted values across fixed bias-parameter scenarios. Full bias parameters and methods are provided in the Methods/Supplement. Analytic Set “All trials” is the primary analysis; “Trials 0 & 1” is a sensitivity analysis.

**Table S9.**
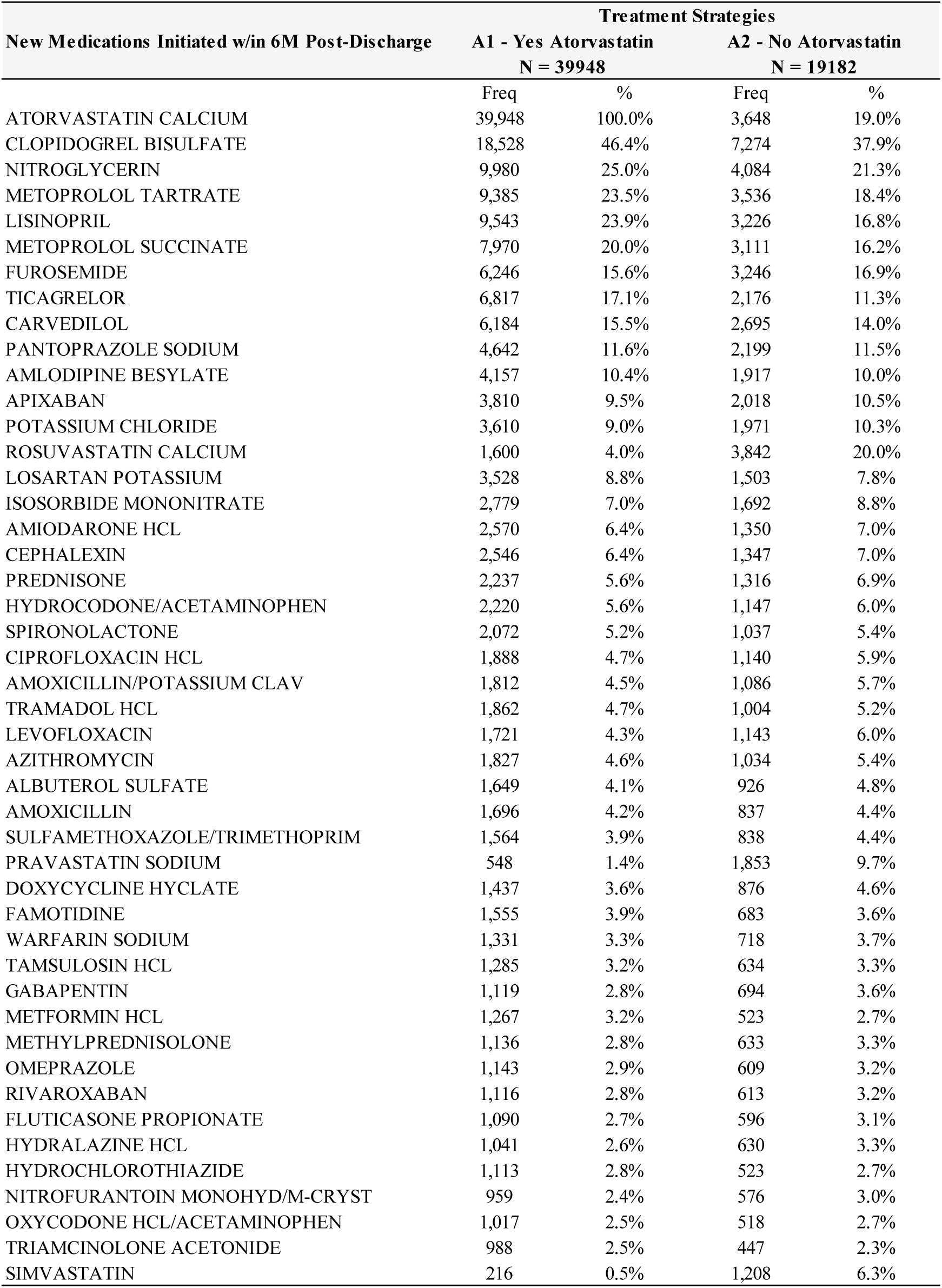
New Medications Initiated w/in 6M Post-Discharge

**Table S10.**
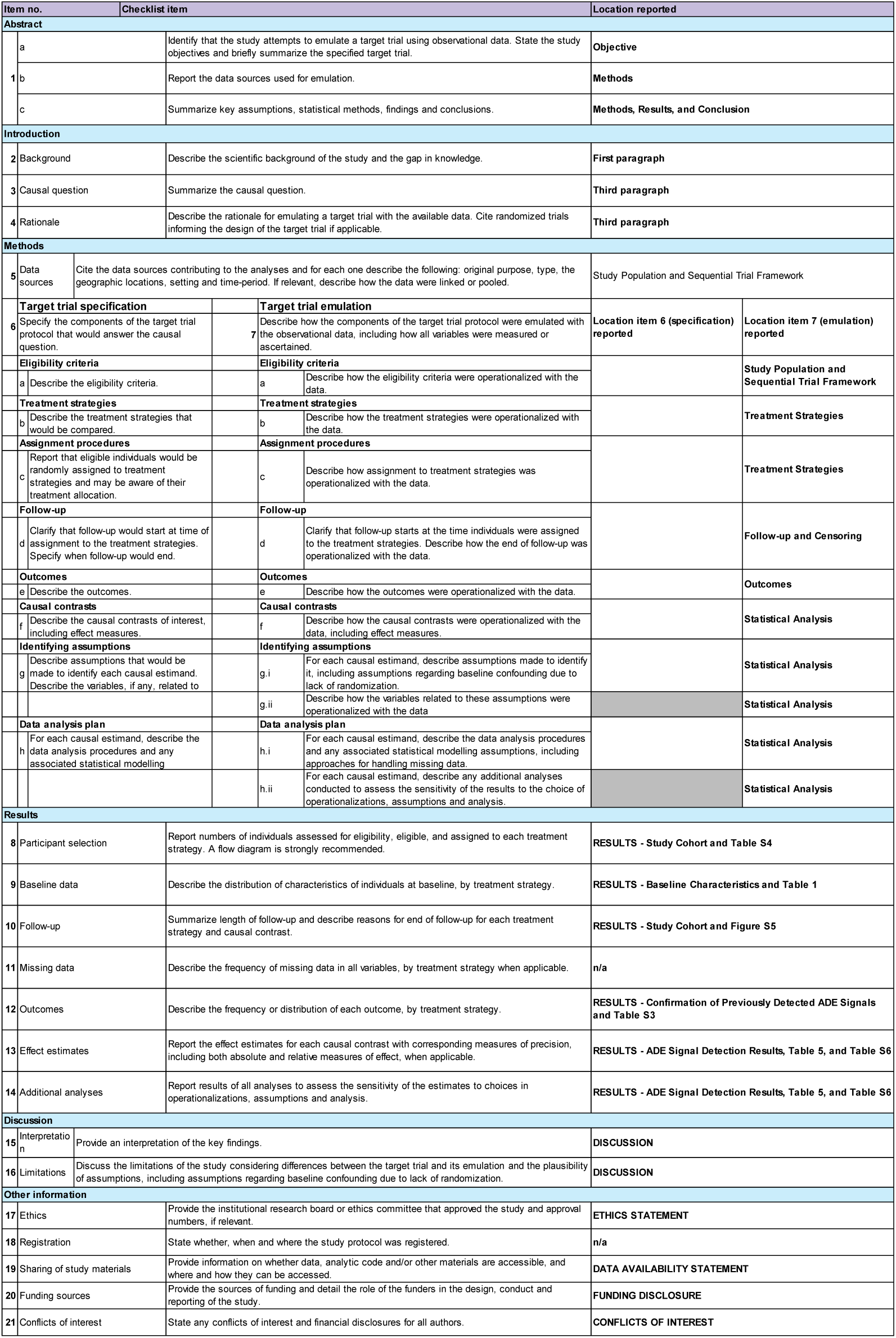
TARGET Checklist

**Table S11.**
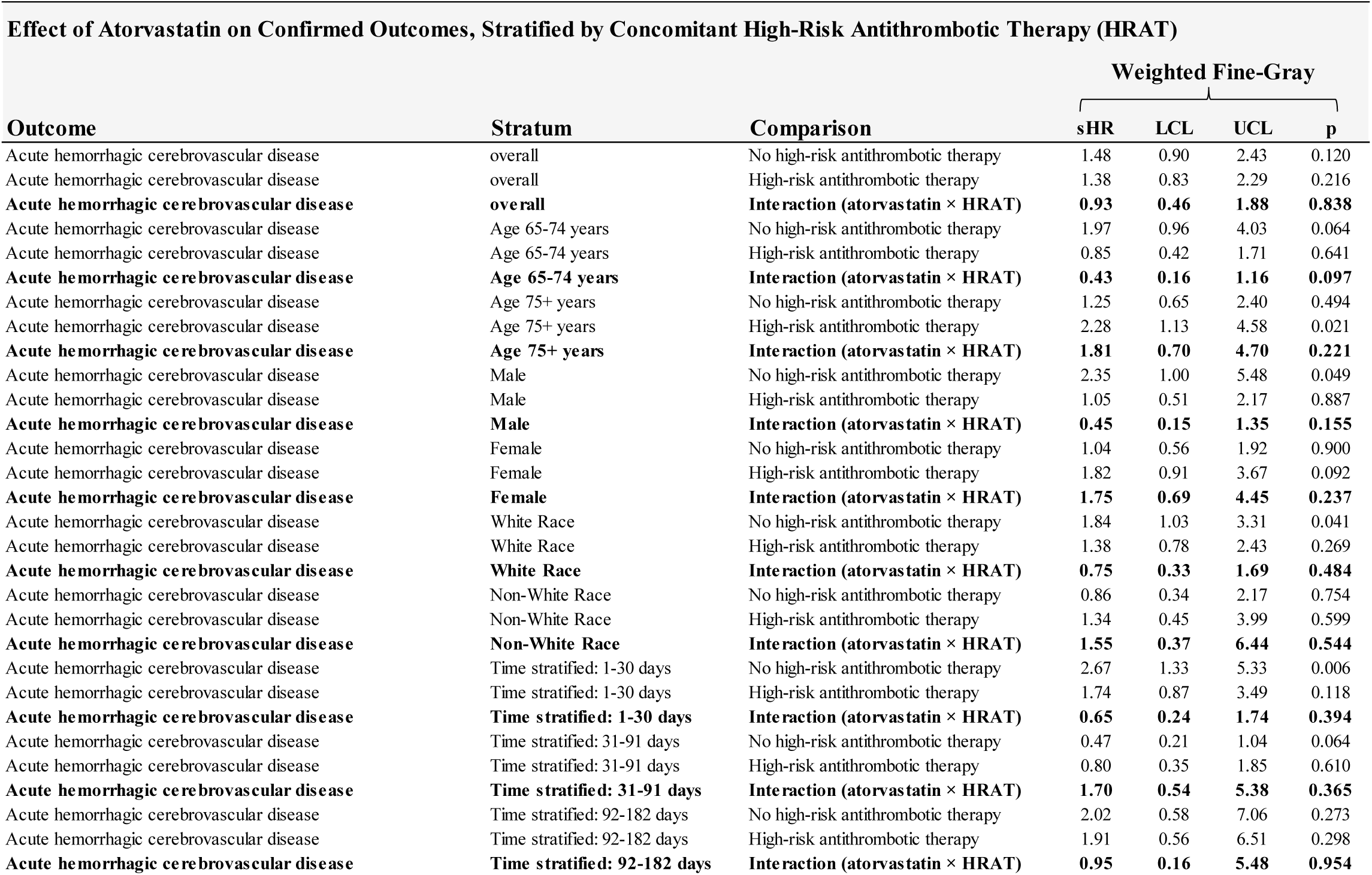
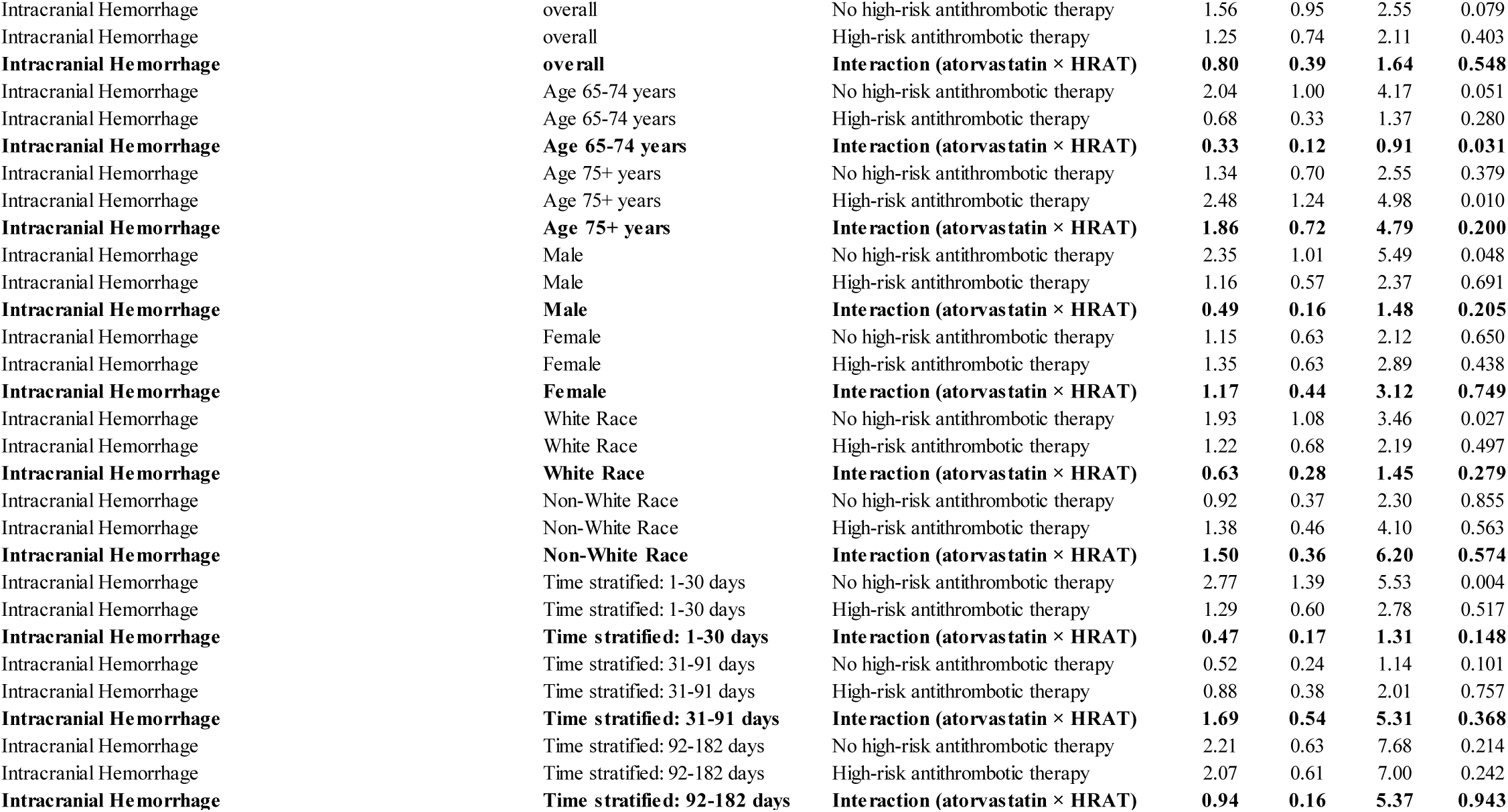
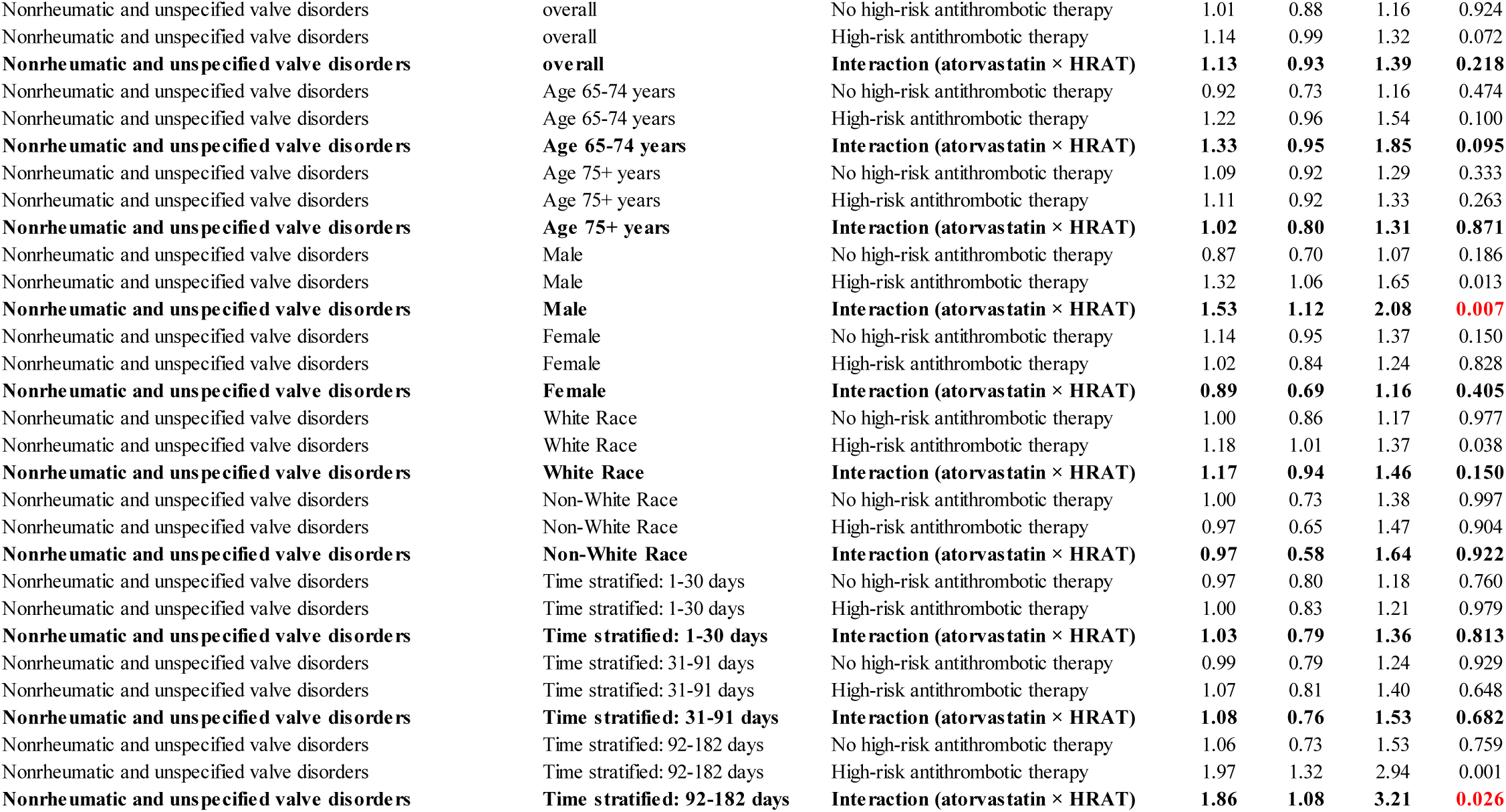
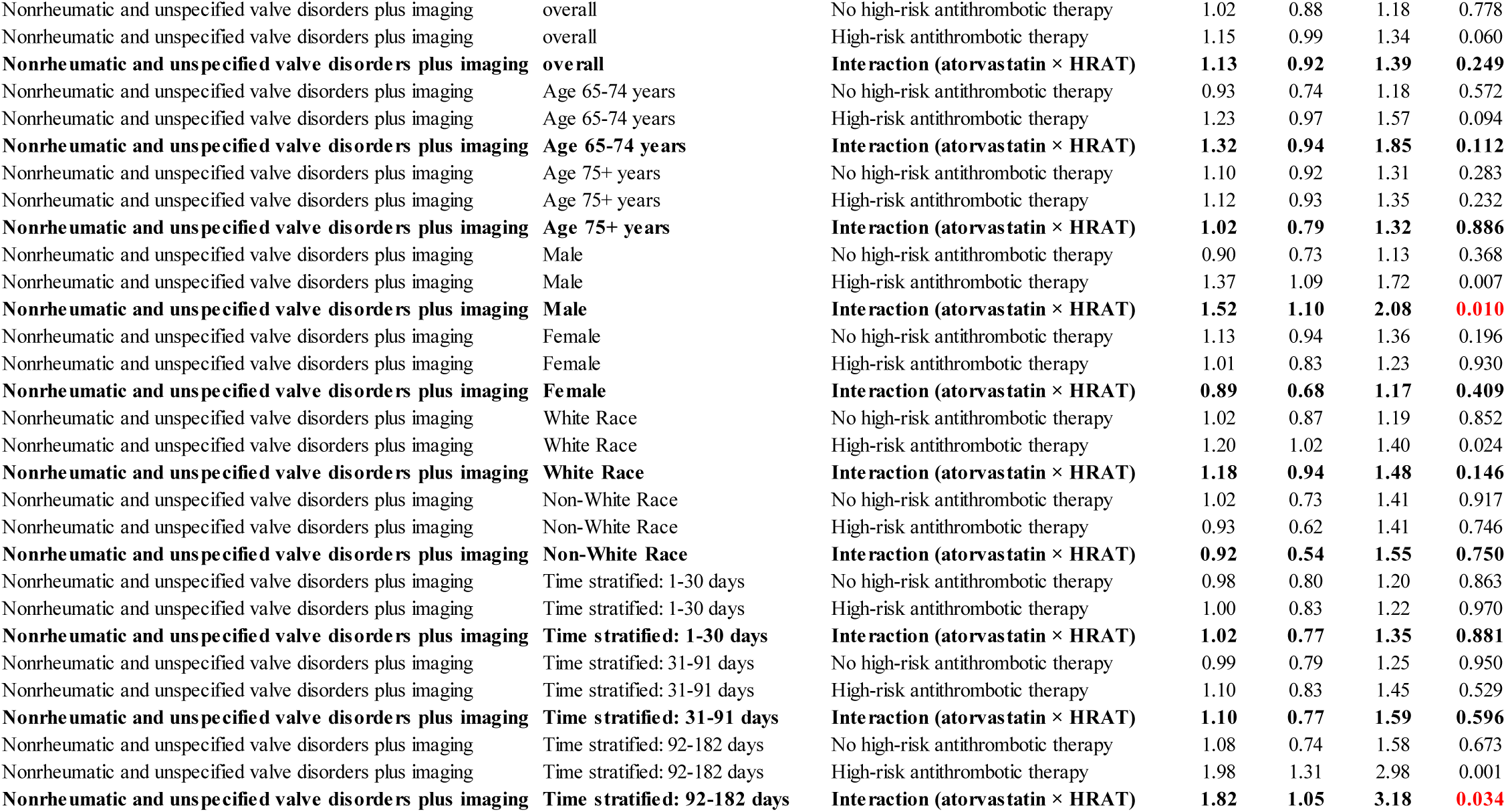
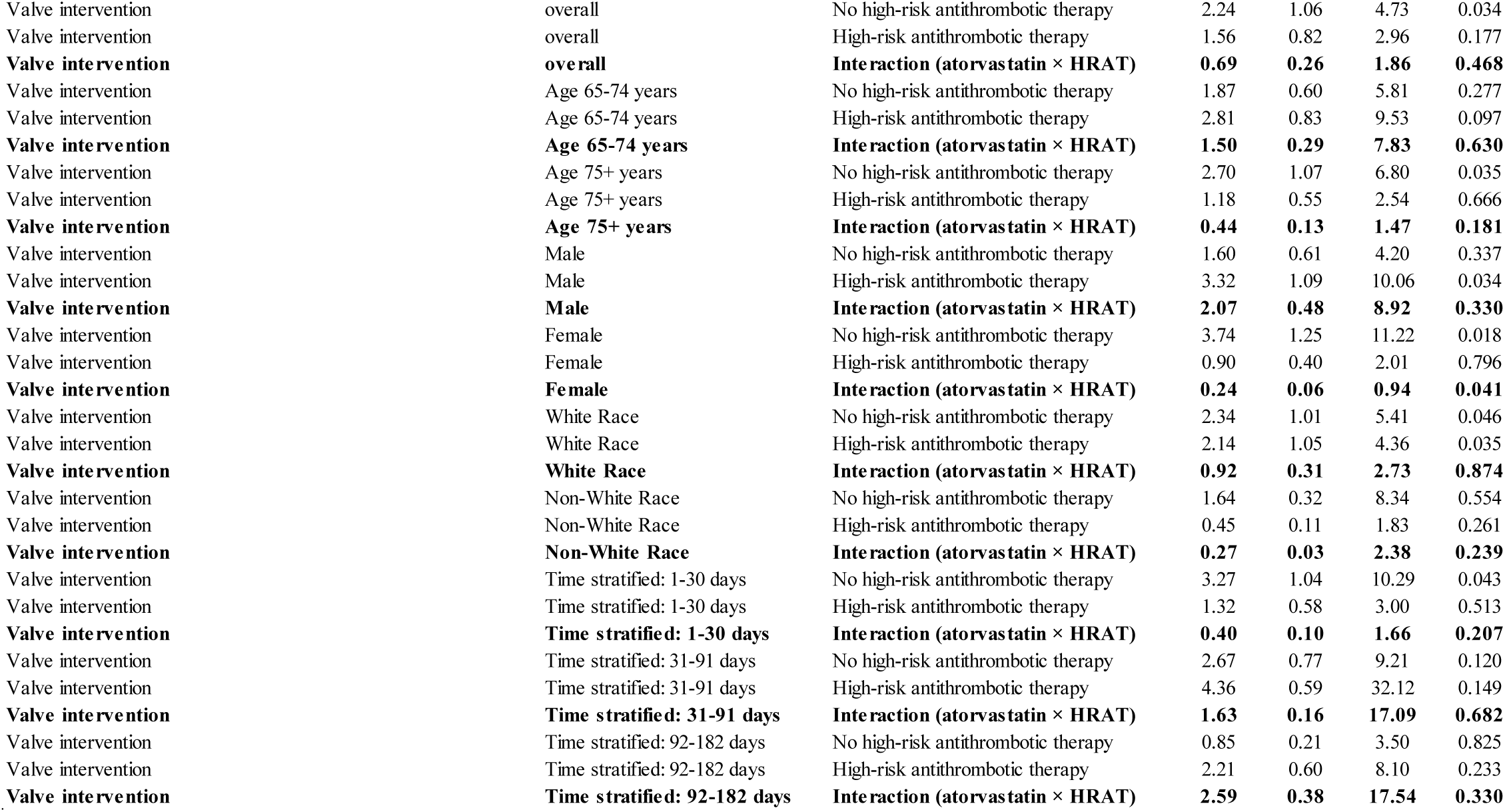
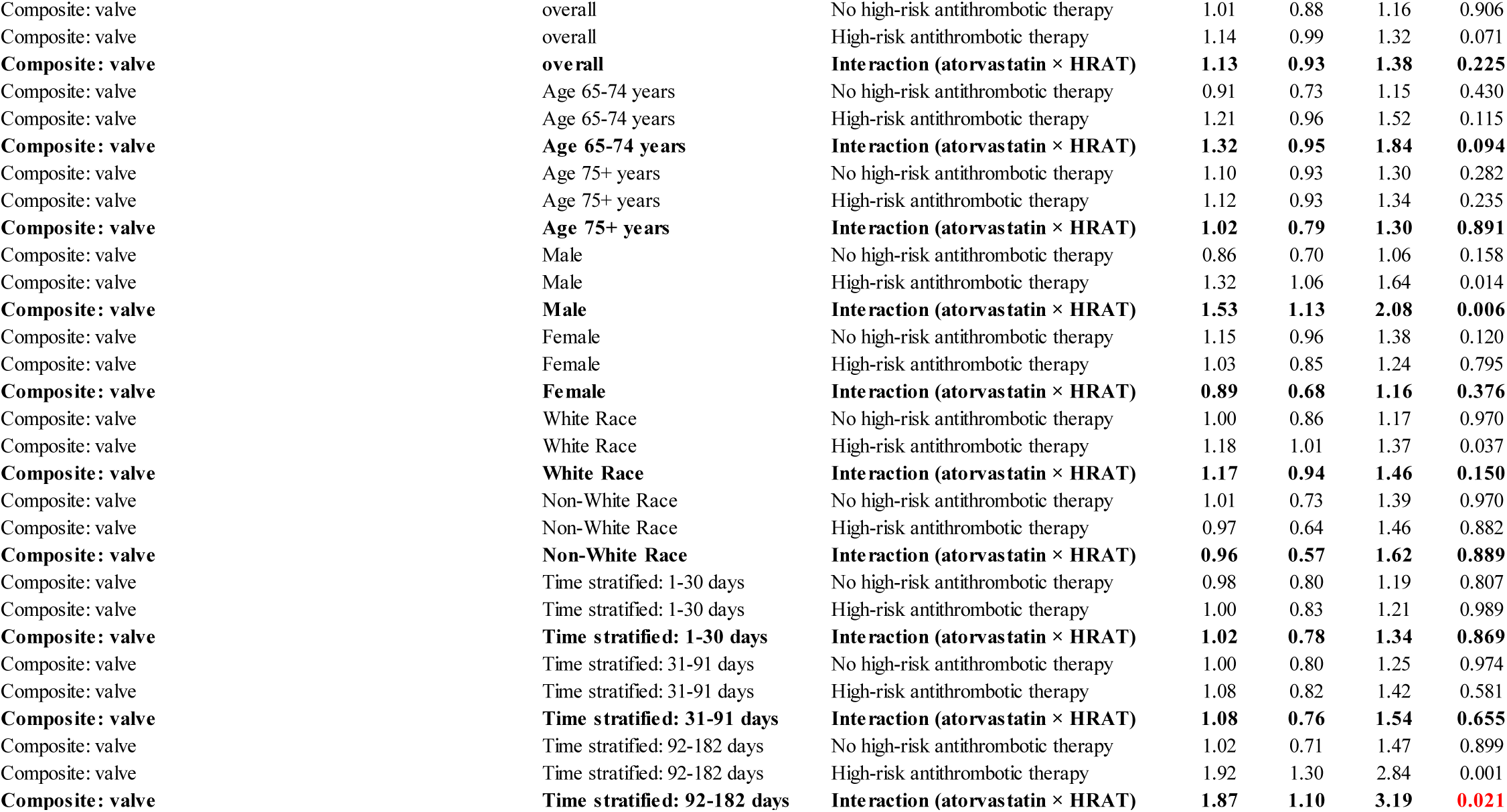
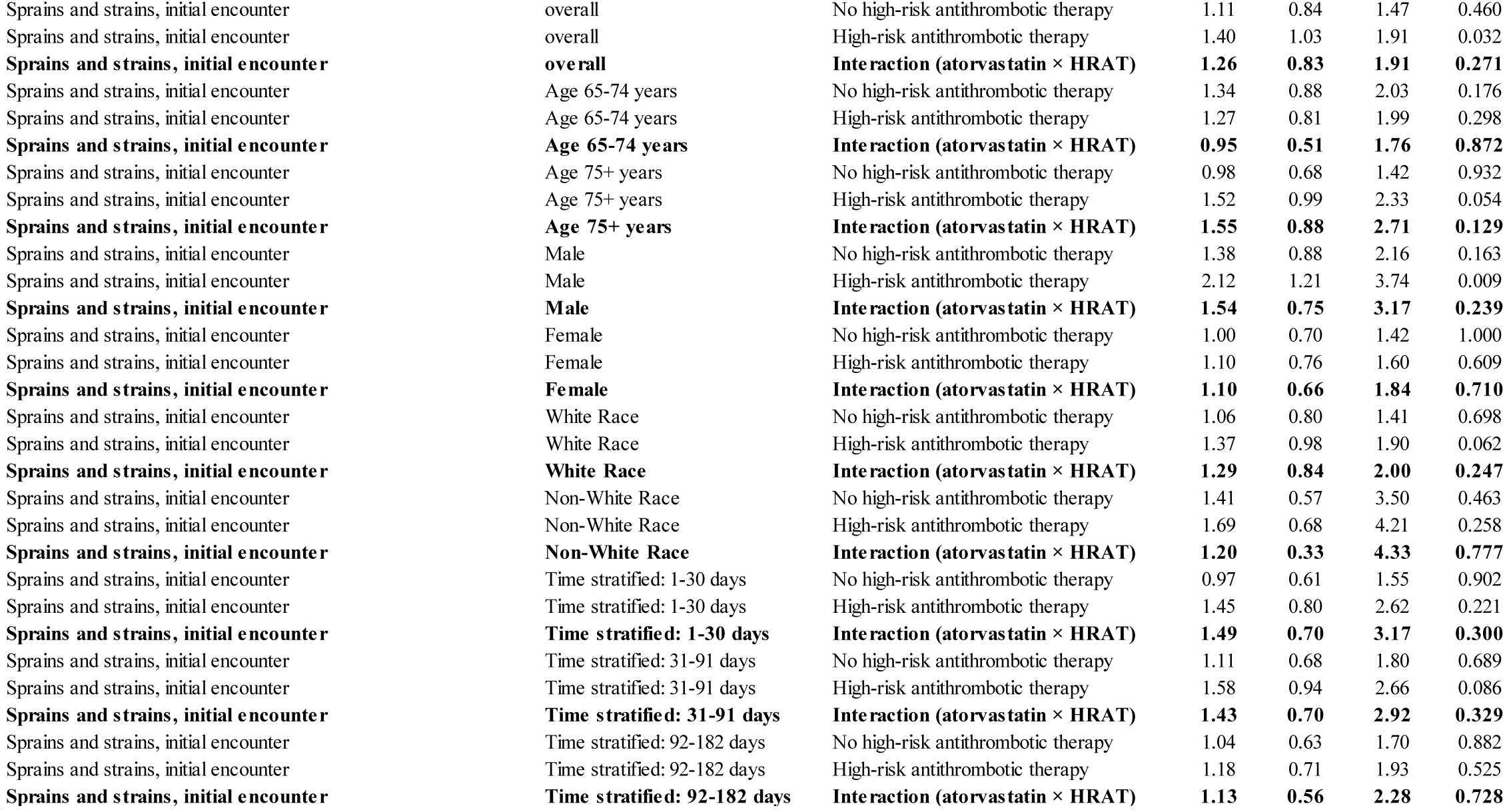
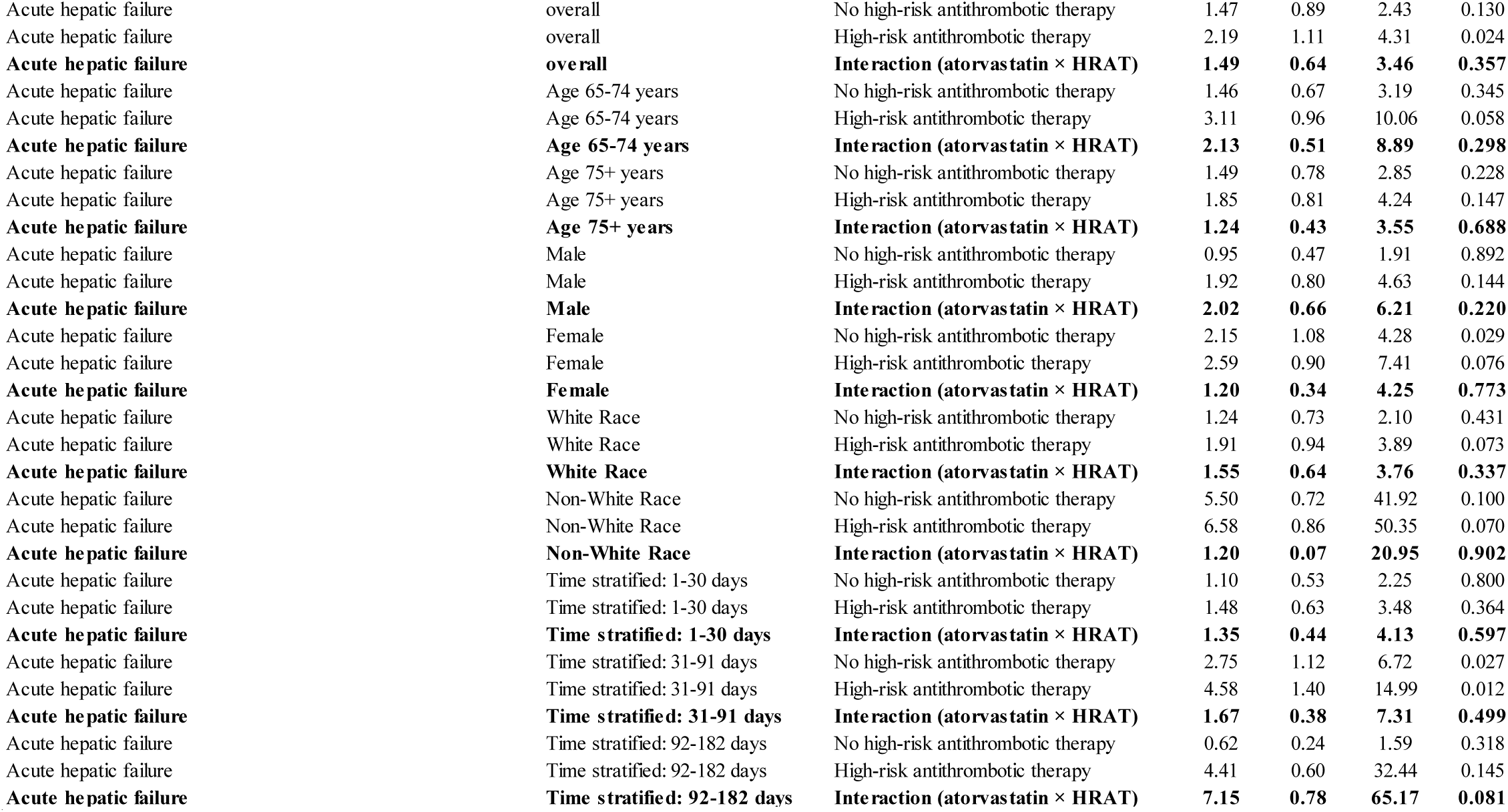
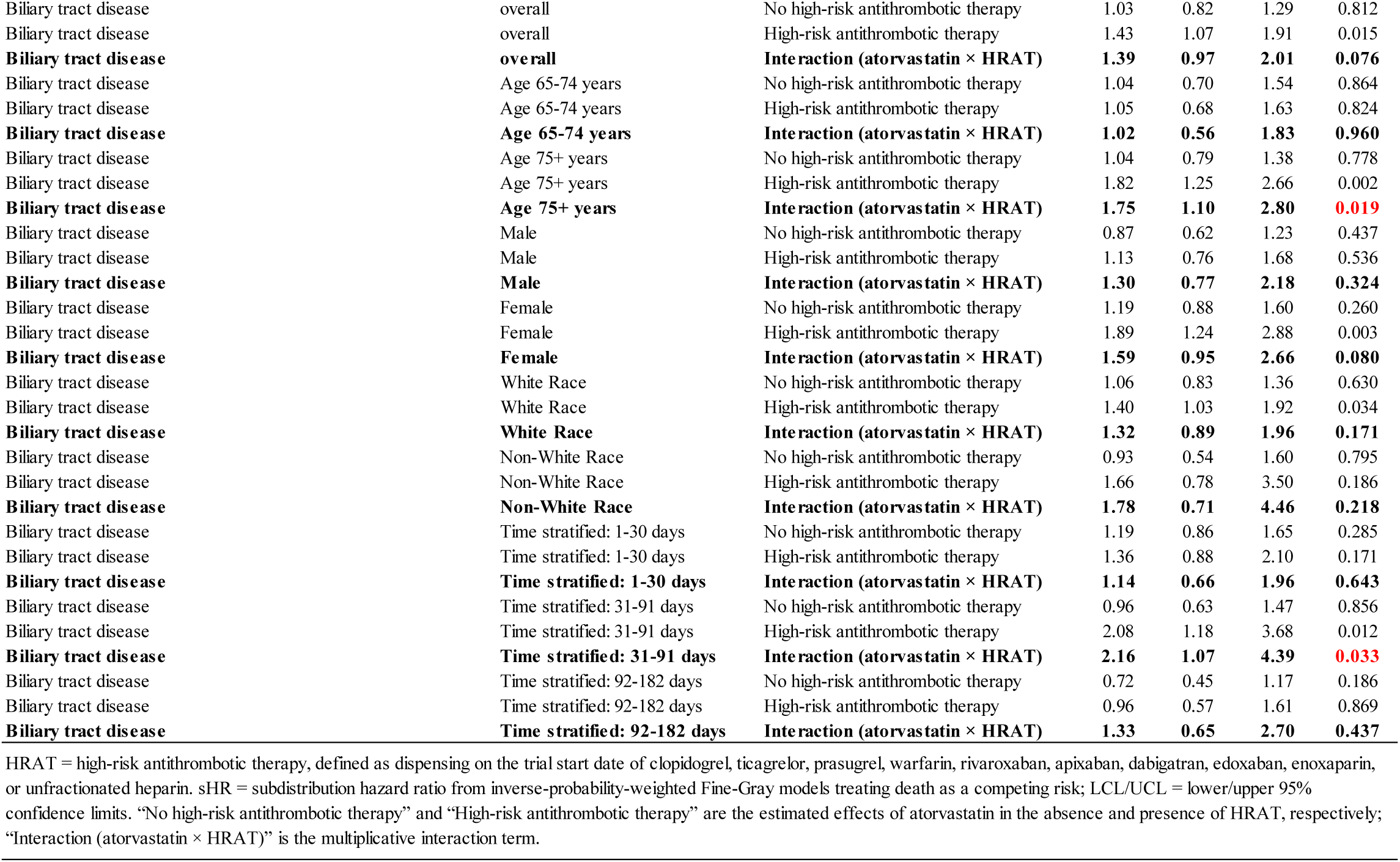
Effect of Atorvastatin on Confirmed Outcomes, Stratified by Concomitant High-Risk Antithrombotic Therapy (HRAT)

|  |  |  |  |  |  |  |
| --- | --- | --- | --- | --- | --- | --- |
| Outcome | Stratum | Comparison | Weighted Fine-Gray |  |  |  |
|  |  |  | sHR | LCL | UCL | p |
| Acute hemorrhagic cerebrovascular disease | overall | No high-risk antithrombotic therapy | 1.48 | 0.90 | 2.43 | 0.120 |
| Acute hemorrhagic cerebrovascular disease | overall | High-risk antithrombotic therapy | 1.38 | 0.83 | 2.29 | 0.216 |
| **Acute hemorrhagic cerebrovascular disease** | **overall** | **Interaction (atorvastatin × HRAT)** | **0.93** | **0.46** | **1.88** | **0.838** |
| Acute hemorrhagic cerebrovascular disease | Age 65-74 years | No high-risk antithrombotic therapy | 1.97 | 0.96 | 4.03 | 0.064 |
| Acute hemorrhagic cerebrovascular disease | Age 65-74 years | High-risk antithrombotic therapy | 0.85 | 0.42 | 1.71 | 0.641 |
| **Acute hemorrhagic cerebrovascular disease** | **Age 65-74 years** | **Interaction (atorvastatin × HRAT)** | **0.43** | **0.16** | **1.16** | **0.097** |
| Acute hemorrhagic cerebrovascular disease | Age 75+ years | No high-risk antithrombotic therapy | 1.25 | 0.65 | 2.40 | 0.494 |
| Acute hemorrhagic cerebrovascular disease | Age 75+ years | High-risk antithrombotic therapy | 2.28 | 1.13 | 4.58 | 0.021 |
| **Acute hemorrhagic cerebrovascular disease** | **Age 75+ years** | **Interaction (atorvastatin × HRAT)** | **1.81** | **0.70** | **4.70** | **0.221** |
| Acute hemorrhagic cerebrovascular disease | Male | No high-risk antithrombotic therapy | 2.35 | 1.00 | 5.48 | 0.049 |
| Acute hemorrhagic cerebrovascular disease | Male | High-risk antithrombotic therapy | 1.05 | 0.51 | 2.17 | 0.887 |
| **Acute hemorrhagic cerebrovascular disease** | **Male** | **Interaction (atorvastatin × HRAT)** | **0.45** | **0.15** | **1.35** | **0.155** |
| Acute hemorrhagic cerebrovascular disease | Female | No high-risk antithrombotic therapy | 1.04 | 0.56 | 1.92 | 0.900 |
| Acute hemorrhagic cerebrovascular disease | Female | High-risk antithrombotic therapy | 1.82 | 0.91 | 3.67 | 0.092 |
| **Acute hemorrhagic cerebrovascular disease** | **Female** | **Interaction (atorvastatin × HRAT)** | **1.75** | **0.69** | **4.45** | **0.237** |
| Acute hemorrhagic cerebrovascular disease | White Race | No high-risk antithrombotic therapy | 1.84 | 1.03 | 3.31 | 0.041 |
| Acute hemorrhagic cerebrovascular disease | White Race | High-risk antithrombotic therapy | 1.38 | 0.78 | 2.43 | 0.269 |
| **Acute hemorrhagic cerebrovascular disease** | **White Race** | **Interaction (atorvastatin × HRAT)** | **0.75** | **0.33** | **1.69** | **0.484** |
| Acute hemorrhagic cerebrovascular disease | Non-White Race | No high-risk antithrombotic therapy | 0.86 | 0.34 | 2.17 | 0.754 |
| Acute hemorrhagic cerebrovascular disease | Non-White Race | High-risk antithrombotic therapy | 1.34 | 0.45 | 3.99 | 0.599 |
| **Acute hemorrhagic cerebrovascular disease** | **Non-White Race** | **Interaction (atorvastatin × HRAT)** | **1.55** | **0.37** | **6.44** | **0.544** |
| Acute hemorrhagic cerebrovascular disease | Time stratified: 1-30 days | No high-risk antithrombotic therapy | 2.67 | 1.33 | 5.33 | 0.006 |
| Acute hemorrhagic cerebrovascular disease | Time stratified: 1-30 days | High-risk antithrombotic therapy | 1.74 | 0.87 | 3.49 | 0.118 |
| **Acute hemorrhagic cerebrovascular disease** | **Time stratified: 1-30 days** | **Interaction (atorvastatin × HRAT)** | **0.65** | **0.24** | **1.74** | **0.394** |
| Acute hemorrhagic cerebrovascular disease | Time stratified: 31-91 days | No high-risk antithrombotic therapy | 0.47 | 0.21 | 1.04 | 0.064 |
| Acute hemorrhagic cerebrovascular disease | Time stratified: 31-91 days | High-risk antithrombotic therapy | 0.80 | 0.35 | 1.85 | 0.610 |
| **Acute hemorrhagic cerebrovascular disease** | **Time stratified: 31-91 days** | **Interaction (atorvastatin × HRAT)** | **1.70** | **0.54** | **5.38** | **0.365** |
| Acute hemorrhagic cerebrovascular disease | Time stratified: 92-182 days | No high-risk antithrombotic therapy | 2.02 | 0.58 | 7.06 | 0.273 |
| Acute hemorrhagic cerebrovascular disease | Time stratified: 92-182 days | High-risk antithrombotic therapy | 1.91 | 0.56 | 6.51 | 0.298 |
| **Acute hemorrhagic cerebrovascular disease** | **Time stratified: 92-182 days** | **Interaction (atorvastatin × HRAT)** | **0.95** | **0.16** | **5.48** | **0.954** |

**Table S11 (continued) Effect of Atorvastatin on Confirmed Outcomes, Stratified by Concomitant High-Risk Antithrombotic Therapy (HRAT)**
|  |  |  |  |  |  |  |
| --- | --- | --- | --- | --- | --- | --- |
| Outcome | Stratum | Comparison | Weighted Fine-Gray |  |  |  |
|  |  |  | sHR | LCL | UCL | p |
| Intracranial Hemorrhage | overall | No high-risk antithrombotic therapy | 1.56 | 0.95 | 2.55 | 0.079 |
| Intracranial Hemorrhage | overall | High-risk antithrombotic therapy | 1.25 | 0.74 | 2.11 | 0.403 |
| **Intracranial Hemorrhage** | **overall** | **Interaction (atorvastatin × HRAT)** | **0.80** | **0.39** | **1.64** | **0.548** |
| Intracranial Hemorrhage | Age 65-74 years | No high-risk antithrombotic therapy | 2.04 | 1.00 | 4.17 | 0.051 |
| Intracranial Hemorrhage | Age 65-74 years | High-risk antithrombotic therapy | 0.68 | 0.33 | 1.37 | 0.280 |
| **Intracranial Hemorrhage** | **Age 65-74 years** | **Interaction (atorvastatin × HRAT)** | **0.33** | **0.12** | **0.91** | **0.031** |
| Intracranial Hemorrhage | Age 75+ years | No high-risk antithrombotic therapy | 1.34 | 0.70 | 2.55 | 0.379 |
| Intracranial Hemorrhage | Age 75+ years | High-risk antithrombotic therapy | 2.48 | 1.24 | 4.98 | 0.010 |
| **Intracranial Hemorrhage** | **Age 75+ years** | **Interaction (atorvastatin × HRAT)** | **1.86** | **0.72** | **4.79** | **0.200** |
| Intracranial Hemorrhage | Male | No high-risk antithrombotic therapy | 2.35 | 1.01 | 5.49 | 0.048 |
| Intracranial Hemorrhage | Male | High-risk antithrombotic therapy | 1.16 | 0.57 | 2.37 | 0.691 |
| **Intracranial Hemorrhage** | **Male** | **Interaction (atorvastatin × HRAT)** | **0.49** | **0.16** | **1.48** | **0.205** |
| Intracranial Hemorrhage | Female | No high-risk antithrombotic therapy | 1.15 | 0.63 | 2.12 | 0.650 |
| Intracranial Hemorrhage | Female | High-risk antithrombotic therapy | 1.35 | 0.63 | 2.89 | 0.438 |
| **Intracranial Hemorrhage** | **Female** | **Interaction (atorvastatin × HRAT)** | **1.17** | **0.44** | **3.12** | **0.749** |
| Intracranial Hemorrhage | White Race | No high-risk antithrombotic therapy | 1.93 | 1.08 | 3.46 | 0.027 |
| Intracranial Hemorrhage | White Race | High-risk antithrombotic therapy | 1.22 | 0.68 | 2.19 | 0.497 |
| **Intracranial Hemorrhage** | **White Race** | **Interaction (atorvastatin × HRAT)** | **0.63** | **0.28** | **1.45** | **0.279** |
| Intracranial Hemorrhage | Non-White Race | No high-risk antithrombotic therapy | 0.92 | 0.37 | 2.30 | 0.855 |
| Intracranial Hemorrhage | Non-White Race | High-risk antithrombotic therapy | 1.38 | 0.46 | 4.10 | 0.563 |
| **Intracranial Hemorrhage** | **Non-White Race** | **Interaction (atorvastatin × HRAT)** | **1.50** | **0.36** | **6.20** | **0.574** |
| Intracranial Hemorrhage | Time stratified: 1-30 days | No high-risk antithrombotic therapy | 2.77 | 1.39 | 5.53 | 0.004 |
| Intracranial Hemorrhage | Time stratified: 1-30 days | High-risk antithrombotic therapy | 1.29 | 0.60 | 2.78 | 0.517 |
| **Intracranial Hemorrhage** | **Time stratified: 1-30 days** | **Interaction (atorvastatin × HRAT)** | **0.47** | **0.17** | **1.31** | **0.148** |
| Intracranial Hemorrhage | Time stratified: 31-91 days | No high-risk antithrombotic therapy | 0.52 | 0.24 | 1.14 | 0.101 |
| Intracranial Hemorrhage | Time stratified: 31-91 days | High-risk antithrombotic therapy | 0.88 | 0.38 | 2.01 | 0.757 |
| **Intracranial Hemorrhage** | **Time stratified: 31-91 days** | **Interaction (atorvastatin × HRAT)** | **1.69** | **0.54** | **5.31** | **0.368** |
| Intracranial Hemorrhage | Time stratified: 92-182 days | No high-risk antithrombotic therapy | 2.21 | 0.63 | 7.68 | 0.214 |
| Intracranial Hemorrhage | Time stratified: 92-182 days | High-risk antithrombotic therapy | 2.07 | 0.61 | 7.00 | 0.242 |
| **Intracranial Hemorrhage** | **Time stratified: 92-182 days** | **Interaction (atorvastatin × HRAT)** | **0.94** | **0.16** | **5.37** | **0.943** |

**Table S11 (continued) Effect of Atorvastatin on Confirmed Outcomes, Stratified by Concomitant High-Risk Antithrombotic Therapy (HRAT)**
|  |  |  |  |  |  |  |
| --- | --- | --- | --- | --- | --- | --- |
| Outcome | Stratum | Comparison | Weighted Fine-Gray |  |  |  |
|  |  |  | sHR | LCL | UCL | p |
| Nonrheumatic and unspecified valve disorders | overall | No high-risk antithrombotic therapy | 1.01 | 0.88 | 1.16 | 0.924 |
| Nonrheumatic and unspecified valve disorders | overall | High-risk antithrombotic therapy | 1.14 | 0.99 | 1.32 | 0.072 |
| **Nonrheumatic and unspecified valve disorders** | **overall** | **Interaction (atorvastatin × HRAT)** | **1.13** | **0.93** | **1.39** | **0.218** |
| Nonrheumatic and unspecified valve disorders | Age 65-74 years | No high-risk antithrombotic therapy | 0.92 | 0.73 | 1.16 | 0.474 |
| Nonrheumatic and unspecified valve disorders | Age 65-74 years | High-risk antithrombotic therapy | 1.22 | 0.96 | 1.54 | 0.100 |
| **Nonrheumatic and unspecified valve disorders** | **Age 65-74 years** | **Interaction (atorvastatin × HRAT)** | **1.33** | **0.95** | **1.85** | **0.095** |
| Nonrheumatic and unspecified valve disorders | Age 75+ years | No high-risk antithrombotic therapy | 1.09 | 0.92 | 1.29 | 0.333 |
| Nonrheumatic and unspecified valve disorders | Age 75+ years | High-risk antithrombotic therapy | 1.11 | 0.92 | 1.33 | 0.263 |
| **Nonrheumatic and unspecified valve disorders** | **Age 75+ years** | **Interaction (atorvastatin × HRAT)** | **1.02** | **0.80** | **1.31** | **0.871** |
| Nonrheumatic and unspecified valve disorders | Male | No high-risk antithrombotic therapy | 0.87 | 0.70 | 1.07 | 0.186 |
| Nonrheumatic and unspecified valve disorders | Male | High-risk antithrombotic therapy | 1.32 | 1.06 | 1.65 | 0.013 |
| **Nonrheumatic and unspecified valve disorders** | **Male** | **Interaction (atorvastatin × HRAT)** | **1.53** | **1.12** | **2.08** | **0.007** |
| Nonrheumatic and unspecified valve disorders | Female | No high-risk antithrombotic therapy | 1.14 | 0.95 | 1.37 | 0.150 |
| Nonrheumatic and unspecified valve disorders | Female | High-risk antithrombotic therapy | 1.02 | 0.84 | 1.24 | 0.828 |
| **Nonrheumatic and unspecified valve disorders** | **Female** | **Interaction (atorvastatin × HRAT)** | **0.89** | **0.69** | **1.16** | **0.405** |
| Nonrheumatic and unspecified valve disorders | White Race | No high-risk antithrombotic therapy | 1.00 | 0.86 | 1.17 | 0.977 |
| Nonrheumatic and unspecified valve disorders | White Race | High-risk antithrombotic therapy | 1.18 | 1.01 | 1.37 | 0.038 |
| **Nonrheumatic and unspecified valve disorders** | **White Race** | **Interaction (atorvastatin × HRAT)** | **1.17** | **0.94** | **1.46** | **0.150** |
| Nonrheumatic and unspecified valve disorders | Non-White Race | No high-risk antithrombotic therapy | 1.00 | 0.73 | 1.38 | 0.997 |
| Nonrheumatic and unspecified valve disorders | Non-White Race | High-risk antithrombotic therapy | 0.97 | 0.65 | 1.47 | 0.904 |
| **Nonrheumatic and unspecified valve disorders** | **Non-White Race** | **Interaction (atorvastatin × HRAT)** | **0.97** | **0.58** | **1.64** | **0.922** |
| Nonrheumatic and unspecified valve disorders | Time stratified: 1-30 days | No high-risk antithrombotic therapy | 0.97 | 0.80 | 1.18 | 0.760 |
| Nonrheumatic and unspecified valve disorders | Time stratified: 1-30 days | High-risk antithrombotic therapy | 1.00 | 0.83 | 1.21 | 0.979 |
| **Nonrheumatic and unspecified valve disorders** | **Time stratified: 1-30 days** | **Interaction (atorvastatin × HRAT)** | **1.03** | **0.79** | **1.36** | **0.813** |
| Nonrheumatic and unspecified valve disorders | Time stratified: 31-91 days | No high-risk antithrombotic therapy | 0.99 | 0.79 | 1.24 | 0.929 |
| Nonrheumatic and unspecified valve disorders | Time stratified: 31-91 days | High-risk antithrombotic therapy | 1.07 | 0.81 | 1.40 | 0.648 |
| **Nonrheumatic and unspecified valve disorders** | **Time stratified: 31-91 days** | **Interaction (atorvastatin × HRAT)** | **1.08** | **0.76** | **1.53** | **0.682** |
| Nonrheumatic and unspecified valve disorders | Time stratified: 92-182 days | No high-risk antithrombotic therapy | 1.06 | 0.73 | 1.53 | 0.759 |
| Nonrheumatic and unspecified valve disorders | Time stratified: 92-182 days | High-risk antithrombotic therapy | 1.97 | 1.32 | 2.94 | 0.001 |
| **Nonrheumatic and unspecified valve disorders** | **Time stratified: 92-182 days** | **Interaction (atorvastatin × HRAT)** | **1.86** | **1.08** | **3.21** | **0.026** |

**Table S11 (continued) Effect of Atorvastatin on Confirmed Outcomes, Stratified by Concomitant High-Risk Antithrombotic Therapy (HRAT)**
|  |  |  |  |  |  |  |
| --- | --- | --- | --- | --- | --- | --- |
| Outcome | Stratum | Comparison | Weighted Fine-Gray |  |  |  |
|  |  |  | sHR | LCL | UCL | p |
| Nonrheumatic and unspecified valve disorders plus imaging | overall | No high-risk antithrombotic therapy | 1.02 | 0.88 | 1.18 | 0.778 |
| Nonrheumatic and unspecified valve disorders plus imaging | overall | High-risk antithrombotic therapy | 1.15 | 0.99 | 1.34 | 0.060 |
| **Nonrheumatic and unspecified valve disorders plus imaging** | **overall** | **Interaction (atorvastatin × HRAT)** | **1.13** | **0.92** | **1.39** | **0.249** |
| Nonrheumatic and unspecified valve disorders plus imaging | Age 65-74 years | No high-risk antithrombotic therapy | 0.93 | 0.74 | 1.18 | 0.572 |
| Nonrheumatic and unspecified valve disorders plus imaging | Age 65-74 years | High-risk antithrombotic therapy | 1.23 | 0.97 | 1.57 | 0.094 |
| **Nonrheumatic and unspecified valve disorders plus imaging** | **Age 65-74 years** | **Interaction (atorvastatin × HRAT)** | **1.32** | **0.94** | **1.85** | **0.112** |
| Nonrheumatic and unspecified valve disorders plus imaging | Age 75+ years | No high-risk antithrombotic therapy | 1.10 | 0.92 | 1.31 | 0.283 |
| Nonrheumatic and unspecified valve disorders plus imaging | Age 75+ years | High-risk antithrombotic therapy | 1.12 | 0.93 | 1.35 | 0.232 |
| **Nonrheumatic and unspecified valve disorders plus imaging** | **Age 75+ years** | **Interaction (atorvastatin × HRAT)** | **1.02** | **0.79** | **1.32** | **0.886** |
| Nonrheumatic and unspecified valve disorders plus imaging | Male | No high-risk antithrombotic therapy | 0.90 | 0.73 | 1.13 | 0.368 |
| Nonrheumatic and unspecified valve disorders plus imaging | Male | High-risk antithrombotic therapy | 1.37 | 1.09 | 1.72 | 0.007 |
| **Nonrheumatic and unspecified valve disorders plus imaging** | **Male** | **Interaction (atorvastatin × HRAT)** | **1.52** | **1.10** | **2.08** | **0.010** |
| Nonrheumatic and unspecified valve disorders plus imaging | Female | No high-risk antithrombotic therapy | 1.13 | 0.94 | 1.36 | 0.196 |
| Nonrheumatic and unspecified valve disorders plus imaging | Female | High-risk antithrombotic therapy | 1.01 | 0.83 | 1.23 | 0.930 |
| **Nonrheumatic and unspecified valve disorders plus imaging** | **Female** | **Interaction (atorvastatin × HRAT)** | **0.89** | **0.68** | **1.17** | **0.409** |
| Nonrheumatic and unspecified valve disorders plus imaging | White Race | No high-risk antithrombotic therapy | 1.02 | 0.87 | 1.19 | 0.852 |
| Nonrheumatic and unspecified valve disorders plus imaging | White Race | High-risk antithrombotic therapy | 1.20 | 1.02 | 1.40 | 0.024 |
| **Nonrheumatic and unspecified valve disorders plus imaging** | **White Race** | **Interaction (atorvastatin × HRAT)** | **1.18** | **0.94** | **1.48** | **0.146** |
| Nonrheumatic and unspecified valve disorders plus imaging | Non-White Race | No high-risk antithrombotic therapy | 1.02 | 0.73 | 1.41 | 0.917 |
| Nonrheumatic and unspecified valve disorders plus imaging | Non-White Race | High-risk antithrombotic therapy | 0.93 | 0.62 | 1.41 | 0.746 |
| **Nonrheumatic and unspecified valve disorders plus imaging** | **Non-White Race** | **Interaction (atorvastatin × HRAT)** | **0.92** | **0.54** | **1.55** | **0.750** |
| Nonrheumatic and unspecified valve disorders plus imaging | Time stratified: 1-30 days | No high-risk antithrombotic therapy | 0.98 | 0.80 | 1.20 | 0.863 |
| Nonrheumatic and unspecified valve disorders plus imaging | Time stratified: 1-30 days | High-risk antithrombotic therapy | 1.00 | 0.83 | 1.22 | 0.970 |
| **Nonrheumatic and unspecified valve disorders plus imaging** | **Time stratified: 1-30 days** | **Interaction (atorvastatin × HRAT)** | **1.02** | **0.77** | **1.35** | **0.881** |
| Nonrheumatic and unspecified valve disorders plus imaging | Time stratified: 31-91 days | No high-risk antithrombotic therapy | 0.99 | 0.79 | 1.25 | 0.950 |
| Nonrheumatic and unspecified valve disorders plus imaging | Time stratified: 31-91 days | High-risk antithrombotic therapy | 1.10 | 0.83 | 1.45 | 0.529 |
| **Nonrheumatic and unspecified valve disorders plus imaging** | **Time stratified: 31-91 days** | **Interaction (atorvastatin × HRAT)** | **1.10** | **0.77** | **1.59** | **0.596** |
| Nonrheumatic and unspecified valve disorders plus imaging | Time stratified: 92-182 days | No high-risk antithrombotic therapy | 1.08 | 0.74 | 1.58 | 0.673 |
| Nonrheumatic and unspecified valve disorders plus imaging | Time stratified: 92-182 days | High-risk antithrombotic therapy | 1.98 | 1.31 | 2.98 | 0.001 |
| **Nonrheumatic and unspecified valve disorders plus imaging** | **Time stratified: 92-182 days** | **Interaction (atorvastatin × HRAT)** | **1.82** | **1.05** | **3.18** | **0.034** |

**Table S11 (continued) Effect of Atorvastatin on Confirmed Outcomes, Stratified by Concomitant High-Risk Antithrombotic Therapy (HRAT)**
|  |  |  |  |  |  |  |
| --- | --- | --- | --- | --- | --- | --- |
| Outcome | Stratum | Comparison | Weighted Fine-Gray |  |  |  |
|  |  |  | sHR | LCL | UCL | p |
| Valve intervention | overall | No high-risk antithrombotic therapy | 2.24 | 1.06 | 4.73 | 0.034 |
| Valve intervention | overall | High-risk antithrombotic therapy | 1.56 | 0.82 | 2.96 | 0.177 |
| **Valve intervention** | **overall** | **Interaction (atorvastatin × HRAT)** | **0.69** | **0.26** | **1.86** | **0.468** |
| Valve intervention | Age 65-74 years | No high-risk antithrombotic therapy | 1.87 | 0.60 | 5.81 | 0.277 |
| Valve intervention | Age 65-74 years | High-risk antithrombotic therapy | 2.81 | 0.83 | 9.53 | 0.097 |
| **Valve intervention** | **Age 65-74 years** | **Interaction (atorvastatin × HRAT)** | **1.50** | **0.29** | **7.83** | **0.630** |
| Valve intervention | Age 75+ years | No high-risk antithrombotic therapy | 2.70 | 1.07 | 6.80 | 0.035 |
| Valve intervention | Age 75+ years | High-risk antithrombotic therapy | 1.18 | 0.55 | 2.54 | 0.666 |
| **Valve intervention** | **Age 75+ years** | **Interaction (atorvastatin × HRAT)** | **0.44** | **0.13** | **1.47** | **0.181** |
| Valve intervention | Male | No high-risk antithrombotic therapy | 1.60 | 0.61 | 4.20 | 0.337 |
| Valve intervention | Male | High-risk antithrombotic therapy | 3.32 | 1.09 | 10.06 | 0.034 |
| **Valve intervention** | **Male** | **Interaction (atorvastatin × HRAT)** | **2.07** | **0.48** | **8.92** | **0.330** |
| Valve intervention | Female | No high-risk antithrombotic therapy | 3.74 | 1.25 | 11.22 | 0.018 |
| Valve intervention | Female | High-risk antithrombotic therapy | 0.90 | 0.40 | 2.01 | 0.796 |
| **Valve intervention** | **Female** | **Interaction (atorvastatin × HRAT)** | **0.24** | **0.06** | **0.94** | **0.041** |
| Valve intervention | White Race | No high-risk antithrombotic therapy | 2.34 | 1.01 | 5.41 | 0.046 |
| Valve intervention | White Race | High-risk antithrombotic therapy | 2.14 | 1.05 | 4.36 | 0.035 |
| **Valve intervention** | **White Race** | **Interaction (atorvastatin × HRAT)** | **0.92** | **0.31** | **2.73** | **0.874** |
| Valve intervention | Non-White Race | No high-risk antithrombotic therapy | 1.64 | 0.32 | 8.34 | 0.554 |
| Valve intervention | Non-White Race | High-risk antithrombotic therapy | 0.45 | 0.11 | 1.83 | 0.261 |
| **Valve intervention** | **Non-White Race** | **Interaction (atorvastatin × HRAT)** | **0.27** | **0.03** | **2.38** | **0.239** |
| Valve intervention | Time stratified: 1-30 days | No high-risk antithrombotic therapy | 3.27 | 1.04 | 10.29 | 0.043 |
| Valve intervention | Time stratified: 1-30 days | High-risk antithrombotic therapy | 1.32 | 0.58 | 3.00 | 0.513 |
| **Valve intervention** | **Time stratified: 1-30 days** | **Interaction (atorvastatin × HRAT)** | **0.40** | **0.10** | **1.66** | **0.207** |
| Valve intervention | Time stratified: 31-91 days | No high-risk antithrombotic therapy | 2.67 | 0.77 | 9.21 | 0.120 |
| Valve intervention | Time stratified: 31-91 days | High-risk antithrombotic therapy | 4.36 | 0.59 | 32.12 | 0.149 |
| **Valve intervention** | **Time stratified: 31-91 days** | **Interaction (atorvastatin × HRAT)** | **1.63** | **0.16** | **17.09** | **0.682** |
| Valve intervention | Time stratified: 92-182 days | No high-risk antithrombotic therapy | 0.85 | 0.21 | 3.50 | 0.825 |
| Valve intervention | Time stratified: 92-182 days | High-risk antithrombotic therapy | 2.21 | 0.60 | 8.10 | 0.233 |
| **Valve intervention** | **Time stratified: 92-182 days** | **Interaction (atorvastatin × HRAT)** | **2.59** | **0.38** | **17.54** | **0.330** |

**Table S11 (continued) Effect of Atorvastatin on Confirmed Outcomes, Stratified by Concomitant High-Risk Antithrombotic Therapy (HRAT)**
|  |  |  |  |  |  |  |
| --- | --- | --- | --- | --- | --- | --- |
| Outcome | Stratum | Comparison | Weighted Fine-Gray |  |  |  |
|  |  |  | sHR | LCL | UCL | p |
| Composite: valve | overall | No high-risk antithrombotic therapy | 1.01 | 0.88 | 1.16 | 0.906 |
| Composite: valve | overall | High-risk antithrombotic therapy | 1.14 | 0.99 | 1.32 | 0.071 |
| **Composite: valve** | **overall** | **Interaction (atorvastatin × HRAT)** | **1.13** | **0.93** | **1.38** | **0.225** |
| Composite: valve | Age 65-74 years | No high-risk antithrombotic therapy | 0.91 | 0.73 | 1.15 | 0.430 |
| Composite: valve | Age 65-74 years | High-risk antithrombotic therapy | 1.21 | 0.96 | 1.52 | 0.115 |
| **Composite: valve** | **Age 65-74 years** | **Interaction (atorvastatin × HRAT)** | **1.32** | **0.95** | **1.84** | **0.094** |
| Composite: valve | Age 75+ years | No high-risk antithrombotic therapy | 1.10 | 0.93 | 1.30 | 0.282 |
| Composite: valve | Age 75+ years | High-risk antithrombotic therapy | 1.12 | 0.93 | 1.34 | 0.235 |
| **Composite: valve** | **Age 75+ years** | **Interaction (atorvastatin × HRAT)** | **1.02** | **0.79** | **1.30** | **0.891** |
| Composite: valve | Male | No high-risk antithrombotic therapy | 0.86 | 0.70 | 1.06 | 0.158 |
| Composite: valve | Male | High-risk antithrombotic therapy | 1.32 | 1.06 | 1.64 | 0.014 |
| **Composite: valve** | **Male** | **Interaction (atorvastatin × HRAT)** | **1.53** | **1.13** | **2.08** | **0.006** |
| Composite: valve | Female | No high-risk antithrombotic therapy | 1.15 | 0.96 | 1.38 | 0.120 |
| Composite: valve | Female | High-risk antithrombotic therapy | 1.03 | 0.85 | 1.24 | 0.795 |
| **Composite: valve** | **Female** | **Interaction (atorvastatin × HRAT)** | **0.89** | **0.68** | **1.16** | **0.376** |
| Composite: valve | White Race | No high-risk antithrombotic therapy | 1.00 | 0.86 | 1.17 | 0.970 |
| Composite: valve | White Race | High-risk antithrombotic therapy | 1.18 | 1.01 | 1.37 | 0.037 |
| **Composite: valve** | **White Race** | **Interaction (atorvastatin × HRAT)** | **1.17** | **0.94** | **1.46** | **0.150** |
| Composite: valve | Non-White Race | No high-risk antithrombotic therapy | 1.01 | 0.73 | 1.39 | 0.970 |
| Composite: valve | Non-White Race | High-risk antithrombotic therapy | 0.97 | 0.64 | 1.46 | 0.882 |
| **Composite: valve** | **Non-White Race** | **Interaction (atorvastatin × HRAT)** | **0.96** | **0.57** | **1.62** | **0.889** |
| Composite: valve | Time stratified: 1-30 days | No high-risk antithrombotic therapy | 0.98 | 0.80 | 1.19 | 0.807 |
| Composite: valve | Time stratified: 1-30 days | High-risk antithrombotic therapy | 1.00 | 0.83 | 1.21 | 0.989 |
| **Composite: valve** | **Time stratified: 1-30 days** | **Interaction (atorvastatin × HRAT)** | **1.02** | **0.78** | **1.34** | **0.869** |
| Composite: valve | Time stratified: 31-91 days | No high-risk antithrombotic therapy | 1.00 | 0.80 | 1.25 | 0.974 |
| Composite: valve | Time stratified: 31-91 days | High-risk antithrombotic therapy | 1.08 | 0.82 | 1.42 | 0.581 |
| **Composite: valve** | **Time stratified: 31-91 days** | **Interaction (atorvastatin × HRAT)** | **1.08** | **0.76** | **1.54** | **0.655** |
| Composite: valve | Time stratified: 92-182 days | No high-risk antithrombotic therapy | 1.02 | 0.71 | 1.47 | 0.899 |
| Composite: valve | Time stratified: 92-182 days | High-risk antithrombotic therapy | 1.92 | 1.30 | 2.84 | 0.001 |
| **Composite: valve** | **Time stratified: 92-182 days** | **Interaction (atorvastatin × HRAT)** | **1.87** | **1.10** | **3.19** | **0.021** |

**Table S11 (continued) Effect of Atorvastatin on Confirmed Outcomes, Stratified by Concomitant High-Risk Antithrombotic Therapy (HRAT)**
|  |  |  |  |  |  |  |
| --- | --- | --- | --- | --- | --- | --- |
| Outcome | Stratum | Comparison | Weighted Fine-Gray |  |  |  |
|  |  |  | sHR | LCL | UCL | p |
| Sprains and strains, initial encounter | overall | No high-risk antithrombotic therapy | 1.11 | 0.84 | 1.47 | 0.460 |
| Sprains and strains, initial encounter | overall | High-risk antithrombotic therapy | 1.40 | 1.03 | 1.91 | 0.032 |
| **Sprains and strains, initial encounter** | **overall** | **Interaction (atorvastatin × HRAT)** | **1.26** | **0.83** | **1.91** | **0.271** |
| Sprains and strains, initial encounter | Age 65-74 years | No high-risk antithrombotic therapy | 1.34 | 0.88 | 2.03 | 0.176 |
| Sprains and strains, initial encounter | Age 65-74 years | High-risk antithrombotic therapy | 1.27 | 0.81 | 1.99 | 0.298 |
| **Sprains and strains, initial encounter** | **Age 65-74 years** | **Interaction (atorvastatin × HRAT)** | **0.95** | **0.51** | **1.76** | **0.872** |
| Sprains and strains, initial encounter | Age 75+ years | No high-risk antithrombotic therapy | 0.98 | 0.68 | 1.42 | 0.932 |
| Sprains and strains, initial encounter | Age 75+ years | High-risk antithrombotic therapy | 1.52 | 0.99 | 2.33 | 0.054 |
| **Sprains and strains, initial encounter** | **Age 75+ years** | **Interaction (atorvastatin × HRAT)** | **1.55** | **0.88** | **2.71** | **0.129** |
| Sprains and strains, initial encounter | Male | No high-risk antithrombotic therapy | 1.38 | 0.88 | 2.16 | 0.163 |
| Sprains and strains, initial encounter | Male | High-risk antithrombotic therapy | 2.12 | 1.21 | 3.74 | 0.009 |
| **Sprains and strains, initial encounter** | **Male** | **Interaction (atorvastatin × HRAT)** | **1.54** | **0.75** | **3.17** | **0.239** |
| Sprains and strains, initial encounter | Female | No high-risk antithrombotic therapy | 1.00 | 0.70 | 1.42 | 1.000 |
| Sprains and strains, initial encounter | Female | High-risk antithrombotic therapy | 1.10 | 0.76 | 1.60 | 0.609 |
| **Sprains and strains, initial encounter** | **Female** | **Interaction (atorvastatin × HRAT)** | **1.10** | **0.66** | **1.84** | **0.710** |
| Sprains and strains, initial encounter | White Race | No high-risk antithrombotic therapy | 1.06 | 0.80 | 1.41 | 0.698 |
| Sprains and strains, initial encounter | White Race | High-risk antithrombotic therapy | 1.37 | 0.98 | 1.90 | 0.062 |
| **Sprains and strains, initial encounter** | **White Race** | **Interaction (atorvastatin × HRAT)** | **1.29** | **0.84** | **2.00** | **0.247** |
| Sprains and strains, initial encounter | Non-White Race | No high-risk antithrombotic therapy | 1.41 | 0.57 | 3.50 | 0.463 |
| Sprains and strains, initial encounter | Non-White Race | High-risk antithrombotic therapy | 1.69 | 0.68 | 4.21 | 0.258 |
| **Sprains and strains, initial encounter** | **Non-White Race** | **Interaction (atorvastatin × HRAT)** | **1.20** | **0.33** | **4.33** | **0.777** |
| Sprains and strains, initial encounter | Time stratified: 1-30 days | No high-risk antithrombotic therapy | 0.97 | 0.61 | 1.55 | 0.902 |
| Sprains and strains, initial encounter | Time stratified: 1-30 days | High-risk antithrombotic therapy | 1.45 | 0.80 | 2.62 | 0.221 |
| **Sprains and strains, initial encounter** | **Time stratified: 1-30 days** | **Interaction (atorvastatin × HRAT)** | **1.49** | **0.70** | **3.17** | **0.300** |
| Sprains and strains, initial encounter | Time stratified: 31-91 days | No high-risk antithrombotic therapy | 1.11 | 0.68 | 1.80 | 0.689 |
| Sprains and strains, initial encounter | Time stratified: 31-91 days | High-risk antithrombotic therapy | 1.58 | 0.94 | 2.66 | 0.086 |
| **Sprains and strains, initial encounter** | **Time stratified: 31-91 days** | **Interaction (atorvastatin × HRAT)** | **1.43** | **0.70** | **2.92** | **0.329** |
| Sprains and strains, initial encounter | Time stratified: 92-182 days | No high-risk antithrombotic therapy | 1.04 | 0.63 | 1.70 | 0.882 |
| Sprains and strains, initial encounter | Time stratified: 92-182 days | High-risk antithrombotic therapy | 1.18 | 0.71 | 1.93 | 0.525 |
| **Sprains and strains, initial encounter** | **Time stratified: 92-182 days** | **Interaction (atorvastatin × HRAT)** | **1.13** | **0.56** | **2.28** | **0.728** |

**Table S11 (continued) Effect of Atorvastatin on Confirmed Outcomes, Stratified by Concomitant High-Risk Antithrombotic Therapy (HRAT)**
|  |  |  |  |  |  |  |
| --- | --- | --- | --- | --- | --- | --- |
| Outcome | Stratum | Comparison | Weighted Fine-Gray |  |  |  |
|  |  |  | sHR | LCL | UCL | p |
| Acute hepatic failure | overall | No high-risk antithrombotic therapy | 1.47 | 0.89 | 2.43 | 0.130 |
| Acute hepatic failure | overall | High-risk antithrombotic therapy | 2.19 | 1.11 | 4.31 | 0.024 |
| **Acute hepatic failure** | **overall** | **Interaction (atorvastatin × HRAT)** | **1.49** | **0.64** | **3.46** | **0.357** |
| Acute hepatic failure | Age 65-74 years | No high-risk antithrombotic therapy | 1.46 | 0.67 | 3.19 | 0.345 |
| Acute hepatic failure | Age 65-74 years | High-risk antithrombotic therapy | 3.11 | 0.96 | 10.06 | 0.058 |
| **Acute hepatic failure** | **Age 65-74 years** | **Interaction (atorvastatin × HRAT)** | **2.13** | **0.51** | **8.89** | **0.298** |
| Acute hepatic failure | Age 75+ years | No high-risk antithrombotic therapy | 1.49 | 0.78 | 2.85 | 0.228 |
| Acute hepatic failure | Age 75+ years | High-risk antithrombotic therapy | 1.85 | 0.81 | 4.24 | 0.147 |
| **Acute hepatic failure** | **Age 75+ years** | **Interaction (atorvastatin × HRAT)** | **1.24** | **0.43** | **3.55** | **0.688** |
| Acute hepatic failure | Male | No high-risk antithrombotic therapy | 0.95 | 0.47 | 1.91 | 0.892 |
| Acute hepatic failure | Male | High-risk antithrombotic therapy | 1.92 | 0.80 | 4.63 | 0.144 |
| **Acute hepatic failure** | **Male** | **Interaction (atorvastatin × HRAT)** | **2.02** | **0.66** | **6.21** | **0.220** |
| Acute hepatic failure | Female | No high-risk antithrombotic therapy | 2.15 | 1.08 | 4.28 | 0.029 |
| Acute hepatic failure | Female | High-risk antithrombotic therapy | 2.59 | 0.90 | 7.41 | 0.076 |
| **Acute hepatic failure** | **Female** | **Interaction (atorvastatin × HRAT)** | **1.20** | **0.34** | **4.25** | **0.773** |
| Acute hepatic failure | White Race | No high-risk antithrombotic therapy | 1.24 | 0.73 | 2.10 | 0.431 |
| Acute hepatic failure | White Race | High-risk antithrombotic therapy | 1.91 | 0.94 | 3.89 | 0.073 |
| **Acute hepatic failure** | **White Race** | **Interaction (atorvastatin × HRAT)** | **1.55** | **0.64** | **3.76** | **0.337** |
| Acute hepatic failure | Non-White Race | No high-risk antithrombotic therapy | 5.50 | 0.72 | 41.92 | 0.100 |
| Acute hepatic failure | Non-White Race | High-risk antithrombotic therapy | 6.58 | 0.86 | 50.35 | 0.070 |
| **Acute hepatic failure** | **Non-White Race** | **Interaction (atorvastatin × HRAT)** | **1.20** | **0.07** | **20.95** | **0.902** |
| Acute hepatic failure | Time stratified: 1-30 days | No high-risk antithrombotic therapy | 1.10 | 0.53 | 2.25 | 0.800 |
| Acute hepatic failure | Time stratified: 1-30 days | High-risk antithrombotic therapy | 1.48 | 0.63 | 3.48 | 0.364 |
| **Acute hepatic failure** | **Time stratified: 1-30 days** | **Interaction (atorvastatin × HRAT)** | **1.35** | **0.44** | **4.13** | **0.597** |
| Acute hepatic failure | Time stratified: 31-91 days | No high-risk antithrombotic therapy | 2.75 | 1.12 | 6.72 | 0.027 |
| Acute hepatic failure | Time stratified: 31-91 days | High-risk antithrombotic therapy | 4.58 | 1.40 | 14.99 | 0.012 |
| **Acute hepatic failure** | **Time stratified: 31-91 days** | **Interaction (atorvastatin × HRAT)** | **1.67** | **0.38** | **7.31** | **0.499** |
| Acute hepatic failure | Time stratified: 92-182 days | No high-risk antithrombotic therapy | 0.62 | 0.24 | 1.59 | 0.318 |
| Acute hepatic failure | Time stratified: 92-182 days | High-risk antithrombotic therapy | 4.41 | 0.60 | 32.44 | 0.145 |
| **Acute hepatic failure** | **Time stratified: 92-182 days** | **Interaction (atorvastatin × HRAT)** | **7.15** | **0.78** | **65.17** | **0.081** |

**Table S11 (continued) Effect of Atorvastatin on Confirmed Outcomes, Stratified by Concomitant High-Risk Antithrombotic Therapy (HRAT)**
|  |  |  |  |  |  |  |
| --- | --- | --- | --- | --- | --- | --- |
| Outcome | Stratum | Comparison | Weighted Fine-Gray |  |  |  |
|  |  |  | sHR | LCL | UCL | p |
| Biliary tract disease | overall | No high-risk antithrombotic therapy | 1.03 | 0.82 | 1.29 | 0.812 |
| Biliary tract disease | overall | High-risk antithrombotic therapy | 1.43 | 1.07 | 1.91 | 0.015 |
| **Biliary tract disease** | **overall** | **Interaction (atorvastatin × HRAT)** | **1.39** | **0.97** | **2.01** | **0.076** |
| Biliary tract disease | Age 65-74 years | No high-risk antithrombotic therapy | 1.04 | 0.70 | 1.54 | 0.864 |
| Biliary tract disease | Age 65-74 years | High-risk antithrombotic therapy | 1.05 | 0.68 | 1.63 | 0.824 |
| **Biliary tract disease** | **Age 65-74 years** | **Interaction (atorvastatin × HRAT)** | **1.02** | **0.56** | **1.83** | **0.960** |
| Biliary tract disease | Age 75+ years | No high-risk antithrombotic therapy | 1.04 | 0.79 | 1.38 | 0.778 |
| Biliary tract disease | Age 75+ years | High-risk antithrombotic therapy | 1.82 | 1.25 | 2.66 | 0.002 |
| **Biliary tract disease** | **Age 75+ years** | **Interaction (atorvastatin × HRAT)** | **1.75** | **1.10** | **2.80** | **0.019** |
| Biliary tract disease | Male | No high-risk antithrombotic therapy | 0.87 | 0.62 | 1.23 | 0.437 |
| Biliary tract disease | Male | High-risk antithrombotic therapy | 1.13 | 0.76 | 1.68 | 0.536 |
| **Biliary tract disease** | **Male** | **Interaction (atorvastatin × HRAT)** | **1.30** | **0.77** | **2.18** | **0.324** |
| Biliary tract disease | Female | No high-risk antithrombotic therapy | 1.19 | 0.88 | 1.60 | 0.260 |
| Biliary tract disease | Female | High-risk antithrombotic therapy | 1.89 | 1.24 | 2.88 | 0.003 |
| **Biliary tract disease** | **Female** | **Interaction (atorvastatin × HRAT)** | **1.59** | **0.95** | **2.66** | **0.080** |
| Biliary tract disease | White Race | No high-risk antithrombotic therapy | 1.06 | 0.83 | 1.36 | 0.630 |
| Biliary tract disease | White Race | High-risk antithrombotic therapy | 1.40 | 1.03 | 1.92 | 0.034 |
| **Biliary tract disease** | **White Race** | **Interaction (atorvastatin × HRAT)** | **1.32** | **0.89** | **1.96** | **0.171** |
| Biliary tract disease | Non-White Race | No high-risk antithrombotic therapy | 0.93 | 0.54 | 1.60 | 0.795 |
| Biliary tract disease | Non-White Race | High-risk antithrombotic therapy | 1.66 | 0.78 | 3.50 | 0.186 |
| **Biliary tract disease** | **Non-White Race** | **Interaction (atorvastatin × HRAT)** | **1.78** | **0.71** | **4.46** | **0.218** |
| Biliary tract disease | Time stratified: 1-30 days | No high-risk antithrombotic therapy | 1.19 | 0.86 | 1.65 | 0.285 |
| Biliary tract disease | Time stratified: 1-30 days | High-risk antithrombotic therapy | 1.36 | 0.88 | 2.10 | 0.171 |
| **Biliary tract disease** | **Time stratified: 1-30 days** | **Interaction (atorvastatin × HRAT)** | **1.14** | **0.66** | **1.96** | **0.643** |
| Biliary tract disease | Time stratified: 31-91 days | No high-risk antithrombotic therapy | 0.96 | 0.63 | 1.47 | 0.856 |
| Biliary tract disease | Time stratified: 31-91 days | High-risk antithrombotic therapy | 2.08 | 1.18 | 3.68 | 0.012 |
| **Biliary tract disease** | **Time stratified: 31-91 days** | **Interaction (atorvastatin × HRAT)** | **2.16** | **1.07** | **4.39** | **0.033** |
| Biliary tract disease | Time stratified: 92-182 days | No high-risk antithrombotic therapy | 0.72 | 0.45 | 1.17 | 0.186 |
| Biliary tract disease | Time stratified: 92-182 days | High-risk antithrombotic therapy | 0.96 | 0.57 | 1.61 | 0.869 |
| **Biliary tract disease** | **Time stratified: 92-182 days** | **Interaction (atorvastatin × HRAT)** | **1.33** | **0.65** | **2.70** | **0.437** |

HRAT = high-risk antithrombotic therapy, defined as dispensing on the trial start date of clopidogrel, ticagrelor, prasugrel, warfarin, rivaroxaban, apixaban, dabigatran, edoxaban, enoxaparin, or unfractionated heparin. sHR = subdistribution hazard ratio from inverse-probability-weighted Fine-Gray models treating death as a competing risk; LCL/UCL = lower/upper 95% confidence limits. “No high-risk antithrombotic therapy” and “High-risk antithrombotic therapy” are the estimated effects of atorvastatin in the absence and presence of HRAT, respectively; “Interaction (atorvastatin × HRAT)” is the multiplicative interaction term.

